# How much is enough? Optimising sampling frames for genomic surveillance of *Escherichia coli* and *Klebsiella* spp. bloodstream infections – a retrospective study

**DOI:** 10.64898/2026.09.28.26364154

**Authors:** Dorottya Nagy, The NEKSUS Consortium, Susan Hopkins, Derrick Crook, Ann Sarah Walker, Julie Robotham, Katie L. Hopkins, Alice Ledda, Daniel J. Wilson, David Williams, Russell Hope, Colin S. Brown, Nicole Stoesser, Samuel Lipworth

**Author notes:** Group authorship and affiliations listed in acknowledgements (The NEKSUS Consortium). Joint senior authors (Nicole Stoesser, Samuel Lipworth). **Corresponding author:** Dorottya Nagy, Modernising Medical Microbiology Unit, Experimental Medicine Division, Nuffield Department of Medicine, Level 5, Room 5800, John Radcliffe Hospital, Headley Way, Oxford, OX3 9DU. **Disclaimers:** None.

## Abstract

**Background:** Optimising sampling frames for genomic surveillance of *E. coli* and *Klebsiella* may support interventions to mitigate bloodstream infections (BSIs), but approaches to estimating sample size and how these relate to bacterial population diversity at multiple genetic levels (strain/plasmid/antimicrobial resistance genes [ARGs]) are lacking.

**Methods:** Using systematically collected, regionally-stratified, whole genome sequencing data from a 6-month genomic survey of English *E. coli*/*Klebsiella* BSI isolates (n=1,939; NEKSUS), we used Bayesian approaches to estimate the sample sizes required to capture genetic diversity at strain- (MLST, fastBAPS clusters), plasmid- and ARG-levels, using two diversity measures: ‘coverage’ (i.e. proportion of the bacterial population/plasmids/ARGs represented by features observed in the sample), and the number/proportion of unique features observed at varying surveillance sample sizes. Our method is available as a user-friendly R Shiny application, EpiSENTRY (https://01a0c3dd-90d2-e597-58a5-51391a468387.share.connect.posit.cloud/).

**Findings:** Randomly sampling 1,400 *E. coli*/*Klebsiella* BSI isolates each achieved ≥80% coverage of bacterial lineages and ARGs (i.e. capture features constituting ≥80% of the population) at 95% certainty, but captured lower proportions of unique features (28% MLSTs, 59% fastBAPS clusters, 19% plasmid subcommunities, and 57% ARGs for *E. coli;* 49%, 87%, 50% and 83% for *Klebsiella*, respectively). Sample sizes required for 80% coverage of *E. coli* plasmid populations were higher than MLSTs (1,928[95% CrI:1,837–2,025] vs 1,351[1,230–1,472]), while for *Klebsiella*, estimates were lower for plasmids (1,115[1,052–1,181] vs 1,422[1,331–1,515] for MLSTs). Lower sample sizes adequately captured 80% coverage of fastBAPS clusters and ARGs (649[575 – 727] and 27[23–30], respectively, for *E. coli*; 113[84–145] and 83[72-96] for *Klebsiella*), due to feature-specific frequency distributions. *Klebsiella* BSIs were more diverse than *E. coli*, with sampling 10.9% vs 3.2% of total annual BSIs in England needed to reach 80% MLST coverage. Plasmid populations and *Klebsiella* MLSTs were region-specific, emphasising the need for regionally-representative sampling.

**Interpretation:** Bayesian analysis facilitates estimates of context-specific genomic surveillance sample sizes. Cost-effective targets for coverage require further optimisation.

**Funding:** National Institute for Health and Social Care Research (NIHR) Health Protection Research Unit in Healthcare Associated Infections and Antimicrobial Resistance (NIHR207397); UKHSA and NIHR Oxford Biomedical Research Centre (BRC); UKHSA PhD Funding Competition; Robertson Fellowship; cloud compute infrastructure for this work was donated by Oracle Corporation Infrastructure.

**Research in context:** *Evidence before this study:* A MEDLINE (Ovid) search for genomic surveillance sample size estimation (“sample size estimation/calculation” AND “surveillance”; no restrictions on publication type, date or language) retrieved 62 records (22 included). Most studies (68% [15/22]) used calculations based on frequentist formulae, which require a user-specified arbitrary minimum frequency threshold for the feature/disease under surveillance. Surveillance outcomes were mostly prevalence/incidence (59% [13/22]) and presence/absence (54% [12/22]) of diseases/genomic features, and all studies considered diseases/features individually. Re-analyses of COVID-19 variant surveillance in high-income settings suggested that one third of the sequencing effort would have sufficed. Several publications highlight estimates should be tailored to specific surveillance goals. No studies extended surveillance sample size calculations to capturing population diversity of multiple genomic features simultaneously (e.g. plasmids, antimicrobial resistance [AMR] genes [ARGs]), although these may be important for bacterial genomic surveillance. Bayesian estimation is a powerful method of statistical inference where prior beliefs about the distribution of a population parameter (e.g.: the frequency of a certain genomic feature) can be updated to a more realistic posterior distribution using the likelihood of the available data, thus incorporating uncertainty from potentially noisy data.

*Added value of this study:* We applied Bayesian analysis to large-scale genomic data from a 6-month nationally-representative survey of English bloodstream infection-associated *E. coli* and *Klebsiella* to estimate the posterior frequency distribution, with uncertainty, of four genomic features (multi-locus sequence type [MLST], fastBAPS cluster, plasmid subcommunities, ARGs). We used estimates to inform meaningful minimum feature frequency thresholds for conventional sample size calculations, by relating frequency to proportion of population diversity captured (i.e.: coverage – the proportion of the population represented by the features detected at least once in the sample), as well as number/proportion of unique features captured. This method overcomes limitations of ecological estimators when scaling to large populations by using all survey data to inform estimates, while placing more weight on epidemiologically important common features. We have made our method available as a user-friendly R Shiny application, EpiSENTRY (https://01a0c3dd-90d2-e597-58a5-51391a468387.share.connect.posit.cloud/), allowing users to easily estimate sample sizes from their own data.

*Implications of all the available evidence:* Randomly sampling 1,400 *E. coli* and *Klebsiella* bloodstream (BSI) isolates, each, in England sufficiently captured strains and ARGs representing 80% of the estimated total bacterial population (80% coverage) at 95% certainty. While this captured the most common BSI-associated bacterial genomic features, >51% of unique MLSTs, >50% of plasmid subcommunities, >13% of fastBAPS clusters, and >17% of ARGs may remain undetected even at high coverages because there are so many unique variants at all genomic levels. Target coverage thresholds may need to be adjusted, or alternative targeted sequencing methods used (e.g. selection based on phenotype) for rare features of critical public health importance (e.g. carbapenemases). *Klebsiella* BSIs were more diverse than *E. coli*, relative to incidence, requiring 10.9% vs 3.2% of BSIs to be sequenced to capture comparable diversity. Plasmid populations and *Klebsiella* MLSTs were also region-specific, emphasising the need for regionally-representative sampling. Sample size estimates should be context-specific to best support national, regional, or local surveillance, and potentially inform infection prevention and control (IPC) efforts. Cost-effectiveness analyses are required to optimise sampling strategies.

## Introduction

The incidence of Gram-negative bloodstream infections (BSIs[GNBSIs]) has increased over the past decade, with widening geographic and socioeconomic inequality in patient outcomes.^1^ The increasing burden of antimicrobial resistance (AMR) in BSIs is driven by important Enterobacterales species, including *Escherichia coli*, *Klebsiella pneumoniae*, and *Klebsiella oxytoca*, comprising 85% of resistant bacteraemias in England in 2024/25.^1,2^ The availability of systematic pathogen genomic data, particularly if available routinely with linked clinical data, may help national^3^ and global^4–6^ efforts to mitigate the increasing incidence of GNBSIs and associated AMR, for example by identifying cryptic transmission chains or mechanisms contributing to increasing resistant BSI incidence. Tracking plasmids, often AMR vectors, may enable faster detection of plasmid-mediated outbreaks, and is increasingly feasible with Nanopore long-read bacterial genome assembly.^7,8^

In England, hospitals are mandated to report microbiological and patient-level data for *E. coli* and *Klebsiella* spp. BSIs for existing national surveillance programmes. There are however substantial financial, logistical, computational and analytical challenges associated with integrating whole genome sequencing (WGS) into national surveillance programmes, meaning that, to date, this has only been implemented for selected pathogens in a few high-income countries.^9^

It may be neither feasible nor necessary to perform WGS on every BSI isolate for public health surveillance. For example, sequencing all of approximately 42,000 *E. coli* BSIs in England in the 2024/25 financial year^2^ would cost ~£2.2M (at £52/isolate^10^). Additionally, it is estimated that 30% of the SARS-CoV-2 sequencing effort would have sufficed in some settings to detect emerging variants,^11^ while detection was delayed/missed elsewhere due to under-sampling.^12^ Sampling frames for GNBSIs have often been pragmatically selected, such as the first n,^13,14^ or every n^th^ isolate^15^, or selected by carriage of certain traits,^16,17^ rather than being informed by surveillance-specific calculations. Optimising sampling frames for genomic surveillance is therefore important for efficient public health resource allocation while meeting surveillance goals, including establishing background population structure and genomic epidemiology of various features, and detecting population-level transmission of novel bacterial lineages, plasmids and ARGs.^3,5,18^

There are currently two significant barriers to defining optimal sampling frames for genomic surveillance: i) a suitable dataset to inform calculations and ii) a suitable methodology. Existing longitudinal genomic surveys of BSIs are either historic, based on short-reads (therefore unable to accurately resolve plasmids), incomplete and/or geographically unrepresentative.^15–17^ Existing sample size formulae^19^ account for the presence/absence or prevalence of a single disease/feature, without accounting for the complexity of WGS data, including information about overall population structure and diversity. Furthermore, existing sample size formulae require assumptions on the minimum frequency of features to detect, without formal guidance on the choice of frequency threshold,^20^ or how this relates to proportion of population diversity captured.

To address these barriers, we i) developed the <u>N</u>ational *<u>E</u>. coli* and *<u>K</u>leb<u>S</u>iella* <u>U</u>K <u>S</u>urveillance Study (NEKSUS), which collected a 6-month nationally representative sample of BSIs from these species and long-read sequenced isolates, and ii) applied Bayesian analysis to this unique dataset to estimate the frequency distribution of genomic feature variants, explicitly allowing for unseen features, and therefore infer the sequencing sample size required to capture representative population diversity without requiring arbitrary frequency thresholds. We also propose the use of an ecological diversity measure, ‘coverage’ (or ‘sample/sampling coverage’ – i.e. proportion of the bacterial/plasmid/ARG population represented by unique features observed at least once in the sample) as an alternative to the *number* of unique features, to overcome issues related to estimating the potentially very large number of rare/singleton features, which may be less important for national surveillance. Here, we demonstrate the utility of this approach to estimate optimal sampling frames for routine genomics enhanced national surveillance, and provide a user-friendly R Shiny application, EpiSENTRY, allowing users to easily apply this method to their own data.

## Methods

### Literature review

Literature searches were conducted on surveillance sample size estimation (supplementary material; Table S1-S2).

### Isolate collection

Ten NHS microbiology laboratories, from hospitals with most emergency admissions, serving distinct geographic regions across all seven NHS England administrative areas were recruited to the NEKSUS Consortium. Consecutive, unselected *E. coli*/*Klebsiella* BSI isolates (routine clinical collection) were retrieved, October 2023-March 2024, deduplicating isolates of the same bacterial species/patient within 14-days.

### DNA extraction and sequencing

DNA was extracted mechanically (N=1,332, GENEWIZ, Leipzig, Germany), or enzymatically (N=860, MicrobesNG, Birmingham, UK). Sequencing libraries were prepared using Rapid Barcoding Kit_96V14 (Oxford Nanopore Technologies, Oxford, UK), and sequenced on a PromethION P2 Solo (R.10.4 flowcell; supplementary material).

### Bioinformatic analysis

POD5s were basecalled and demultiplexed using Dorado^21,22^ v5.0.0 (super-high accuracy). Long-read-only assembly/annotation was implemented in Nextflow^7,23,24^ v24.04.3.5916. Raw-reads were evaluated with SeqKit^25^ v2.9.0, subsampled 4-times to 60×,^8,26,27^ each assembled with five assemblers,^28–32^ combined with Autocycler^33,34^ v0.2.1, Medaka^7,35^ v2.0.1 polished, and quality-controlled using CheckM2.^36^ Assemblies were grouped using: 1) multi-locus sequence type (MLST)^37^ and 2) fastBAPS^38^ (alignment-based clustering). Plasmids were reconstructed and annotated (MOB-suite^39^ v3.1.9) and clustered (pling^40^ v2.0.0). ARGs were annotated using AMRFinderPlus^41^ v4.2.27. Recombination corrected trees were generated (Verticall^42^ v0.4.3; FAST-ME^43^ v2.1.6.3; supplementary material).

### Ecological Estimators of Diversity

Two ecological estimators, iNEXT^44^ and preseqR^45^ were used to estimate the number of unique *E. coli*/*Klebsiella* MLSTs (i.e. ‘species richness’) in the English BSI population. Species richness and three other diversity measures (Shannon diversity, Simpson diversity, and coverage) were investigated using rarefaction for stability to down-sampling and novel lineage incursion. Four methods (iNEXT,^44^ preseqR,^45^ rarefaction, and Bayesian) for estimating coverage (i.e. proportion of the bacterial population represented by features observed at least once) of *E. coli*/*Klebsiella* MLSTs were compared (supplementary material).

### Bayesian Estimation

Four Bayesian^46^ frameworks were used to estimate posterior frequency distributions: 1) Overall isolate-level features (MLSTs/fastBAPS); 2) Overall sub-isolate-level features (plasmid subcommunities/ARGs); 3) Regional isolate-level features; 4) Regional sub-isolate-level features (Figure S1).

#### 1) Overall isolate-level features

For isolate-level features (MLSTs/fastBAPS), where each sampling unit/isolate contributed exactly one categorical feature, we derived the posterior frequency distribution of all features (i.e. each ST/cluster), plus frequencies of ‘novel’/unseen categories. We assumed a prior Dirichlet distribution on feature frequency:

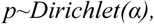

where *α* is a vector of ‘pseudo-counts’, one for each of *k* features (α_1_,α_2_,…,α_k_), plus ‘novel’ categories (*α_novel_1_*,*α_novel_2,_…,α_novel_*__*m*_). We assumed a symmetric uniform prior (*α*_1-*k*_ = *α_novel_1-novel_m_*=1). The number of ‘novel’ features, *m*, was estimates by empirical Bayes using the Chinese restaurant process^47^ (Table S3; supplementary material). We assumed a multinomial likelihood for observed feature counts, *y* (vector of length *k+m,*[*y*_1_,*y*_2_,…,*y_k_*, *y_novel_1_,…,y_novel_m_*], with 0 counts for novel features), over N isolates, yielding a posterior Dirichlet distribution on feature frequency:

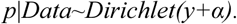

#### 2) Overall sub-isolate-level features

For sub-isolate-level features (plasmids/ARGs), where features are non-mutually-exclusive, a Beta-Binomial framework was used. This is analogous to 1), but with a binary (presence/absence) not categorical outcome, with separate prior distributions for each feature, *k*:

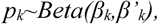

where *β_k_* is the prior pseudo-count of feature presence, and *β’_k_*, of absence. We assumed *β_k_=β’_k_,* although this is not required. The ‘novel’ features number was estimated from singleton frequency^48^, and each novel feature given the same prior as observed features (*β_k_=β’_k_=β_novel_1-novel_m_=β’_novel_1-novel_m_=*1. We assumed a Binomial likelihood for observed feature counts, *y*_1_,*y*_2_,…,*y_k_* (plus zero-counts for novel features), over N isolates, yielding a posterior Beta distribution on feature frequency:

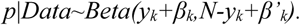

For 1-2) we sampled directly from the posterior.

#### 3) Regional isolate-level features

Region-specific frequencies for isolate-level features were modelled using a hierarchical extension of 1), involving multiple levels of prior (‘child’ distributions parameterised by ‘parents’). The likelihood for each region(*r*)-specific vector of feature counts, *y_r_*, was given by:

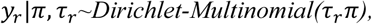

where *π* is a vector denoting global feature frequencies (length *k* plus ‘novel’ categories, determined as for 1)), and r_r_ is a region-specific concentration parameter. *π* was drawn from a ‘parent’ Dirichlet distribution parameterised by uniform prior vector, *α* (each element=1), and r_r_ was drawn from a ‘parent’ lognormal distribution. To allow regions to deviate from global frequencies by differing magnitudes, each r_r_was drawn from a different distribution, parameterised using ‘grand-parent’ distributions.

#### 4) Regional sub-isolate-level features

Region-specific frequencies for sub-isolate-level features were modelled using a hierarchical extension of 2), with each feature sampled from separate distributions. The likelihood of the observed count of each feature, *k*, in each region, *r*, *y_r,k_*, was given by:

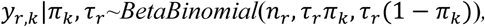

where *π*_k_ is the global frequency for feature *k*, including ‘novel’ categories (as for 2)), and r_r_ is a region-specific concentration parameter. *π*_k_ was draws from a ‘parent’ Beta distribution (with prior parameters *β_k_*,*β’_k_*), and a region-specific r_r_ was drawn as for 3). The effects of prior assumptions were investigated in sensitivity analyses (Figures S2-S6; Table S3-S4; supplementary material).

### Power calculation

Bayesian posterior frequency distributions were integrated and subtracted from 1, yielding an inverse cumulative frequency distribution (iCDF), showing the proportion of the population occurring at or above frequency, *f*. The iCDF was used to determine the minimum feature frequency, *f_min_*, capturing ≥80% of the population mass (i.e. ≥80% coverage). The numbers of unique features occurring ≥*f_min_* were expressed as percentages of features detected in NEKSUS. Frequency was used to link Bayesian posterior iCDFs (analogous to coverage) to sample size from power calculations,^19,49^ where the sample size, *n*, required to detect features of frequency ≥*f_min_*, with probability/certainty, *p*, is given by:

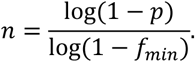

Sample sizes were expressed as percentages of the 42,224 *E. coli* and 13,078 *Klebsiella* BSIs occurring in England in the 2023/2024 financial year.^2,50^

### Implementation

Analyses were conducted in R^51^ v4.5.2. Standard Bayesian models were implemented using custom code, with posterior summaries (median; 95% credible interval [CrI]) from 10,000 draws (https://github.com/oxfordmmm/How_much_is_enough_for_genomic_surveillance_of_BSIs), and our method is available as an R Shiny application, EpiSENTRY (https://01a0c3dd-90d2-e597-58a5-51391a468387.share.connect.posit.cloud/). Hierarchical Bayesian models were implemented using CmdStanR^52,53^ v0.9.0, with four Markov chains, and 1,000 warm-up/1,000 sampling iterations each. Convergence was assessed using standard diagnostics (Table S4).

### Comparison to existing datasets

Bayesian estimation was applied to MLST data from two existing *E. coli* BSI datasets: NORM^16^ (2011-2017;n=1,997) and BSAC^15^ (2003-2012;n=1,326), covering periods of stable population structure (defined by original authors).

### Ethical approval

This work on de-identified NEKSUS sequence data was carried out as an extension of UKHSA national GNBSI surveillance following approval from the UKHSA Research Ethics & Governance Group (NR0429).

## Results

Of 1,781 *E. coli* and 582 *Klebsiella* BSI isolates collected within NEKSUS, 1,471 (83%) and 468 (80%) were assembled to sufficient quality (Figures S7-S8), carrying 3,361 and 847 plasmids, and 11,288 and 3,258 ARGs, respectively. These *E. coli* and *Klebsiella* isolates represented 263 and 295 unique MLSTs, 161 and 29 fastBAPS clusters, 825 and 417 plasmid subcommunities, and 189 and 199 unique ARGs, respectively (Table S5). The ten laboratories (‘regions’) individually contributed median 128 *E. coli* isolates (range:35-283) and 39 *Klebsiella* isolates (6-90; Table S5). The frequency distributions of genomic features were positively skewed nationally (Figure 1) and regionally, with many singleton/rare features (Figure S9-S10).

**Figure 1:**
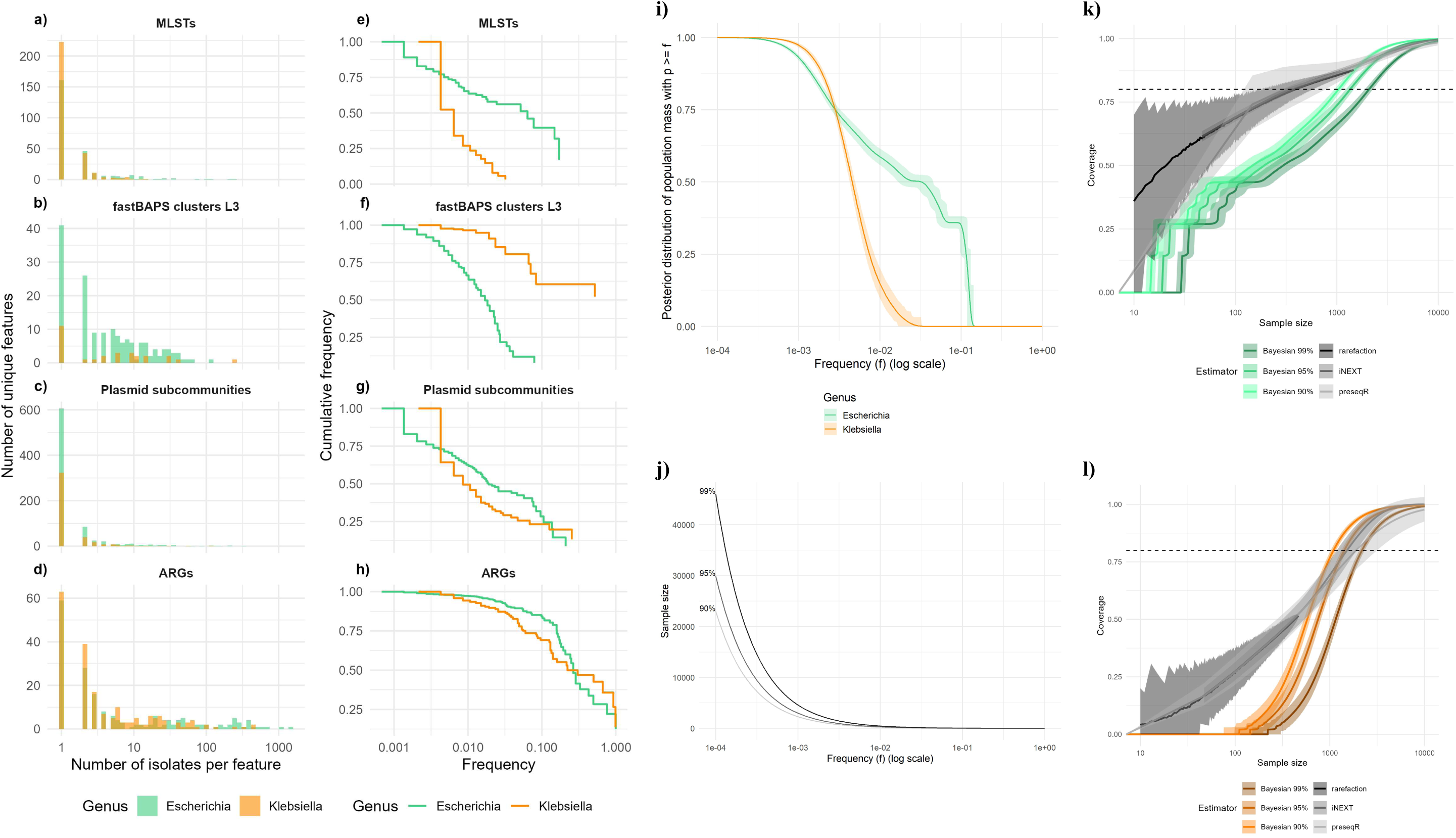
Frequency distributions of genomic features annotated in the NEKSUS dataset, shown as a-d) histogram of counts of each genetic feature, and e-h) cumulative population mass of features occurring at or above a given frequency. Features shown are, top to bottom rows: **a,e)** MLSTs, **b,f)** fastBAPS clusters, **c,g)** plasmid subcommunities, and **d,h)** AMR genes, for *E. coli* (green) and *Klebsiella* (orange). Note the x-axis for both left and right columns is on the log scale. The cumulative population mass is analogous to the inverse of (i.e: one minus) the cumulative distribution function (iCDF) of the frequency distribution (1-CDF), and shows the proportion of the population of *isolates* (for isolate-level features MLSTs and fastBAPS clusters), or the proportion of the population of *features* (for sub-isolate-level features plasmids and AMR genes) that have/are a feature that occurs *at or above* a certain frequency, ≥*f*. **i-l) Comparison of ecological estimators vs Bayesian modelling to estimate coverage at different sample sizes**. **i)** Median posterior probability estimates and 95% credible interval (CrI) obtained from 1,000 draws for the proportion of the population occurring at a frequency ≥*f*, for *E. coli* (green) and *Klebsiella* (orange). This plot is analogous to the top left plot of Figure 2b). **j)** Power calculations from minimum sample size needed to detect a feature (e.g. MLST) in a population occurring at frequency ≥*f*, with 90% (light grey), 95% (middle grey), and 99% (dark grey) certainty. **k,l)** Comparison of different methods of estimating the minimum sample sizes needed to achieve a given coverage for k) *E. coli* and l) *Klebsiella*. Coloured lines and curves show the median and 95% CrI, respectively, based on Bayesian modelling of the MLST frequency distributions in **i)** combined with the power calculations at 90%, 95%, or 99% certainty in **j)**, while the grey curves show estimates and 95% confidence intervals for standard ecological estimators. Sampling coverage can be interpreted as the proportion of the total population that would belong to MLSTs that occur frequently enough to be detected in a sample of a certain size with 90, 95, or 99% certainty. In comparison to Bayesian modelling, ecological estimators underestimate the sample size required at lower sampling coverages. Confidence intervals for ecological estimators are also very wide at higher coverages compared to Bayesian CrI because the latter exploit assumptions regarding the prior distribution whereas ecological estimators reflect uncertainty in unobserved distributions.

We first estimated the number of unique *E. coli*/*Klebsiella* MLSTs (i.e.: ‘species richness’) among English BSIs, using the two best-performing ecological estimators.^44,45,54^ We found that preseqR produced highly uncertain estimates, while iNEXT, produced considerably lower estimates, supporting previous evaluations suggesting iNEXT underestimates diversity (Figure S11; Table S6). Comparison with alternative diversity measures using rarefaction suggested that ‘coverage’ (i.e.: proportion of the population with features observed at least once in the sample) showed more favourable characteristics of plateauing/stabilising towards larger sample sizes, unlike species richness, while maintaining sensitivity to novel lineage incursion, unlike Shannon/Simpson diversity (Figures S12-13; supplementary material). Coverage was therefore used subsequently to quantify diversity. Comparison of four coverage estimators revealed that Bayesian analysis combined with power calculation showed lower uncertainty than ecological estimators, while yielding more conservative sample size estimates to capture 80% MLST coverage (Figure 1i-l; Table S6). The Bayesian estimation was implemented as an R Shiny application: EpiSENTRY – Epidemiological Sampling frame Estimation for geNomic surveillance of microbial diversiTY (https://01a0c3dd-90d2-e597-58a5-51391a468387.share.connect.posit.cloud/).

Bayesian analysis was used to estimate the posterior frequency distributions of genomic features (MLSTs, fastBAPS clusters, plasmid subcommunities, ARGs). Frequency distributions were integrated and subtracted from 1, yielding the cumulative population mass of bacteria with features occurring at/above frequency, ≥*f*. Frequency was used to link Bayesian posterior estimates of cumulative population mass (analogous to ‘coverage’) to power calculation sample size estimates (deterministic formulae to calculate the sample size, *n*, needed to detect features occurring at frequency ≥*f* with certainty, *p*; Tables 1; S6-S7). 80% coverage was selected as an arbitrary threshold, and the corresponding sample sizes (to observe at least one feature in the sample among those features comprising >80% of the bacterial population, at different levels of certainty), reported.

**Table 1:** *E. coli* and *Klebsiella* bloodstream infection sequencing sample size estimates derived from Bayesian analysis combined with power calculation to reach 80% coverage (i.e. to detect features representing 80% of the population) at 95% certainty, as well as the percentage of unique features and the minimum feature frequency detected at the estimated sample size.

| Estimator \ Coverage | Sample size to reach 80% coverage (95% credible interval) | Sample size as a percentage of annually-occurring BSIs* (95% credible interval) | Percentage** of unique features detected at 80% coverage (95% credible interval) | Minimum feature frequency at 80% coverage |
| --- | --- | --- | --- | --- |
| *E. coli* |  |  |  |  |
| MLST | 1,351 (1,230 - 1,472) | 3.2% (2.9-3.5%) | 27% (21-34%) | 0.0022 (0.0020-0.0024) |
| fastBAPS (level 3) | 649 (575 - 727) | 1.5% (1.4-1.7%) | 36% (29-43%) | 0.0046 (0.0041-0.0052) |
| Plasmid subcommunity | 1,928 (1,837 - 2,025) | 4.6% (4.4-4.8%) | 30% (27-34%) | 0.0016 (0.0015-0.0016) |
| ARG | 27 (23 - 30) | 0.06% (0.06-0.07%) | 11% (10-13%) | 0.1087 (0.0953-0.1267) |
| *Klebsiella* |  |  |  |  |
| MLST | 1,422 (1,331 - 1,515) | 10.9% (10.2-11.6%) | 49% (42-55%) | 0.0021 (0.0020-0.0022) |
| fastBAPS (level 3) | 113 (84 - 145) | 0.9% (0.6-1.1%) | 23% (13-37%) | 0.0260 (0.0204-0.0348) |
| Plasmid subcommunity | 1,115 (1,052 - 1,181) | 8.5% (8.0-9.0%) | 41% (36-45%) | 0.0027 (0.0025-0.0028) |
| ARG | 83 (72 - 95) | 0.6% (0.6-0.7%) | 16% (12-19%) | 0.0356 (0.0310-0.0410) |

\*Percentages calculated compared to the 42,224 *E. coli* and 13,078 *Klebsiella* BSIs occurring in England in the 2023/2024 financial year.<sup>50</sup>
\*\*As a percentage of the total number of unique features detected in a nationally-representative genomic survey of English BSIs (the NEKSUS Study).

Estimates varied by genomic feature, with more *E. coli*/*Klebsiella* isolates needed to reach 80% coverage with 95% certainty for MLSTs (1,351[95% credible interval (CrI):1,230-1,472] and 1,422[1,331-1,515], respectively) compared with fastBAPS clusters (649[575–727] and 113[84–145] respectively) and ARGs (27[23–30] and 83[72– 95], respectively; Tables 1; S6-S7; Figure 2). Higher sample sizes were required to reach 80% coverage for *E. coli* plasmids compared with other features (1,928[1,837–2,205]); however, for *Klebsiella*, sample sizes to reach 80% plasmid coverage (1,115[1,052–1,181]) were lower than for *Klebsiella* MLSTs. Sample size estimates increased with higher coverage, and certainty of detection (Tables 1; S6; Figure 2a-b). Although ~1,400 *E. coli*/*Klebsiella* isolates each would capture genomic features representing ≥80% of the estimated population of BSI lineage/ARG diversity (i.e. 80% coverage), percentages of unique features captured was lower. Specifically, these sample sizes captured features occurring at frequencies ≥0.22% and ≥0.21% for *E. coli* and *Klebsiella*, corresponding to 28% (95% CrI:24-31%) and 49% (44-52%) of unique MLSTs detected for each genus overall in the NEKSUS dataset, 59% (55-64%) and 87% (77-97%) of unique fastBAPS clusters, 19% (17-20%) and 50% (47-52%) of unique plasmid subcommunities, and 57% (52-62%) and 83% (78%-87%) of unique ARGs, respectively (Tables 1; S7-8).

**Figure 2:**
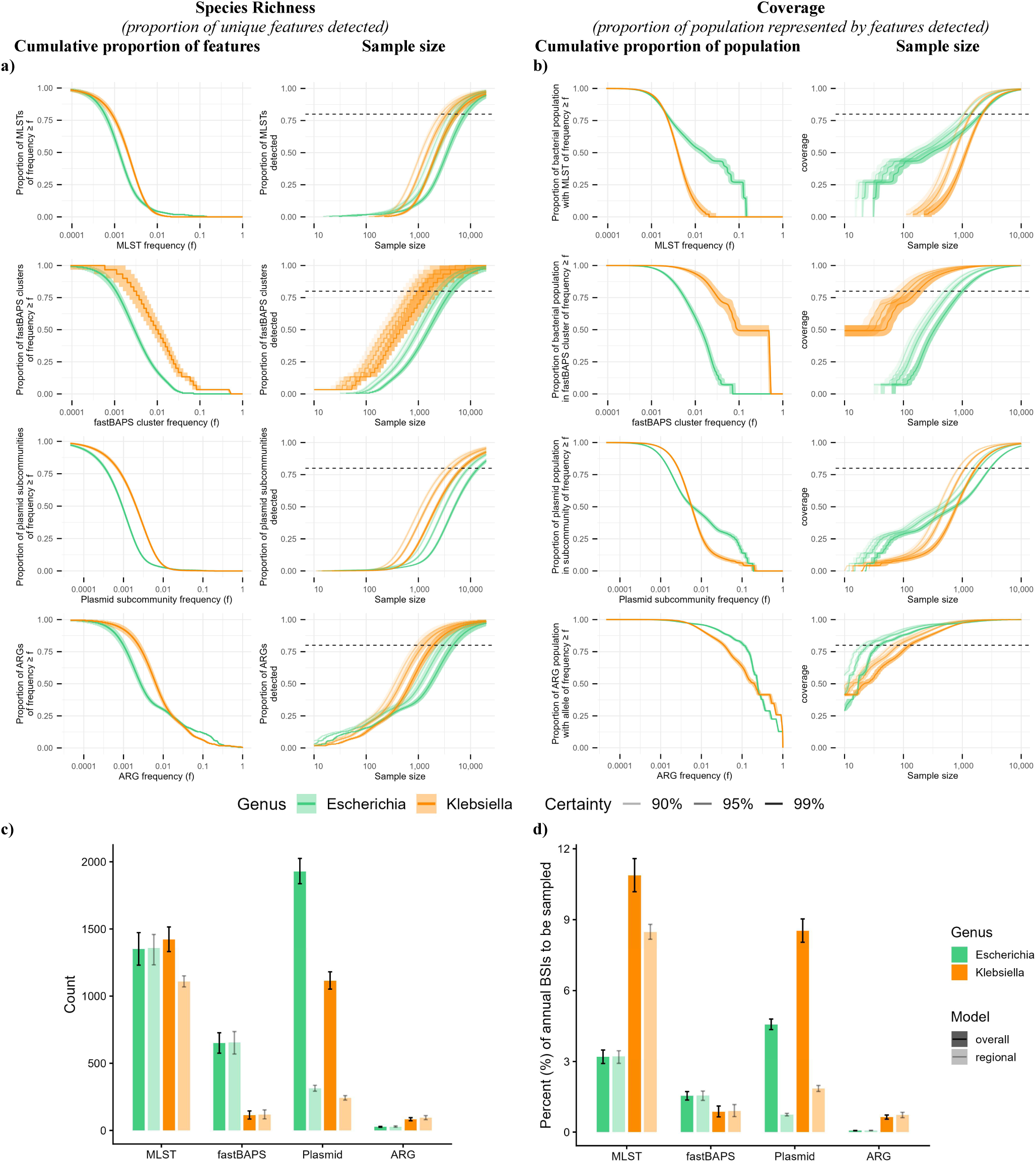
Posterior estimates of the frequency distributions of different genomic features obtained from Bayesian analysis of the NEKSUS dataset, and sample size estimates required to capture genomic diversity of different genomic features, measured as a) the number/proportion of unique features captured, and b-d) as the proportion of the total population represented by features observed in the sample (i.e. coverage). **a)** shows the cumulative proportion of unique features (y-axis, left column) that occur at a frequency ≥*f* (x-axis, left column), and the sample sizes (x-axis, right column) required to detect unique features (y-axis, right column) as a proportion of those detected in the NEKSUS study for *E. coli* (green) and *Klebsiella* (orange), at 90, 95 and 99% certainty. **b)** shows the cumulative population mass of genetic features (i.e. coverage; y-axis, left column) that occur at frequency ≥ *f* (x-axis, left column), and the minimum sample size (x-axis, right column) required to reach a certain coverage (y-axis; right column) for *E. coli* (green) and *Klebsiella* (orange), at 90%, 95%, and 99% certainty. Genomic features shown, from top to bottom, are MLSTs, fastBAPS clusters, plasmid pling subcommunities, and antimicrobial resistance genes (ARGs). Note that the ‘total population mass’ refers to all *isolates* for isolate-level features (MLSTs, fastBAPS clusters), while for sub-isolate-level features (plasmids, ARGs), this relates to the total population of *features* (i.e.: all plasmids or all ARGs). The difference between **a)** and **b)** is that in **a)**, all unique genomic feature categories contribute equally to the ‘proportion of unique features’, while in **b)**, common features contribute more to the ‘population mass’. Solid lines indicate median posterior estimates, and ribbons indicate 95% credible intervals from Bayesian estimates. The horizontal dashed line indicates 80% for both **a)** (80% of unique features detected) and **b)** (80% coverage). Note that the top left plot of d) is analogous to Figure 1i). **c-d)** Mean (+/− 95% credible interval [CrI]) Bayesian posterior estimates using NEKSUS data. **c)** and **d)** show the estimated samples sizes required for genomic surveillance to capture 80% of diversity at 95% certainty (i.e. where the dashed line from Figure 2b intersects the middle curves for each genus), **c)**, as absolute numbers, and **d)** as a percentage of the number of the total *E. coli*/ *Klebsiella* BSIs that occurred in England in the 2023/24 financial year (42,224 *E. coli* and 13,078 *Klebsiella*)^55^. Dark bars show estimates from the ‘overall’ models shown in **a-b)**, while translucent bars show national-level estimates from hierarchical models including regional information.

*Klebsiella* BSIs were more diverse than *E. coli*, relative to incidence in the 2023/24 financial year (42,224 *E. coli* and 13,078 *Klebsiella*),^55^ with sampling 10.9% (95% CrI:10.2-11.6%) vs 3.2% (2.9-3.5%) of total annual BSIs in England needed to reach 80% MLST coverage, respectively. fastBAPS clusters were an exception, with a lower percentage of *Klebsiella* required (0.9%[0.6-1.1%] vs 1.5%[1.4-1.7%] *E. coli*; Table 1; Figure 2d).

Incorporating regionally-stratified data did not substantially change overall national-level estimates except for *Klebsiella* MLSTs, and plasmids for both genera, where estimates incorporating regional-stratification were lower (Tables S9-10; Figure 2c-d; 3). However, these regionally-stratified estimates may be unreliable due to the high proportion of singleton plasmids/*Klebsiella* MLSTs, which resulted in many zero-count features per regions, reflected in poorer fit to overall observed feature frequencies (Table S3-S4; Figures S5-S6; supplementary material). Most regions were similarly representative of national-level diversity for *E. coli*/*Klebsiella* fastBAPS clusters and ARGs, with regional estimates similar to each other, and to overall estimates. Nevertheless, the magnitude of MLST diversity was regionally variable, particularly for *Klebsiella*, with several poorly-representative ‘outlier’ regions, where detected MLSTs represented <80% of the national population, even at large sample sizes (North West for *E. coli*; London and South East B for *Klebsiella*). These poorly-representative regions were those contributing fewest isolates to NEKSUS (86 *E. coli* from the North West; six *Klebsiella* from South East B; 28 *Klebsiella* from London; Table S5). Plasmids for both genera were region-specific, with individual regions capturing <50% and <30% coverage of national plasmid populations for *E. coli* and *Klebsiella*, respectively (Figure 3; Tables S9-10). Variation in proportions of unique NEKSUS features detected followed a similar pattern to coverage for *E. coli*, but with greater inter-regional variation for *Klebsiella* (Figure 3; Tables S11-12).

**Figure 3:**
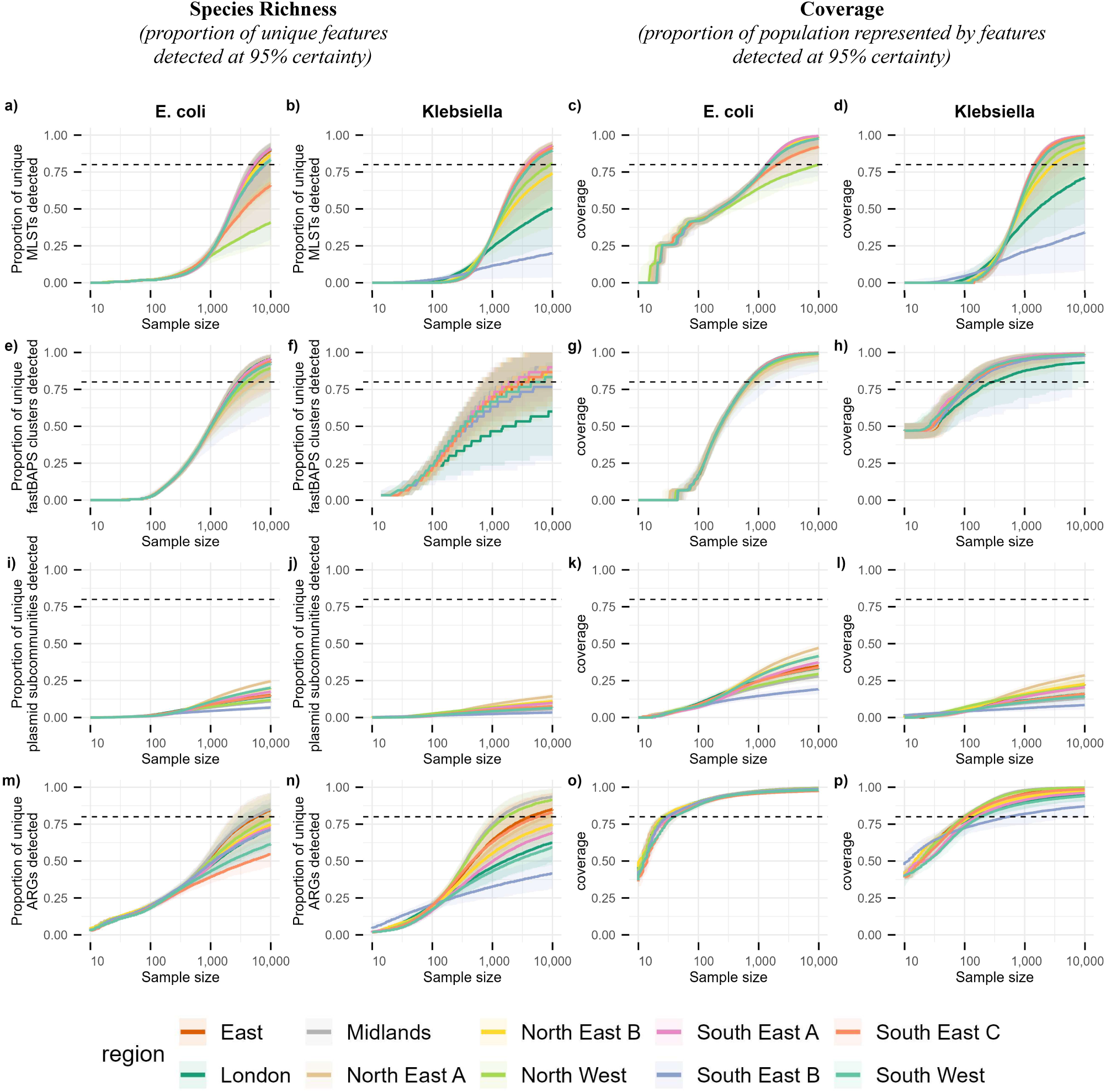
Regional estimates of sequencing sample sizes required to capture 80% of unique features (i.e. species richness; a-b, e-f, i-j, m-n), and features representing 80% of the national population (i.e. coverage; c-d, g-h, k-l, o-p), for *E. coli* (a,c,e,g,i,k,m,o), and *Klebsiella* (b,d,f,h,j,l,n,p) BSIs, for MLSTs (a-d), fastBAPS clusters (e-h), plasmids (i-l), and antimicrobial resistance genes (ARGs) (m-p), at 95% certainty. The horizontal dashed line indicates 80% diversity (80% unique features detected, or 80% coverage).

We also applied Bayesian analysis to external datasets of longitudinally-collected *E. coli* BSI isolates from Norway,^16^ and the UK.^15^ Estimated sample sizes to capture ≥80% MLSTs coverage with 95% certainty were more similar to previous UK data (median: 1,022[95% CrI:902-1,137]) than Norway (1,874[1,677-2,079]; Table S3;S6, Figure S14).

## Discussion

Genomic surveillance at scale promises to substantially enhance our understanding of BSI epidemiology, potentially supporting local/regional, and national mitigation efforts. In research settings, arbitrary decisions have been made about which and how many isolates are sequenced. As this technology is increasingly translated into routine clinical and public health settings, more principled decisions are needed about the design of optimal sampling frames to inform interventions and future research. Here, we demonstrated the utility of Bayesian analysis combined with power calculation as a scalable sample size estimation method for *E. coli* and *Klebsiella* BSI genomic surveillance, incorporating uncertainty and ‘unseen’ categories of genomic features, and available as an interactive R Shiny application, EpiSENTRY (https://01a0c3dd-90d2-e597-58a5-51391a468387.share.connect.posit.cloud/). We show that relatively modest sample sizes, 1,400 *E. coli*/*Klebsiella* isolates, represent ≥80% of the overall estimated population mass of BSIs. In contrast, rarer, but potentially clinically important genomic features, including ARGs and plasmids, remain undetected even with much higher sample sizes, reflecting the need to develop other enrichment methods for such features. We found that in general, there was greater diversity in *Klebsiella* spp. vs *E. coli*, and that diversity may be region-specific for *Klebsiella* MLSTs and plasmids of both genera. Importantly, our method can be generalised to other pathogen populations with available genomic data, as demonstrated using longitudinal UK and Norwegian surveys of *E. coli* BSI, to obtain comparable sample size estimates, potentially reflecting similar pathogen population structures.

Variation in sample sizes needed to achieve 80% coverage by genomic feature may be explained by feature-specific frequency distributions. The most conservative estimates were for plasmid subcommunities (~2,000 isolates), and most optimistic for AMR genes (<100 isolates). The appropriate coverage threshold may therefore depend on the genomic feature targeted by surveillance. For MLSTs, 80% coverage may be appropriate, corresponding to features occurring ≥0.22% and ≥0.21% frequency for *E. coli* and *Klebsiella*, respectively, with 95% certainty. Detecting rarer lineages may be less important for understanding national-level pathogen population structure, even if ≤50% of unique MLSTs remain undetected. Nevertheless, rarer bacterial lineages may be involved in local outbreaks, and rare ARGs (e.g: carbapenemase-encoding genes), may be of high public health importance nationally, necessitating alternative diversity measures, or targeted sequencing based on epidemiological/phenotypic metadata in these settings.

The greater diversity in *Klebsiella* MLSTs has been previously documented,^17^ and is confirmed here by *Klebsiella* requiring a greater percentage of annually-occurring English BSIs to be sampled at any given coverage threshold compared with *E. coli*. Compared with the NEKSUS dataset size, the estimated sample size to capture 80% *E. coli* MLST coverage was similar, while the *Klebsiella* estimate was >3x higher, suggesting the 10-laboratory NEKSUS consortium may be appropriate for *E. coli* surveillance, while for *Klebsiella*, the surveillance network or sampling period requires extension.

While poorly-representative ‘outlier’ regions for MLST diversity are likely related to biases from small numbers of isolates submitted to NEKSUS from these regions, all regions showed region-specific plasmid populations, where sampling a single region would capture <50% of national diversity. These results emphasise the importance of regionally-representative sampling to generate data to support local/regional and national surveillance.

Study strengths include scalability and adaptability. Being a frequency-based method, Bayesian analysis is independent of population size, therefore not subject to the same challenges as ecological estimators^44,48,54,56–58^ when extrapolating to large populations. Accounting for ‘unseen’ features in sample size estimation may also help account for incomplete sampling in the survey informing estimates. Furthermore, using coverage (proportion of the population represented by features detected in the sample) as a diversity measure may be advantageous compared with ‘species richness’ (total number of unique features), by reducing uncertainty where the number of unique features is unknown or infinite due to mutation/sequencing errors, and placing more weight on common features that are of greater public health importance. Coverage may also be informative for other applications, such as estimating bacterial surface antigen diversity for vaccine design and monitoring, where it may directly correlate to vaccine coverage. A further strength is the large nationally-representative long-read genomic dataset of 2000 BSI isolates (NEKSUS). The unselected sampling frame, including both resistant and susceptible isolates, enables unbiased estimation of background genomic diversity among English BSIs. The regional stratification, and use of long-read sequencing, were both crucial in elucidating inter-regional differences in plasmid diversity, which may not have been possible with short-reads.

Limitations include the requirement for a context-specific, representative genomic survey on which to base estimates, which may be a barrier in under-studied settings such as low-middle income countries, which may have different clinical *E. coli* population structures compared with the UK/Norway. Furthermore, the validity of estimates is dependent on assembly/annotation accuracy. While NEKSUS bioinformatic workflows aimed to maximise accuracy with consensus-grade assembly and validated annotation tools, residual errors will propagate to sample size estimates. The choice of ‘level’ of genomic diversity also influences estimates. Broad lineage-level features (MLSTs/fastBAPS clusters) may conceal functionally and epidemiologically important diversity, such as *E. coli* ST131 clades with distinct adhesins and resistance associations.^59^ Conversely, overly-narrow labels (e.g.: strict plasmid clustering thresholds) may produce prohibitively high sample size estimates for national surveillance, and may be more appropriately applied to selected high-risk local settings. Discrete labels fail to account for continuity in relatedness, a particular challenge for capturing complex plasmid dynamics. Estimates for very low frequencies, corresponding to <1 NEKSUS isolate may be unreliable.

In conclusion, Bayesian estimation combined with power calculation may be a feasible method for estimating sample sizes for cost-effective genomic pathogen surveillance for public health, noting that attrition factors should be applied to account for data loss between isolate collection and quality-controlled genomic assemblies (83% and 80% of *E. coli* and *Klebsiella* isolates collected here). It may also hold potential applications in supporting therapeutics development, e.g.: vaccine target antigens. Incorporating serial sampling,^49^ and investigating further surveillance endpoints, including time-to-detection of clusters, would also be informative for public health policy. However, estimates must be tailored to context-specific data and surveillance goals.

## Data availability

Reads and assemblies have been uploaded to the European Nucleotide Archive (PRJEB93885; PRJEB108305). The Nextflow assembly/annotation pipeline (https://github.com/oxfordmmm/autocycler_ont_long_read_assembly_pipeline), and R analysis code (https://github.com/oxfordmmm/How_much_is_enough_for_genomic_surveillance_of_BSIs) is available on GitHub. Sample accessions and intermediate genomic data files are available on FigShare (https://doi.org/10.6084/m9.figshare.32326584). The EpiSENTRY sample size estimator tool is available as a Posit Cloud Connect R Shiny application (https://01a0c3dd-90d2-e597-58a5-51391a468387.share.connect.posit.cloud/).

## Funding

This research was supported by the National Institute for Health and Social Care Research (NIHR) Health Protection Research Unit in Healthcare Associated Infections and Antimicrobial Resistance (NIHR207397), a partnership between the UK Health Security Agency (UKHSA) and the University of Oxford. This work was also supported by the UKHSA and the NIHR Oxford Biomedical Research Centre (BRC) and the UKHSA PhD Funding Competition. DJW was supported by a Robertson Fellowship. ASW is an NIHR Senior Investigator. The cloud compute infrastructure for this work was donated by Oracle Corporation Infrastructure. The views expressed are those of the authors and not necessarily those of the NIHR, UKHSA or the Department of Health and Social Care. The authors were not precluded from accessing data in the study, and all authors accept responsibility to submit for publication.

## Environmental Impact Statement

The Nextflow pipeline for assembly and annotation of the ~2000 NEKSUS Enterobacterial genomes was run on Oracle cloud VM, and took 4224 GPUh on NVIDI Tesla V100 GPUs, each with 16GB RAM, drawing 2.02 MWh (Dorado super-high accuracy basecalling), and 8800 CPUh on an AMD EPYC 9J14 96-Core Processor with 1Tb RAM, drawing 109.29 kWh (assembly/annotation). Bayesian analysis including sensitivity analysis was run on a Dell Latitude 7430 desktop computer with a 12 i7-1270P Inter® Cores, taking 768CPUh and drawing 59.78 kWh. Running on the UK electricity grid, this has a carbon footprint of 505.04 kgCO2e, equivalent to 45.91 tree-years, and 2883 km in a passenger car (calculated using green-algorithms.org v2.2^60^). The carbon cost of pipeline/code development and that of wet lab isolate processing and transportation is not included in this calculation and is therefore likely to be an underestimate.

## Conflicts of Interest

The authors have no conflicts of interest to declare.

## Author Contributions

SH, DC, ASW, JR, KLH, AL, DJW, DW, RH, CSB, NS, and SL were involved in conceptualisation, funding acquisition, project administration, provision of resources and supervision. NEKSUS consortium members were involved in isolate collection and processing. Methodological development and validation of bioinformatic and statistical methods and software was done by DN under the supervision of SL and NS. DN, SL and NS were involved with data curation, analysis, investigation, visualisation and writing/editing. All authors approved the final draft.

## Acknowledgements

The authors would like to acknowledge all participating laboratories in the NEKSUS consortium who were responsible for isolate collection, Zeynab Yusuf from UKHSA for her role in sample transportation from UKHSA to Oxford, laboratory and bioinformatician colleagues at the Modernising Medical Microbiology Unit at the University of Oxford for support in methodological development and execution, as well as GENEWIZ Germany GmbH (Leipzig, Germany) and MicrobesNG (Birmingham, United Kingdom) for performing long-read sequencing.

Individuals within the NEKSUS consortium group authorship are (listed alphabetically):

- Alan McNally (University Hospitals Birmingham NHS Foundation Trust)
- Arthur Clegg (Barts Heath NHS Trust)
- Caroline Cullerton (The Newcastle-upon-Tyne Hospitals NHS Foundation Trust)
- Christopher R. Jones (UKHSA)
- Conor Bowman (Barts Heath NHS Trust)
- Gabriella Shanks (Barts Heath NHS Trust)
- Gillian Rodger (UKHSA)
- James Price (University Hospital Sussex NHS Foundation Trust)
- Jasvir Nahl (Leeds Teaching Hospitals NHS Trust)
- Jenny Bradbury (UKHSA)
- Jonathan Lambourne (Barts Health NHS Trust)
- Julie Samuel (The Newcastle-upon-Tyne Hospitals NHS Foundation Trust)
- Jumoke Sule (UKHSA/ Cambridge University Hospitals NHS Foundation Trust)
- Ian Butler (Barts Health NHS Trust)
- Katie Hopkins (University of Oxford)
- Katie Jeffrey (Oxford University Hospitals NHS Foundation Trust)
- Kavita Sethi (Leeds Teaching Hospitals NHS Trust)
- Lucinda Barrett (Oxford University Hospitals NHS Foundation Trust)
- Mark Garvey (University Hospitals Birmingham NHS Foundation Trust)
- Martin Williams (University Hospitals Bristol and Weston NHS Foundation Trust)
- Nicholas Brown (Cambridge University Hospitals NHS Foundation Trust)
- Nicola Childs (North Bristol NHS Trust)
- Paul Randell (University Hospital Sussex NHS Foundation Trust)
- Poorvi Patel (Cambridge University Hospitals NHS Foundation Trust)
- Robert Morran (University of Birmingham)
- Samuel Stafford (North Bristol NHS Trust)
- Samuel Tetley (University Hospital Sussex NHS Foundation Trust)
- Sarah Oakley (Oxford University Hospitals NHS Foundation Trust)
- Simon Eccles (Manchester University Hospitals NHS Foundation Trust)
- Valentina Pennetta (University of Oxford)
- Uchenna Ume (Barts Heath NHS Trust)

## Notes

### Competing Interest Statement

The authors have declared no competing interest.

### Author Declarations

The Research Ethics & Governance Group of the United Kingdom Health Security Agency (UKHSA) waived ethical approval for this work (NR0429) as it was carried out as an extension of UKHSA national surveillance of Gram-negative bloodstream infections (which is required/allowed by national legislation). All work was done on de-identified bacterial sequence data.

### Summary of Updates

Updated links to R Shiny app for EpiSENTRY sample size estimation tool

