## supplementary material for "How much is enough? Optimising sampling frames for genomic surveillance of *Escherichia coli* and *Klebsiella* spp. bloodstream infections – a retrospective study"

### Supplementary Methods

#### 1. Literature review

A rapid systematic literature review was conducted on MEDLINE (via Ovid), 13-14/05/2026, to evaluate existing evidence on sample size estimation methods for public health surveillance (search concepts: sample size estimation/calculation AND surveillance; Table S1-S2). No exclusions based on publication date, language, and publication type were applied. Inclusion criteria were records reporting sample size estimation or calculations methods in a public health setting. Exclusion criteria were studies simply reporting the use of an established sample size estimation method. Title/abstract and full-text screening was conducted by a single reviewer (DN).

#### 2. Isolate processing, DNA extraction and sequencing

Isolates were stored in brain–heart infusion broth with 10% glycerol at −70°C and then grown on blood agar for 24h at 37°C, following which a sweep of the pure bacterial culture was suspended in 1ml phosphate buffer saline, pelleted and cold-packed (N=1,332, GENEWIZ) or suspended in 1ml DNA/RNA Shield (Zymo Research, USA) and shipped at ambient temperature (N=860, MicrobesNG). Sentence cut form methods: Bacteria were subcultured for a further 24h at 37°C where there was insufficient 24h growth.

For GENEWIZ-processed isolates (N=1,332), DNA was extracted using the MagMAX Microbiome Ultra Nucleic Acid Isolation Kit with bead plate (Life Technologies, Carlsbad, CA, USA), quantified using the Qubit 4.0 Fluorometer and qualified using the Agilent 5600 Fragment Analyzer. For MicrobesNG-processed isolates (N=860), cells were lysed using TE buffer containing lysozyme (MPBio, USA), metapolyzyme (Sigma-Aldrich, USA) and RNase A (ITW Reagents, Spain), followed by treatment with proteinase K (VWR Chemicals, Ohio, USA) and SDS (Sigma-Aldrich, Missouri, USA). Genomic DNA was purified using SPRI beads, resuspended in EB buffer (10 mM Tris-HCl, pH 8.0), and quantified with the Quant-iT dsDNA HS (ThermoFisher Scientific) assay in an Eppendorf AF2200 plate reader (Eppendorf UK Ltd, United Kingdom) and diluted as appropriate.

For both sequencing providers, DNA libraries were prepared for sequencing using Rapid Barcoding Kit 96 V14 (Oxford Nanopore Technologies, Oxford, UK) according to manufacturer's recommendations. Barcoded samples were pooled (96-plexed) before solid-phase reversible immobilisation-cleaning and addition of Rapid Adapters to the tagged ends. The library pools were loaded onto ONT PromethION flow cells (R10[M Version]) – one 96-plex pool per flow cell – and sequenced on a PromethION P2 Solo for 72h (GENEWIZ), or were loaded onto a FLO-PRO114M (R.10.4.1) flow cell and sequenced according to manufacturer's instructions (MicrobesNG).

#### 3. Bioinformatic analysis

Computational analysis was performed on a virtual machine in the Oracle Cloud Infrastructure. POD5 files were basecalled and demultiplexed using Dorado[1,2] v5.0.0 (super-high accuracy 5mCG, 5hmCG and 6mA methylation aware simplex DNA model). Long-read-only assembly and annotation was implemented in Nextflow[3] v24.04.3.5916, using a pipeline[4] previously shown to produce Enterobacterales assemblies of superior completeness and comparable accuracy to hybrid assembly (i.e. assemblies combining nanopore long-read and Illumina short-read data).[5] All bioinformatic tools were run using default settings unless specified. Raw-read quality was evaluated with SeqKit[6] v2.9.0. Long-reads were subsampled to generate four independent sets of 60× reads using subsampling and genome size estimation scripts from Autocycler[7,8] v0.2.1. Genomes were assembled with using Autocycler[7,8] v0.2.1 using 20 input assemblies (4x each of Canu[9] v2.2, Flye[10] v2.9.5, Raven[11] v1.8.3, Miniasm[12] v0.3 and Plassembler[13]). A single 60× subsampling depth was used based on previous benchmarking[14-16] showing Flye performance was comparable at 60–100× read depth and a 60x depth minimised compute. Assemblies were polished using Medaka[17] v2.0.1 (--bacteria flag) using un-subsampled long-reads.[5] Assembly quality was assessed for completeness (>99%; longest contig at least 4Mb) and contamination (<5%) using CheckM2.[18]

Polished assemblies passing quality control were grouped using two methods: 1) multi-locus sequence type (MLST; a 7-locus lineage assignation) and 2) fastBAPS clusters (an alignment-based rapid clustering approach). MLSTs, and clonal complex/clonal group where appropriate, were annotated using Kleborate[19] v3.1.2. fastBAPS[20] v1.0.8 clusters were generated by annotating assemblies with Bakta[21] v1.10.4, aligning core genomes using Panaroo[22] v1.6.0 (95% presence threshold defining 'core') and extracting variable sites using snp-sites[23] v2.5.1. Separate core genome alignments were generated for each species complex (*E. coli*, *Klebsiella pneumoniae*, *K. oxytoca*, *K. aerogenes*). fastBAPS clustering was run using symmetric optimisation of hyperparameters, with the third level of clustering (level 3) selected as this gave the most appropriate lineage resolution by partitioning large MLSTs (e.g. *E. coli* ST131) while minimising singletons. Plasmids were reconstructed and annotated using MOB-suite[24] v3.1.9 (mob_recon and mob_typer). Plasmids were clustered into communities and subcommunities with pling[25] v2.0.0 default containment and DCJ-Indel thresholds (0.5, and ≤4, respectively). ARGs were annotated using AMRFinderPlus[26] v4.2.27, using --species flags assigned by Kraken2[27] v2.1.3 (Table S5; Figure S7-10).

Recombination corrected distance matrices were generated for all *E.coli* and all *Klebsiella* spp. BSI isolates using Verticall v0.4.3[28] (distance tree workflow), and Genus-specific phylogenetic trees built using FAST-ME v2.1.6.3[29] (Figure S10).

#### 4. Comparison to ecological estimators of diversity

##### a. Best performing ecological estimators applied to the NEKSUS dataset

The two best-performing ecological diversity estimator tools, iNEXT[30] and preseqR,[31] identified from a comprehensive review and evaluation of ecological estimators of diversity by Schmitz et al.,[32] were applied to BSI-associated *E. coli* MLST data from the NEKSUS study. 'Up-sampling' (extrapolation) was applied to population size of up to 42,224 for *E. coli* and 13,078 for *Klebsiella* spp., corresponding to the number of BSIs occurring in England in the 2023/24 financial year[33] for the respective genera. These tools were implemented in R, following the online documentation of these tools. Bespoke code for the investigation of ecological estimators and all other below analyses is available online (https://github.com/oxfordmmm/How_much_is_enough_for_genomic_surveillance_of_BSIs; Figure S11).

##### b. Rarefaction

To produce rarefaction curves (species/feature accumulation curves), sampling both *without* (more intuitive results) and *with* (simulates an infinite population) replacement was conducted using bespoke R code, on *E. coli* and *Klebsiella* MLST, AMR gene, and plasmid pling subcommunity data from the NEKSUS dataset. For each integer sample size between 1 isolate and the maximum number of isolates collected in the NEKSUS dataset for each genus (1,471 for *Escherichia*, and 468 for *Klebsiella*), 1000 random samples were taken, and four metrics were calculated from each draw: 1) Simpson diversity, 2) Shannon diversity, 3) species richness (i.e. the total number of unique MLSTs observed in the sample),[34] and 4) coverage (i.e. the proportion of the population represented by MLSTs observed in the sample, as estimated by Good's coverage[35,36]). The metrics were summarised across the 1000 draws with mean and 95% confidence intervals (CI; Figure S12).

##### c. Incursion of a novel species

To assess the sensitivity of each diversity metric (Shannon diversity, Simpson diversity, species richness and coverage) to detecting a novel species/feature (e.g. the emergence of a novel bacterial lineage), incursion was simulated by replacing the specific MLST label of randomly selected *E. coli* and *Klebsiella* isolates from the NEKSUS dataset with a 'novel' label. This simulated dataset was then randomly sampled at different sample sizes, without replacement, up to the actual NEKSUS sample size (1,471 for *E. coli* and 468 for *Klebsiella*). Each simulation was repeated 100 times and mean and 95% credible interval (CrI) summary metrics for each of the four metrics was calculated. The random replacement was repeated 100 times at each number of isolates to be replaced by the 'novel' label. This simulation was implemented using bespoke R code (Figure S13).

**d. Comparison to sample size estimation**

We compared sample size estimates obtained from Bayesian modelling to those obtained from ecological estimators as follows: First, the posterior frequency distribution of *E. coli* and *Klebsiella* MLSTs was estimated using Bayesian modelling. These posterior frequencies were then integrated to produce a cumulative population mass curve and subtracted from 1, to estimate the proportion of the population possessing a feature occurring at a frequency $\geq f$ (analogous to the inverse of the cumulative distribution function [iCDF]). A power calculation was separately performed to obtain the sample size, *n*, required to detect a feature occurring at frequency $\geq f$. These two estimates were combined, using the common 'frequency' axes, to obtain the proportion of the population mass captured (i.e. coverage) at various sample sizes. Coverage estimates from rarefaction, and the tools iNEXT[30] and preseqR,[31] were presented on the same axes to compare the absolute sample size estimates obtained by the different methods (Figure 1; Table S5).

### 5. Bayesian modelling frameworks

The four Bayesian[37] frameworks used to estimate posterior frequency distributions of genomic features are illustrated in Figure S1. Details not covered in the main text are described in detailed below, for: 1) Overall isolate-level features (MLSTs, fastBAPS clusters); 2) Overall sub-isolate-level features (plasmid subcommunities, ARGs); 3) Regional isolate-level features; 4) Regional sub-isolate-level features.

***1) Overall isolate-level features***

The number of 'novel' features (unseen in the NEKSUS dataset), *m*, was derived using the Chinese restaurant process (CRP).[38] In the CRP, 'guests' arrive at a restaurant (analogous to sampling isolates) and take a seat at either at a new table (analogous to sampling a novel feature), or a table that already has guests on it, with a probability proportional to the number of guests already seated at that table (analogous to sampling a feature that has already been observed). There are a potentially infinite number of tables, each with infinite capacity. The 'novelty hungriness' of a CRP-like system can be summarised using a single parameter, $\theta$, and the probability, $u$, of the next guest (or isolate) falling into a novel category is given by $u = \frac{\theta}{N+\theta}$, where N is the total NEKSUS sample for that bacterial genus. As *u* is expected to decay with increased sampling, this was used as an upper estimate to inform the prior number of novel features. $\theta$ was estimated from NEKSUS data using a Gamma prior:

$$\theta \sim Gamma(1, 1),$$

and likelihood:

$$p(counts\ |\theta) \propto \theta^k \frac{\Gamma(\theta)}{\Gamma(\theta+N)},$$

where k is the number of unique feature categories (i.e.: MLSTs or fastBAPS clusters). This gave the posterior:

$$p(\theta|\ counts) \propto \theta^k\ e^{-\theta} \frac{\Gamma(\theta)}{\Gamma(\theta+N)}.$$

We sampled the posterior directly with 5000 draws, summarised the median and 95% CrI for $\theta$, and calculated the corresponding probability of a novel feature, $u$, form the posterior median $\theta$ for each genomic feature/genus (Table S3). All observed and 'novel' features were given the same prior 'pseudo-count' in the prior parameter vector, $\alpha$. Therefore, to ensure the prior mass (i.e. probability) of all novel features collectively equalled the novelty probability, $u$, this required the number of novel features, *m*, for frameworks 1 and 3) to be given by:

$$m = \frac{uk}{1-u}.$$

***2) Overall sub-isolate-level features***

For sub-isolate-level features (plasmids/ARGs), the CRP could not be applied as features are non-mutually-exclusive. Instead, the prior number of novel features was estimated by the Good-Turing estimator,[36,39] which used the frequency of singletons, $y_k=1$ (i.e: features occurring only once in the sample data), out of a sample of N, to approximate the probability, $u$, of discovering new features:

$$u = \frac{\sum(y_k = 1)}{N}.$$

As for frameworks 1) and 3), all observed and 'novel' features were given the same prior distribution (both for $\beta$ and $\beta'$). Therefore, to ensure the prior mass (i.e. probability) of all novel features collectively equalled the novelty probability, $u$, this required the number of novel features, $m$, for frameworks 2) and 4) to be given by:

$$m = \frac{uk}{1-u}.$$

***3) Regional isolate-level features and 4) Regional sub-isolate-level features***

The number of prior novel features were determined as for 1) and 2), respectively. The region-specific concentration parameter, $\tau_r$, was sampled from a lognormal parent distribution:

$$\tau_r \sim Lognormal(\mu_{log\tau}, \sigma_{log\tau}),$$

whose parameters were sampled from Normal and exponential distributions:

$$\mu_{log\tau} \sim N(log(\mu_{\mu log\,\tau}), \sigma_{\mu log\,\tau}) \text{ and } \sigma_{log\,\tau} \sim Exp(\lambda_{\sigma log\,\tau}).$$

Hyperparameter values are given in Table S4.

**6. Sensitivity analysis and assessment of goodness of fit**

The impact of prior assumptions on Bayesian posterior estimates was assessed. For 'standard' Bayesian modelling (without regional stratification), the impact of varying the value of three parameters was explored: i) the prior pseudo-count on each isolate-level feature, $\alpha_k$, or prior pseudo-count of success/presence of each sub-isolate-level feature $\beta_k$; ii) the number of novel features, $m$, and iii) the total mass of novel features. All the prior Beta distributions in framework 2) were assumed symmetrical, with the two parameters equal to each other, $\beta_k = \beta'_k$. For standard Bayesian modelling, the prior pseudo-count of observed features was allowed to deviate from that of novel features, to enable the comparison of including e.g.: 100 uncommon novel features, vs 1 novel feature at 100x the frequency. Default parameters, and parameter ranges explored are shown in Table S3. Goodness of fit was measured using log10-root mean squared error (log10-RMSE) by comparing median posterior estimates to actual observed feature frequencies (overall in the NEKSUS dataset), and observed feature frequencies plotted against Bayesian posterior predictions (Figure S2-3).

For hierarchical Bayesian modelling (i.e.: models including regional stratification) of isolate-level features, the impact of varying five parameters was explored: i) the prior pseudo-count on each isolate-level feature, $\alpha_k$; ii) the number of novel features; iii-v) the three hyperparameters defining the distribution of, $\tau_r$, the region-specific concentration parameter, $\mu_{\mu log\,\tau}$, $\sigma_{\mu log\,\tau}$, and $\lambda_{\sigma log\,\tau}$. Uniform priors were used where the prior pseudo-counts of each novel feature were kept equal to that of observed features (therefore the novel mass was equal to the number of novel features for $\alpha_k$=1). For hierarchical Bayesian modelling of sub-isolate-level features, the impact of varying six parameters was explored: i) the prior pseudo-count of success/ presence of each sub-isolate-level feature $\beta_k$; ii) the number of novel features; iii-v) the tree hyperparameters of the $\tau_r$ distribution, as for isolate-level features detailed above; vi) the second prior Beta distribution parameter, $\beta'_k$, reflecting the prior frequency of failure/absence of a feature. As for isolate-level features, the same prior distribution was used for observed and novel features. Default parameters and prior parameter ranges explored in hierarchical models are shown in Table S4.

For all hierarchical Bayesian models, predictive performance was assessed using a held-out log predictive density, where a regionally-stratified, randomly-selected 2/3 of the NEKSUS dataset were used for model fitting (training-set), and the remaining 1/3 was used for assessing predictive performance (test-set). Held-out log predictive density was computed, for each posterior draw, using the Dirichlet-multinomial likelihood, $\log p\,(y|\alpha)$, for isolate-level features, and Beta-binomial likelihood, $\log p\,(x|n,\alpha,\beta)$, for sub-isolate-level features of the held-out (test-set) data (i.e. the log likelihood of the observed counts in the test-set data given the model fitted to the training-set; Figure S4).

The Dirichlet-Multinomial log-likelihood was given by:

$$\log p\,(y|\alpha) = \log\left(\frac{n!}{\prod y_k!}\right) + \log\left(\frac{\Gamma(\sum_k \alpha_k)}{\Gamma(\sum_k \alpha_k + n)}\right) + \sum_k [\log\Gamma(y_k + \alpha_k) - \log\Gamma(\alpha_k)],$$

and the Beta-Binomial likelihood was given by the closed-form marginal likelihood:

$$\log p\,(x|n,\alpha,\beta) = \log\binom{n}{x} + \log Beta(x + \alpha, n - x + \beta) - \log Beta(\alpha,\beta)\,.$$

Likelihoods were summed across regions for Dirichlet-Multinomial likelihoods, and for features and regions for Beta-Binomial likelihoods, and then mean averaged over posterior draws. For numerical stability, the mean was calculated on the exponentiated log likelihoods (on the probability scale), and results transformed back to the log scale ($\log(mean(e^{likelihood}))$). Scores were normalised per isolate.

Sampling was run with 1 Markov chain with 250 warm-up and 250 sampling iterations for sensitivity analyses for computational efficiency. The thresholds used to indicate acceptable chain convergence for hierarchical Bayesian models in both main estimation and sensitivity analyses (Table S4) were: divergent transitions and transitions hitting maximum tree depth <5% of sampling iteration; the third quantile of $\hat{R}$ <1.05, and lower quantile of effective sample size >100. Fit of posterior estimates for hierarchical Bayesian models to observed NEKSUS data with default parameters was visualised using scatterplots (Figure S5-6).

### Supplementary Results

#### 1. Literature review

A systematic search of MEDLINE (via Ovid) for the search concepts "sample size estimation/calculation" AND "surveillance" (Table S1), with no restrictions on publication type, date or language retrieved 62 records, of which 22 were eligible after title/abstract and full-text screening. Inclusion criteria included being a methodological study on sample size estimation, while studies applying established sample size calculations were excluded. Over two-thirds (68% [15/22]) of studies reported surveillance sample size calculation based on conventional frequentist sample size formulae, while 36% (8/22) reported simulation/regression modelling-based methods. The most common surveillance outcomes were prevalence/incidence (59% [13/22]) and presence/absence (54% [12/22]) of a disease/trait while all studies considered diseases/traits individually for calculations. Aspects of sample size estimation that were focused on were complex/non-random sampling (72% [16/22]), sampling over time (27% [6/22]), imperfect diagnostic test performance (18% [4/22]), non-random disease distribution (14% [3/22]), and missing data (14% [3/22]. Nevertheless, no studies provided formal guidance on selecting the minimum frequency threshold for diseases/traits required for classical sample size calculations,[40] or how calculations could be extended to surveying multiple traits/features simultaneously (Table S2).

#### 2. Comparison to ecological estimators of diversity

We first attempted to estimate the total number of unique *E. coli*/*Klebsiella* MLSTs (i.e.: 'species richness') in the population of English BSIs occurring in one year, using two of the best-performing ecological estimators available.[30-32] We found that one of these estimators, preseqR, produced highly uncertain estimates (median: 657 [95% confidence interval (CI): 418–1,032] *E. coli* and 1,322 [623-2,805] *Klebsiella* unique MLSTs), while the other, iNEXT, produced a considerably lower median estimates (545 [438-651] *E. coli* and 872 [687-1,057] *Klebsiella* unique MLSTs; Figure S11). The lower iNEXT estimates are in-line with previous evaluations suggestion this estimator may underestimate diversity[32].

We then used rarefaction (sampling without replacement) to investigate species richness, and three other ecological diversity measures, Shannon diversity, Simpson diversity, and coverage, for their stability to down-sampling, and their sensitivity to novel lineage incursion. We found species richness continued increasing steeply over the entire range of the NEKSUS sample size for all genomic features/genera. Shannon and Simpson diversity were stable from low sample sizes (<500 isolates), while coverage increased steeply at low sample sizes, then tended to plateau (Figure S12). Species richness and coverage showed similarly good sensitivity to incursion, while Shannon and Simpson diversity were insensitive, requiring novel lineages to reach ≥10% prevalence before changing substantially (Figure S13). For comparison, only three of the dominant *E. coli* MLSTs, and none of the *Klebsiella* spp. MLSTs, occurred at a frequency of above 10%.

Given the superior performance of coverage in both stability and sensitivity, coverage was investigated further as a diversity measure for this application to sample size estimation. Four methods of estimating coverage were considered: three ecological estimators (Good's coverage estimator[36,39] on rarefied samples, iNEXT,[30] and preseqR[31]), and Bayesian modelling combined with power calculation (at 90, 95 and 99% certainty). Bayesian estimates showed lower uncertainty, and more conservative (higher) sample size estimates than ecological estimators to reach 80% coverage for *E. coli* MLSTs (i.e.: capture at least one MLSTs of those that collectively constitute ≥80% of the bacterial population), with sample sizes of median: 1,351 (95% credible intervals (CrI): 1,230-1,472) estimates from the Bayesian method at 95% certainty, compared to 390 (95% CI: 2-566) from rarefaction, 374 (328-421) from iNEXT and 441 (147-882) from preseqR (Table S6; Figure 1). For *Klebsiella* MLSTs, although sample size estimates to reach 80% coverage were more similar between the Bayesian [plus power calculation at 95% certainty] and ecological estimators, the latter still showed higher uncertainty, with median 1,422 (95% CrI: 1,331-1,515) *Klebsiella* isolates estimated by the Bayesian method, compared to 1,544 (95% CI: 1,217-1,919) from iNEXT and 1,918 (1,263-3,182) from preseqR. Rarefaction failed to produce a sample size estimate for

80% coverage of *Klebsiella* MLSTs, as the estimated coverage was consistently below 80% at the maximum NEKSUS sample size for *Klebsiella*, 468, and rarefaction cannot produce valid estimates when extrapolated beyond the collected sample size. Considering a wider range of coverages, *E. coli* and *Klebsiella* showed similar patterns where ecological estimators produced lower sample size estimates than Bayesian estimation to reach the same coverage at lower ranges of population coverage, while estimates from different methods became more similar at higher coverages (>90% coverage for *E. coli* and >60% coverage for *Klebsiella*; Figure 1).

### 3. Sensitivity analysis and goodness of fit

#### a. Sensitivity analysis for 'standard' Bayesian models

In standard Bayesian estimation (without regional stratification), the parameters with the strongest influence on the posterior predicted frequency distribution (as compared to the observed NEKSUS data using RMSE) was the prior pseudo-count of isolate-level features, $\alpha$, and the prior counts of success/presence of sub-isolate-level features, $\beta_k$, with progressively worse fitting posteriors as the prior pseudo-count increased from 0.1 to 1000 (Figure S2-S3). This might be expected, since the posterior becomes less informed by the data as its parameters become 'flooded' by very large prior values, and all posterior feature frequencies tend towards a uniform high frequency (i.e.: all features become similarly common in the population) that is dominated by the large prior. This is reflected by scatterplots of observed vs estimated frequencies becoming less steep/closer to horizontal for higher values of $\alpha_k$ or $\beta_k$ (Figure S3). The difference between posterior estimates produced by smaller prior pseudo-count values, 0.1-0.5, compared to the default value, 1, was minimal.

For both isolate-level and sub-isolate-level features, changing the prior number of novel features, m, while keeping the total 'mass' of novel features consistent had no effect on the fit of posterior estimates, apart from $m$=0, as this removed all novel feature mass (Figure S2-3). This suggests that it may not make a big difference to posterior estimates whether novel features are modelled as a single common category, or multiple less frequent categories. However, as the latter prior structure is more intuitive, using multiple low frequency novel feature categories was retained for standard and hierarchical modelling.

For isolate-level features only, increasing the novel mass to very high values (>500 for $\alpha_k$=1) worsened the fit of posterior estimates to observed data, demonstrated by a darkening towards the right of each heatmap (Figure S2a-d), and systematic downward shift of observed vs posterior frequency estimate scatter plots (Figure S3a-d). However, this effect was attenuated for high values of $\alpha_k$. This again might be expected, since a large mass of mutually-exclusive novel features supresses the frequency of observed features, as frequencies have to sum to 1, and this effect becomes less predominant when high prior pseudo-counts dominate the data. This effect was not observed for sub-isolate-level features, where features are non-mutually-exclusive, and therefore frequencies are not constrained to sum to 1, so the addition of novel features does not affect the posterior estimate of the frequencies of observed features (Figure S2e-h; S3e-h).

#### b. Sensitivity analysis for hierarchical Bayesian modelling framework

For isolate-level hierarchical Bayesian models (i.e.: including regionally-stratified data), the prior parameter with the strongest influence on the predictive performance of posterior estimates was, once again, the prior pseudo-count of features, $\alpha_{1:k}$ (i.e.: the values of the elements of the uniform prior vector $\alpha$). The default parameter choice, $\alpha_k = 1$, produced the best, or one of the best predictive performances. For MLSTs, $\alpha_k$=0.1 was the worst-performing prior, while $\alpha_k$ = 10 produced a similar predictive performance to $\alpha_k$=1 for *Klebsiella*, while for *E. coli* MLSTs, $\alpha_k$=1 was the best, followed by $\alpha_k$=10 then $\alpha_k$=0.1. $\alpha_k$ values had similar predictive performance for fastBAPS cluster frequency, with $\alpha_k$ =1, being only marginally better. The number of novel features, $m$, also slightly reduced predictive performance for >500 novel isolates. As for standard Bayesian models, the same intuitive explanation may hold for hierarchical models for this pattern: for mutually-exclusive isolate-level features, as more of the population mass is taken over by 'novel' isolates, the posterior estimated frequency of observed features goes down, as frequencies must sum to 1. The number of novel features had a greater effect that the value of $\alpha_k$ for fastBAPS clusters, while opposite was true for MLSTs (Figure S4a-d).

$\mu_{\mu log\tau}$, $\sigma_{\mu log\tau}$, and $\lambda_{\sigma log\tau}$, the three hyperparameters defining the distribution of the region-specific concentration parameter, $\tau_r$, had minimal influence on predictive performance of the model, and instead affected chain convergence, i.e. how well the Markov chain was able to explore the posterior parameter space to produce unbiased estimates. Values of $\lambda_{\sigma log\tau}$ deviating from 1, especially lower values of 0.1, produced more models failing chain convergence diagnostic checks, as well as more highly variable estimates. Notably, some parameter combinations with $\lambda_{\sigma log\tau}$=0.1 performed better than expected given their neighbouring parameter space, although these estimates are highly likely to be biased, as models run with these prior parameter combinations failed chain convergence diagnostic checks. Lower values of $\sigma_{\mu log\tau}$ also produced better mixing chains (with a higher proportion of models passing chain diagnostics). Varying the value of $\mu_{\mu log\tau}$ in the range 0.1-10 seemed to have no consistent substantial effect on either predictive performance of model mixing (Figure S4).

For sub-isolate-level hierarchical Bayesian models, the patterns of effect of prior parameters were largely similar compared to those for isolate-level features: the strongest influence on model predictive performance was from the parameters of the prior distribution defining prior isolate frequency ($\beta_k$ and $\beta'_k$), a very high number of novel features, $m$ >1000, slightly reduced predictive performance, and the three regional concentration hyperparameters, $\mu_{\mu log\tau}$, $\sigma_{\mu log\tau}$, and $\lambda_{\sigma log\tau}$, had minimal effect on predictive performance and instead had a small impact on chain convergence. A notable difference was that the Beta prior distribution for these models had two parameters ($\beta_k$ and $\beta'_k$). The default value chosen for the first parameter, the prior pseudo-count of success/feature presence, $\beta_k$=1 tended to have the best fit for all models, while a larger value for the prior pseudo-count of failure/feature absence, $\beta'_k$ =100, produced a better fit. This effect was most marked for plasmids, while for ARGs, the prior value of $\beta'_k$ made little difference with $\beta_k$ =1. This may be explained by the highly right-skewed distribution of plasmid subcommunities, with many singleton subcommunities, and many 0-count subcommunities in certain regions, which makes the also highly right-skewed Beta(1,100) prior distribution more appropriate for plasmids compared to a uniform/symmetric distribution (Figure S4e-h).

**c. Goodness of fit**

Overall, hierarchical Bayesian models run with default parameter combinations (Table S4) produced excellent fits to overall observed feature frequencies for *E. coli* MLSTs, and *E. coli* and *Klebsiella* fastBAPS clusters, good fits for *E. coli* and *Klebsiella* ARGs, and poor fits to overall plasmid frequencies for both genera and *Klebsiella* MLSTs (Figure S5). In particular, the posterior frequency estimates of *Klebsiella* MLSTs were generally underestimated, especially for more common features (Figure S5b), which may potentially inflate posterior estimates of the sample sizes required to capture a certain coverage. For plasmids of both genera, plasmid subcommunity frequency tended to be over-estimated for rarer subcommunities (Figure 5e-f), which is likely to shrink resultant sample size estimates. For ARGs, rare gene frequency was also slightly over-estimated, but by a much smaller magnitude than for plasmids. Despite this, regional feature frequencies were predicted well by all modelling frameworks, including for plasmids, except Klebsiella MLST, where there was under-estimation of regional feature frequencies, especially in regions with few unique MLST types (Figure S6b).

### Supplementary Tables

| Search number | Search terms | Number of results |
|---|---|---|
| 1 | exp Public Health Surveillance/ | 5438 |
| 2 | Epidemiological Monitoring/ | 8978 |
| 3 | public health surveillance.mp. | 9359 |
| 4 | surveillance.mp. | 356614 |
| 5 | sampl* size calc*.mp. [mp=title, book title, abstract, original title, name of substance word, subject heading word, floating sub-heading word, keyword heading word, organism supplementary concept word, protocol supplementary concept word, rare disease supplementary concept word, unique identifier, synonyms, population supplementary concept word, anatomy supplementary concept word] | 4023 |
| 6 | sampl* size estim*.mp. [mp=title, book title, abstract, original title, name of substance word, subject heading word, floating sub-heading word, keyword heading word, organism supplementary concept word, protocol supplementary concept word, rare disease supplementary concept word, unique identifier, synonyms, population supplementary concept word, anatomy supplementary concept word] | 1333 |
| 7 | 1 or 2 or 3 or 4 | 360712 |
| 8 | 5 or 6 | 5213 |
| 9 | 7 and 8 | 62 |

**Supplementary Table S1: Search strategy and number of records retrieved from MEDLINE (accessed via Ovid), from 1946 to 13/05/2026.**

| Publication | Area | Frequentist sample size | Modelling / simulation | Other exact calculation | surveillance goal | Sampling over time | Account for non-random disease distribution | Account for non-random/ complex sampling | Account for imperfect test sensitivity/ specificity | Account for missing data | Comment |
|---|---|---|---|---|---|---|---|---|---|---|---|
| Green[41] | Veterinary (freedom form disease) | Y | - | Y | presence/absence | - | - | - | Y | - | Exact solution feasible compared to simulation based. |
| Belsare et al[42] | Veterinary (wildlife surveillance via hunting) | - | Agent based modelling | - | presence/absence | - | Y | Y | - | - | |
| De Blas et al.[43] | Veterinary (aquaculture) | - | Random statically simulated population | - | presence/absence, prevalence | - | Y | Y | - | - | Use a finite population sample size formula. |
| Wohl et al.[44] | COVID-19 genomic surveillance | Y | - | Y | presence/absence, prevalence, time to detection | Y | - | Y | Y | - | Account for detection bias of more severe/ VOC infections, and time to detection. |
| Niu et al.[45] | HIV/AIDS dynamics at subnational level | - | Generalised linear mixed effects models | - | presence/absence | - | - | Y | - | Y | Regression including borrowing from data rich areas improves prediction from EPP package |
| Fearon et al.[46] | Concealed population size estimation (sex workers, Zimbabwe) | Y | Multiplier methods (reduce random error) | - | population size | - | - | Y | - | - | Multiplier method aims to estimate total population size from respondent-driven sampling surveys |
| Vinh & Boni[47] | Influenza seroepidemiology | - | Serial sero-epidemiological model | - | presence/absence | Y | - | - | - | - | Alternative likelihood-based approach to modelling disease dynamics, avoids estimating proportion of infections detected. |
| Weijnert et al.,[48] | Respondent driven sampling of injection drug users in US | Y | - | - | presence/absence, population size | - | - | Y | - | - | Design effect should be 4, not 2, as per previous studies (estimates parameter from 43 existing datasets). |
| Reist et al., [49] | Veterinary (public health risk-based surveillance) | Y | - | - | presence/absence, prevalence | Y | - | - | - | - | Includes scenario tree modelling, risk-based [drug] residue monitoring, qualitative analysis, and accounting for data from previous surveys. |
| Nanan et al., [50] | Low-prevalence HIV prevalence estimation | Y | - | - | prevalence | - | - | Y | - | - | Conclude that sentinel surveillance should be used for low-prevalence HIV settings. |
| Fu & Xi[51] | Post-marketing evaluation of Chinese traditional medicine | Y | - | - | prevalence | - | - | Y | - | - | Qualitative discussion based on classical sample size calculations, application to new domain. |
| Feng et al[52] | Post-marketing evaluation of Chinese traditional medicine | Y | - | - | prevalence, difference in effect | - | - | Y | - | - | Discussion of considerations and application of sample size estimation to complex RCT designs. |
| Segura-Correa et al., [53] | Veterinary (cattle diseases in Mexico) | Y | - | - | prevalence | - | - | Y | Y | - | Estimated parameters for 2-stage sampling (intra-herd correlation coefficient and design effect), Bayesian accounting for test sensitivity and specificity. |
| Ogungbenro & Aarons[54] | Population pharmacodynamic experiments | Y | Mixed effects modelling | - | difference in effect | Y | - | Y | - | - | Method development accounting from serial sampling from the same individual. |
| Ngondi et al.,[55] | Trachoma surveillance | Y | - | - | prevalence | - | Y | Y | - | - | Review 3 trachoma survey protocols: cluster random sampling better than trachoma rapid assessment, and acceptance sampling trachoma rapid assessment |
| Fosgate[56] | Veterinary | Y | - | - | presence/absence, diagnostic test accuracy | - | - | - | - | Y | Review of sample size calculation methods, discussion of practical implementation tools; mention need to increase sample size by expected data/animal loss |
| Knopf et al.,[57] | Veterinary (cattle disease in Switzerland) | - | Stochastic simulation model | - | presence/absence | Y | - | - | - | - | Stochastic model for risk-based sample size calculation for serological surveys for disease freedom for consecutive national surveys. Tool for public use. |

| Publication | Area | Frequentist sample size | Modelling / simulation | Other exact calculation | surveillance goal | Sampling over time | Account for non-random disease distribution | Account for non-random/ complex sampling | Account for imperfect test sensitivity/ specificity | Account for missing data | Comment |
|---|---|---|---|---|---|---|---|---|---|---|---|
| Taylor et al.,[58] | Sever malaria in African children | Y | - | - | Sprevalence, incidence | - | - | Y | - | Y | Estimate expected prevalence / incidence of severe malarial syndromes to inform future study design. Network of sited needs to expand or 3-4 years needed |
| Stark et al. [59] | Veterinary (risk-based surveillance) | - | - | - | presence/absence, prevalence | - | - | Y | - | - | Review of qualitative risk-based sampling based on risk assessment. |
| Vansteelandt et al.[60] | HIV in Kenya | - | Regression from pooled sera | | prevalence | - | - | Y | Y | - | Age-homogenised optimal pool size of 7 reduces cost by 44% vs individual samples with no loss of precision |
| Tubert-bitter et al., [61] | Pharmacoepidemiology (post-market cohort studies) | Y | - | Y | incidence | - | - | - | - | - | Poisson distribution derived sample size estimates are more appropriate for rare events like adverse drug reactions than the Normal distribution assumption |
| Gillum et al., [62] | Total community trials | Y | - | - | incidence | Y | - | Y | - | - | Describe method of computing event rate in community trials that accounts for in- and out-migration. |

**Supplementary Table S2: Results of the systematic literature search for methods of sample size calculation for surveillance.**

| Isolate-level feature priors | $\alpha_{1:k}$ | $\beta_{1:k}$ | *m* novel feature number | Σ*m* novel feature mass | Frequency of singletons in NEKSUS | θ (novelty hungriness) median (95% CrI) | *u* (median novelty probability) |
|---|---|---|---|---|---|---|---|
| **NEKSUS Dataset** | | | | | | | |
| ***E. coli* MLSTs** | 1 | *NA* | 12 | 12 | 0.109 | 62.3 (54.4-71.4) | 0.041 |
| ***Klebsiella* MLSTs** | 1 | *NA* | 71 | 71 | 0.476 | 111.2 (97.0-126.9) | 0.192 |
| ***E. coli* fastBAPS clusters** | 1 | *NA* | 4 | 4 | 0.028 | 33.4 (28.1 – 39.5) | 0.022 |
| ***Klebsiella* fastBAPS clusters** | 1 | *NA* | 1 | 1 | 0.023 | 5.1 (3.3 – 7.5) | 0.011 |
| ***E. coli* Plasmids** | *NA* | 1 | 605 | 605 | 0.423 | *NA* | 0.423 |
| ***Klebsiella* Plasmids** | *NA* | 1 | 998 | 998 | 0.705 | *NA* | 0.705 |
| ***E. coli* ARGs** | *NA* | 1 | 9 | 9 | 0.041 | *NA* | 0.041 |
| ***Klebsiella* ARGs** | *NA* | 1 | 32 | 32 | 0.137 | *NA* | 0.137 |
| **Sensitivity analysis parameter ranges** | | | | | | | |
| ***E. coli* MLST sensitivity analysis** | 0.1-1000 | *NA* | 0-1000 | 0-1000 | *NA* | *NA* | *NA* |
| ***Klebsiella* MLST sensitivity analysis** | 0.1-1000 | *NA* | 0-1000 | 0-1000 | *NA* | *NA* | *NA* |
| ***E. coli* fastBAPS sensitivity analysis** | 0.1-1000 | *NA* | 0-1000 | 0-1000 | *NA* | *NA* | *NA* |
| ***Klebsiella* fastBAPS sensitivity analysis** | 0.1-1000 | *NA* | 0-1000 | 0-1000 | *NA* | *NA* | *NA* |
| ***E. coli* Plasmid sensitivity analysis** | *NA* | 0.1-1000 | 0-1000 | 0-1000 | *NA* | *NA* | *NA* |
| ***Klebsiella* Plasmid sensitivity analysis** | *NA* | 0.1-1000 | 0-1000 | 0-1000 | *NA* | *NA* | *NA* |
| ***E. coli* ARG sensitivity analysis** | *NA* | 0.1-1000 | 0-1000 | 0-1000 | *NA* | *NA* | *NA* |
| ***Klebsiella* ARG sensitivity analysis** | *NA* | 0.1-1000 | 0-1000 | 0-1000 | *NA* | *NA* | *NA* |
| **External Datasets** | | | | | | | |
| **NORM *E. coli* MLSTs (2011-2017)** | 1 | *NA* | 11 | 11 | 0.097 | 67.1 (59.0-76.1) | 0.033 |
| **BSAC *E. coli* MLSTs (2003-2012)** | 1 | *NA* | 8 | 8 | 0.099 | 45.9 (39.5-53.6) | 0.033 |

**Supplementary Table S3: Prior parameters used in standard Bayesian modelling, and prior parameter ranges investigated in sensitivity analyses for *E. coli* and *Klebsiella* MLSTs, fastBAPS clusters, plasmid subcommunities, and antimicrobial resistance genes (ARGs) in the NEKSUS dataset, and external validation dataset.** $\alpha_{1:k}$ represents the prior pseudo-count for isolate-level features (where feature labels are mutually-exclusive in isolates), and $\beta_{1:k}$ represents the prior pseudo-count of presence for sub-isolate-level features (where feature labels are non-mutually-exclusive in isolates, and each feature has a 50:50 prior probability of presence/absence, so that the two prior Beta distribution parameters are equal $\beta = \beta'$ ). θ represents the 'novelty hungriness' parameter estimated from applying the Chinese restaurant process to the NEKSUS data on categorical counts of mutually-exclusive features, therefore only applying to isolate-level features, not to sub-isolate-level features. *u* represents the 'novelty probability'. For isolate-level features, u = θ/(N + θ), as defined by the Chinese restaurant process[38], and for sub-isolate-level features, u was equal to the frequency of singleton features, as defined by Good-Turing[39]. The number of novel features, *m*, was given by $\frac{uK}{1-u}$ , so that, given all novel and observed features had the same prior, the combined prior 'mass' of all novel features was equal to the novelty probability from the data. The novel feature mass, Σ*m*, was given by the novel feature number times the prior pseudo-count (*α* or *β*). Note that only the $2^{nd}$-$5^{th}$ columns form the left were directly included as model priors and other columns were used in intermediate calculations. Abbreviations: CrI – credible interval; NEKSUS – National *E. coli* and *Klebsiella* bloodstream infection and carbapenemase-producing Enterobacterales UK surveillance study; NORM – Norwegian Surveillance Program on Resistant Microbes;[63] BSAC – British Society of Antimicrobial Chemotherapy;[64] MLST – Multi-Locus Sequence Type.

| Metric / Model | Prior parameters | | | | | | | | Model Diagnostics | | | | | |
|---|---|---|---|---|---|---|---|---|---|---|---|---|---|---|
| | **$\alpha_k$** | **$\beta_k$** | **$\beta'_k$** | ***m* novel feature number** | **Σ*m* novel feature mass** | **$\mu_{\mu_{\log\tau}}$** | **$\sigma_{\mu_{\log\tau}}$** | **$\lambda_{\sigma_{\log\tau}}$** | **Number of divergent transitions *** | **Number of max tree depth transitions** | **Mean E-BFMI** | **Mean ESS Bulk** | **Mean ESS Tail** | **Mean $\hat{R}$** |
| ***E. coli* MLST** | 1 | *NA* | *NA* | 12 | 12 | 1 | 1 | 1 | 5 | 0 | 0.9241 | 6317 | 2430 | 1.0009 |
| ***Klebsiella* MLST** | 1 | *NA* | *NA* | 71 | 71 | 1 | 1 | 1 | 2 | 0 | 0.9005 | 6367 | 2369 | 1.0008 |
| ***E. coli* fastBAPS cluster** | 1 | *NA* | *NA* | 4 | 4 | 1 | 1 | 1 | 27 | 0 | 0.9223 | 4051 | 2254 | 1.0013 |
| ***Klebsiella* fastBAPS cluster** | 1 | *NA* | *NA* | 1 | 1 | 1 | 1 | 1 | 101 | 0 | 0.5192 | 1996 | 1691 | 1.0037 |
| ***E. coli* plasmids** | *NA* | 1 | 50 | 605 | 30,250 | 1 | 1 | 1 | 0 | 0 | 0.9056 | 6134 | 2358 | 1.0008 |
| ***Klebsiella* plasmids** | *NA* | 1 | 50 | 998 | 49,900 | 1 | 1 | 1 | 0 | 0 | 0.8915 | 5875 | 2299 | 1.0007 |
| ***E. coli* ARGs** | *NA* | 1 | 1 | 9 | 9 | 1 | 1 | 1 | 6 | 0 | 0.9460 | 4341 | 2360 | 1.0010 |
| ***Klebsiella* ARGs** | *NA* | 1 | 1 | 32 | 32 | 1 | 1 | 1 | 0 | 0 | 0.9274 | 4836 | 2432 | 1.0009 |
| **Sensitivity analyses** | | | | | | | | | | | | | | |
| ***E. coli* MLST sensitivity analysis** | 0.1-10 | *NA* | *NA* | 1-1000 | $m*\alpha_k$ | 0.1-10 | 0.1-10 | 0.1-10 | *NA* | *NA* | *NA* | *NA* | *NA* | *NA* |
| ***Klebsiella* MLST sensitivity analysis** | 0.1-10 | *NA* | *NA* | 1-1000 | $m*\alpha_k$ | 0.1-10 | 0.1-10 | 0.1-10 | *NA* | *NA* | *NA* | *NA* | *NA* | *NA* |
| ***E. coli* fastBAPS sensitivity analysis** | 0.1-10 | *NA* | *NA* | 1-1000 | $m*\alpha_k$ | 0.1-10 | 0.1-10 | 0.1-10 | *NA* | *NA* | *NA* | *NA* | *NA* | *NA* |
| ***Klebsiella* fastBAPS sensitivity analysis** | 0.1-10 | *NA* | *NA* | 1-1000 | $m*\alpha_k$ | 0.1-10 | 0.1-10 | 0.1-10 | *NA* | *NA* | *NA* | *NA* | *NA* | *NA* |
| ***E. coli* Plasmid sensitivity analysis** | *NA* | 0.1-10 | 0.1-100 | 10-1000 | $m*\beta_k$ | 0.1-10 | 0.1-10 | 0.1-10 | *NA* | *NA* | *NA* | *NA* | *NA* | *NA* |
| ***Klebsiella* Plasmid sensitivity analysis** | *NA* | 0.1-10 | 0.1-100 | 10-1000 | $m*\beta_k$ | 0.1-10 | 0.1-10 | 0.1-10 | *NA* | *NA* | *NA* | *NA* | *NA* | *NA* |
| ***E. coli* ARG sensitivity analysis** | *NA* | 0.1-10 | 0.1-100 | 10-1000 | $m*\beta_k$ | 0.1-10 | 0.1-10 | 0.1-10 | *NA* | *NA* | *NA* | *NA* | *NA* | *NA* |
| ***Klebsiella* ARG sensitivity analysis** | *NA* | 0.1-10 | 0.1-100 | 10-1000 | $m*\beta_k$ | 0.1-10 | 0.1-10 | 0.1-10 | *NA* | *NA* | *NA* | *NA* | *NA* | *NA* |

**Supplementary Table S4: Priors parameters, prior parameter ranges and chain diagnostics for hierarchical Bayesian models, and for *E. coli* and *Klebsiella* MLSTs, fastBAPS clusters, plasmid subcommunities, and antimicrobial resistance genes (ARGs).** Each model was run with 4 Markov chains, each with 1000 warm-up and 1000 sampling iterations, whereas each parameter combination in sensitivity analyses was run with 1 Markov chain with 250 warm-up and 250 sampling iterations. *Divergent transitions and transitions hitting the maximum tree depth are shown as the sum of the four Markov chains and may indicate poorly or inefficiently explored posteriors geometry. All other chain diagnostic metrics are shown as the mean across all chains/parameters. Abbreviations: E-BMFI – Energy Bayesian Fraction of Missing Information, where >0.3 indicates adequate efficiency of momentum exploration in the Markov chain; ESS – effective sample size, where ESS bulk and ESS tail >1000 indicate efficient sampling from the centre and the tails of the posterior, respectively; $\hat{R}$– close to 1 (i.e.: <0.01) indicates good agreement between chains[65].

| | Isolates | | | Plasmids | | ARGs | |
|---|---|---|---|---|---|---|---|
| | Total | Unique MLSTs | Unique fastBAPS clusters | Total | Unique subcommunities | Total | Unique ARGs |
| ***E. coli*** | | | | | | | |
| **East** | 138 | 52 | 67 | 298 | 134 | 1,100 | 87 |
| **London** | 95 | 47 | 56 | 225 | 113 | 817 | 64 |
| **Midlands** | 95 | 47 | 52 | 193 | 88 | 693 | 81 |
| **North East A** | 283 | 82 | 88 | 753 | 257 | 2,407 | 109 |
| **North East B** | 185 | 61 | 71 | 453 | 187 | 1,594 | 95 |
| **North West** | 86 | 29 | 45 | 192 | 92 | 663 | 67 |
| **South East A** | 208 | 75 | 84 | 390 | 171 | 1,420 | 96 |
| **South East B** | 35 | 20 | 25 | 64 | 40 | 234 | 54 |
| **South East C** | 118 | 45 | 59 | 275 | 125 | 771 | 67 |
| **South West** | 228 | 75 | 84 | 518 | 200 | 1,589 | 88 |
| **Overall** | 1,471 | 263 | 161 | 3,361 | 825 | 11,288 | 189 |
| ***Klebsiella*** | | | | | | | |
| **East** | 65 | 60 | 15 | 105 | 81 | 475 | 83 |
| **London** | 28 | 22 | 6 | 55 | 42 | 197 | 52 |
| **Midlands** | 29 | 27 | 12 | 47 | 33 | 218 | 75 |
| **North East A** | 90 | 71 | 16 | 168 | 115 | 639 | 97 |
| **North East B** | 61 | 49 | 13 | 141 | 80 | 475 | 82 |
| **North West** | 24 | 22 | 8 | 69 | 53 | 178 | 63 |
| **South East A** | 88 | 73 | 15 | 128 | 80 | 544 | 86 |
| **South East B** | 6 | 5 | 3 | 15 | 9 | 78 | 35 |
| **South East C** | 36 | 34 | 10 | 60 | 49 | 227 | 68 |
| **South West** | 41 | 36 | 12 | 59 | 43 | 227 | 60 |
| **Overall** | 468 | 295 | 29 | 847 | 417 | 3,258 | 199 |

**Supplementary Table S5: Summary of the total number of bacterial isolates, plasmids, and antimicrobial resistance genes (ARGs) and unique MLSTs, fastBAPS clusters, plasmid pling subcommunities, and ARGs per region and overall (nationally) assembled and annotated in the NEKSUS dataset for *E. coli* and *Klebsiella* bloodstream infection (BSI) isolates.**

| Estimator \ Coverage | 75% | 80% | 85% | 90% | 95% | 99% |
|---|---|---|---|---|---|---|
| **NEKSUS *E. coli*** | | | | | | |
| **MLST 90%** | 821 (729 - 907) | 1,038 (946 - 1,131) | 1,314 (1,209 - 1,419) | 1,721 (1,595 - 1,851) | 2,513 (2,306 - 2,735) | 5,187 (4,547 - 5,897) |
| **MLST 95%** | 1,068 (949 - 1,181) | 1,351 (1,230 - 1,472) | 1,710 (1,573 - 1,846) | 2,239 (2,075 - 2,409) | 3,270 (3,001 - 3,559) | 6,749 (5,917 - 7,672) |
| **MLST 99%** | 1,643 (1,459 - 1,815) | 2,077 (1,892 - 2,263) | 2,629 (2,418 - 2,838) | 3,443 (3,191 - 3,703) | 5,027 (4,613 - 5,471) | 10,375 (9,095 - 11,794) |
| **MLST Rarefaction (ecological)** | 201 (2 - 330) | 390 (2 - 566) | 906 (2 - 1,114) | NA (2 - NA) | NA (2 - NA) | NA (2 - NA) |
| **MLST iNEXT (ecological)** | 234 (187 - 234) | 374 (328 - 421) | 749 (655 - 889) | 1,732 (1,451 - 2,059) | 3,510 (2,902 - 4,212) | 7,628 (6,131 - 8,892) |
| **MLST preseqR (ecological)** | 294 (147 - 588) | 441 (147 - 882) | 882 (147 - 1,323) | 1,765 (735 - 2,353) | 3,971 (2,794 - 5,589) | 13,386 (7,355 - 23,388) |
| **fastBAPS L3 90%** | 404 (355 - 454) | 499 (441 - 558) | 637 (564 - 712) | 864 (765 - 964) | 1,336 (1,180 - 1,500) | 2,944 (2,487 - 3,473) |
| **fastBAPS L3 95%** | 526 (463 - 591) | 649 (575 - 727) | 828 (734 - 926) | 1,124 (995 - 1,255) | 1,738 (1,535 - 1,952) | 3,830 (3,236 - 4,518) |
| **fastBAPS L3 99%** | 809 (711 - 909) | 999 (883 - 1,118) | 1,274 (1,128 - 1,424) | 1,728 (1,530 - 1,929) | 2,671 (2,360 - 3,001) | 5,888 (4,975 - 6,946) |
| **Plasmid subcommunity 90%** | 1,230 (1,167 - 1,294) | 1,482 (1,412 - 1,556) | 1,832 (1,747 - 1,920) | 2,389 (2,274 - 2,511) | 3,586 (3,403 - 3,776) | 8,415 (7,828 - 9,043) |
| **Plasmid subcommunity 95%** | 1,599 (1,518 - 1,683) | 1,928 (1,837 - 2,025) | 2,383 (2,273 - 2,497) | 3,108 (2,959 - 3,266) | 4,666 (4,427 - 4,913) | 10,949 (10,184 - 11,765) |
| **Plasmid subcommunity 99%** | 2,459 (2,334 - 2,588) | 2,963 (2,823 - 3,112) | 3,663 (3,493 - 3,839) | 4,778 (4,548 - 5,021) | 7,172 (6,806 - 7,552) | 16,830 (15,656 - 18,086) |
| **ARGs 90%** | 16 (14 - 18) | 21 (18 - 24) | 36 (31 - 41) | 70 (61 - 79) | 162 (139 - 189) | 1,184 (1,058 - 1,315) |
| **ARGs 95%** | 21 (18 - 23) | 27 (23 - 30) | 47 (40 - 54) | 91 (80 - 103) | 211 (180 - 246) | 1,540 (1,376 - 1,711) |
| **ARGs 99%** | 32 (28 - 34) | 41 (34 - 47) | 72 (62 - 82) | 139 (122 - 158) | 323 (277 - 377) | 2,367 (2,116 - 2,630) |
| **NEKSUS *Klebsiella*** | | | | | | |
| **MLST 90%** | 956 (896 - 1,018) | 1,093 (1,023 - 1,164) | 1,283 (1,197 - 1,373) | 1,588 (1,473 - 1,707) | 2,232 (2,041 - 2,440) | 4,693 (4,057 - 5,427) |
| **MLST 95%** | 1,243 (1,165 - 1,324) | 1,422 (1,331 - 1,515) | 1,669 (1,557 - 1,787) | 2,067 (1,917 - 2,221) | 2,905 (2,655 - 3,175) | 6,107 (5,279 - 7,061) |
| **MLST 99%** | 1,912 (1,792 - 2,036) | 2,186 (2,047 - 2,329) | 2,566 (2,395 - 2,747) | 3,177 (2,946 - 3,415) | 4,465 (4,082 - 4,881) | 9,387 (8,115 - 10,854) |
| **MLST Rarefaction (ecological)** | NA | NA | NA | NA | NA | NA |
| **MLST iNEXT (ecological)** | 1,264 (983 - 1,591) | 1,544 (1,217 - 1,919) | 1,872 (1,451 - 2,340) | 2,387 (1,825 - 2,902) | 3,229 (2,434 - 3,838) | 5,148 (3,838 - 5,990) |
| **MLST preseqR (ecological)** | 1,497 (1,029 - 2,340) | 1,918 (1,263 - 3,182) | 2,574 (1,544 - 4,492) | 3,650 (2,012 - 7,113) | 6,037 (2,667 - NA) | NA (3,744 - NA) |
| **fastBAPS L3 90%** | 68 (44 - 90) | 87 (65 - 111) | 112 (85 - 145) | 155 (115 - 206) | 278 (194 - 369) | 759 (521 - 1,059) |
| **fastBAPS L3 95%** | 89 (57 - 117) | 113 (84 - 145) | 146 (111 - 188) | 202 (150 - 268) | 362 (253 - 480) | 987 (677 - 1,378) |
| **fastBAPS L3 99%** | 138 (89 - 181) | 175 (130 - 223) | 224 (172 - 290) | 311 (231 - 412) | 557 (388 - 739) | 1,518 (1,042 - 2,118) |
| **Plasmid subcommunity 90%** | 731 (690 - 774) | 857 (809 - 908) | 1,039 (980 - 1,102) | 1,341 (1,263 - 1,422) | 2,012 (1,887 - 2,150) | 4,825 (4,440 - 5,253) |
| **Plasmid subcommunity 95%** | 951 (897 - 1,006) | 1,115 (1,052 - 1,181) | 1,352 (1,276 - 1,433) | 1,744 (1,642 - 1,849) | 2,618 (2,455 - 2,796) | 6,277 (5,776 - 6,834) |
| **Plasmid subcommunity 99%** | 1,461 (1,379 - 1,547) | 1,714 (1,618 - 1,815) | 2,078 (1,960 - 2,203) | 2,681 (2,525 - 2,843) | 4,024 (3,774 - 4,299) | 9,650 (8,879 - 10,505) |
| **ARGs 90%** | 48 (42 - 55) | 64 (56 - 73) | 103 (86 - 121) | 186 (163 - 211) | 341 (307 - 376) | 849 (747 - 964) |
| **ARGs 95%** | 62 (54 - 71) | 83 (72 - 95) | 134 (112 - 158) | 242 (211 - 274) | 444 (399 - 489) | 1,104 (972 - 1,254) |
| **ARGs 99%** | 95 (83 - 109) | 128 (111 - 146) | 205 (172 - 242) | 372 (325 - 421) | 681 (613 - 751) | 1,697 (1,494 - 1,927) |
| **External Datasets** | | | | | | |
| **NORM *E. coli* MLSTs 90%** | 927 (782 - 1,081) | 1,440 (1,281 - 1,597) | 2,051 (1,889 - 2,206) | 2,879 (2,697 - 3,065) | 4,419 (4,135 - 4,706) | 9,417 (8,550 - 10,367) |
| **NORM *E. coli* MLSTs 95%** | 1,207 (1,018 - 1,407) | 1,874 (1,667 - 2,079) | 2,669 (2,457 - 2,870) | 3,746 (3,510 - 3,988) | 5,749 (5,379 - 6,123) | 12,252 (11,123 - 13,488) |
| **NORM *E. coli* MLSTs 99%** | 1,855 (1,564 - 2,163) | 2,881 (2,563 - 3,195) | 4,102 (3,778 - 4,412) | 5,759 (5,395 - 6,130) | 8,838 (8,270 - 9,413) | 18,835 (17,100 - 20,735) |
| **BSAC *E. coli* MLSTs 90%** | 584 (488 - 673) | 785 (694 - 873) | 1,030 (930 - 1,127) | 1,383 (1,264 - 1,506) | 2,062 (1,877 - 2,266) | 4,333 (3,747 - 4,988) |
| **BSAC *E. coli* MLSTs 95%** | 760 (634 - 876) | 1,022 (902 - 1,137) | 1,340 (1,210 - 1,466) | 1,799 (1,645 - 1,960) | 2,683 (2,442 - 2,948) | 5,638 (4,874 - 6,489) |
| **BSAC *E. coli* MLSTs 99%** | 1,168 (976 - 1,346) | 1,571 (1,388 - 1,747) | 2,060 (1,860 - 2,255) | 2,766 (2,528 - 3,013) | 4,124 (3,755 - 4,532) | 8,667 (7,494 - 9,976) |

**Supplementary Table S6: *E. coli* and *Klebsiella* sequencing sample size estimates to reach different coverage thresholds (the proportion of the population represented by a feature detected in the sample), obtained from Bayesian modelling of frequency distributions combined with power calculation of different genomic features at 90, 95, and 99% certainty.**

Ecological estimators (MLST rarefaction, iNEXT, and preseqR) are shown for comparison. These were not applied to other genomic features beyond MLST due to their limitations (inability to extrapolate beyond the actual sample, systematic underestimation of diversity, and high uncertainty). 'NA' in the rarefaction estimates indicates that that level of coverage could not be reached by sampling without replacement. The maximal coverage reached for *Klebsiella* MLSTs by rarefaction (sampling without replacement) was ~55%, hence all rarefaction estimates are NA. Note that results from fastBAPS level 3 are used elsewhere in the results. Level 1 corresponds to clonal complexes/clonal groups, and levels 2-3 partition the population into progressively smaller clusters; however, this did not produce robust clustering for *Klebsiella*. Abbreviations: ARG – Antimicrobial resistance gene; NEKSUS – National *E. coli* and *Klebsiella* bloodstream infection and carbapenemase-producing Enterobacterales UK surveillance study; NORM – Norwegian Surveillance Program on Resistant Microbes[63] (data from 2011-2017 were included to capture the period of author-defined stable population structure); BSAC – British Society of Antimicrobial Chemotherapy[64] (data from 2003-2012 were included to capture the period of author-defined stable population structure); MLST – Multi-locus sequence type.

| Feature \ Species richness | 30% | 40% | 50% | 60% | 70% | 80% | 90% | 95% |
|---|---|---|---|---|---|---|---|---|
| MLST 90% | 1,116 (1,015 - 1,232) | 1,417 (1,289 - 1,562) | 1,780 (1,613 - 1,978) | 2,227 (1,993 - 2,502) | 2,886 (2,548 - 3,297) | 3,933 (3,385 - 4,649) | 6,462 (5,239 - 8,177) | 10,347 (7,707 - 14,750) |
| MLST 95% | 1,452 (1,321 - 1,603) | 1,844 (1,678 - 2,032) | 2,317 (2,099 - 2,574) | 2,898 (2,594 - 3,255) | 3,755 (3,315 - 4,290) | 5,118 (4,404 - 6,048) | 8,407 (6,817 - 10,639) | 13,462 (10,027 - 19,190) |
| MLST 99% | 2,233 (2,030 - 2,465) | 2,834 (2,579 - 3,124) | 3,561 (3,227 - 3,957) | 4,455 (3,987 - 5,004) | 5,773 (5,097 - 6,595) | 7,867 (6,770 - 9,298) | 12,924 (10,479 - 16,355) | 20,694 (15,415 - 29,500) |
| fastBAPS L3 90% | 406 (361 - 457) | 568 (504 - 643) | 788 (694 - 899) | 1,061 (927 - 1,225) | 1,474 (1,263 - 1,753) | 2,106 (1,745 - 2,595) | 3,618 (2,768 - 4,921) | 5,685 (3,964 - 8,887) |
| fastBAPS L3 95% | 528 (470 - 594) | 739 (656 - 837) | 1,025 (902 - 1,170) | 1,381 (1,206 - 1,594) | 1,918 (1,643 - 2,281) | 2,739 (2,271 - 3,376) | 4,707 (3,601 - 6,403) | 7,396 (5,157 - 11,562) |
| fastBAPS L3 99% | 812 (722 - 913) | 1,136 (1,009 - 1,287) | 1,576 (1,388 - 1,799) | 2,123 (1,855 - 2,451) | 2,949 (2,526 - 3,507) | 4,211 (3,491 - 5,190) | 7,236 (5,536 - 9,843) | 11,370 (7,928 - 17,775) |
| Plasmids 90% | 1,465 (1,387 - 1,549) | 1,900 (1,796 - 2,013) | 2,461 (2,317 - 2,619) | 3,257 (3,045 - 3,487) | 4,528 (4,187 - 4,905) | 6,996 (6,340 - 7,748) | 14,215 (12,308 - 16,496) | 28,626 (23,068 - 35,681) |
| Plasmids 95% | 1,905 (1,804 - 2,014) | 2,472 (2,336 - 2,619) | 3,201 (3,014 - 3,407) | 4,237 (3,961 - 4,536) | 5,891 (5,447 - 6,381) | 9,101 (8,249 - 10,080) | 18,495 (16,013 - 21,461) | 37,242 (30,012 - 46,421) |
| Plasmids 99% | 2,929 (2,773 - 3,097) | 3,799 (3,592 - 4,026) | 4,921 (4,634 - 5,237) | 6,514 (6,090 - 6,973) | 9,055 (8,373 - 9,810) | 13,991 (12,680 - 15,495) | 28,430 (24,615 - 32,991) | 57,251 (46,135 - 71,361) |
| ARGs 90% | 230 (192 - 279) | 575 (495 - 667) | 841 (734 - 971) | 1,162 (1,009 - 1,348) | 1,597 (1,366 - 1,881) | 2,309 (1,926 - 2,829) | 3,990 (3,112 - 5,381) | 6,693 (4,672 - 10,615) |
| ARGs 95% | 298 (250 - 362) | 748 (645 - 868) | 1,094 (955 - 1,263) | 1,511 (1,312 - 1,754) | 2,077 (1,777 - 2,447) | 3,005 (2,506 - 3,680) | 5,191 (4,048 - 7,000) | 8,708 (6,078 - 13,811) |
| ARGs 99% | 459 (384 - 557) | 1,149 (990 - 1,333) | 1,681 (1,468 - 1,942) | 2,323 (2,017 - 2,695) | 3,193 (2,731 - 3,762) | 4,618 (3,851 - 5,657) | 7,980 (6,223 - 10,761) | 13,386 (9,343 - 21,230) |

**Supplementary Table S7: *E. coli* sequencing sample size estimates to reach different 'species richness' thresholds (the number of unique features detected as a proportion of the unique features in the NEKSUS sample) obtained from Bayesian modelling of frequency distributions combined with power calculation of different genomic features at 90, 95, and 99% certainty.**

| Feature \ Species richness | 30% | 40% | 50% | 60% | 70% | 80% | 90% | 95% |
|---|---|---|---|---|---|---|---|---|
| MLST 90% | 722 (678 - 774) | 903 (844 - 970) | 1,122 (1,039 - 1,218) | 1,425 (1,299 - 1,567) | 1,884 (1,689 - 2,114) | 2,688 (2,337 - 3,143) | 4,891 (3,949 - 6,262) | 8,782 (6,325 - 13,085) |
| MLST 95% | 940 (882 - 1,007) | 1,176 (1,098 - 1,262) | 1,460 (1,353 - 1,584) | 1,853 (1,691 - 2,038) | 2,451 (2,198 - 2,750) | 3,497 (3,040 - 4,089) | 6,364 (5,139 - 8,147) | 11,426 (8,229 - 17,023) |
| MLST 99% | 1,445 (1,356 - 1,549) | 1,807 (1,688 - 1,941) | 2,244 (2,079 - 2,436) | 2,850 (2,599 - 3,134) | 3,768 (3,379 - 4,228) | 5,376 (4,674 - 6,286) | 9,783 (7,899 - 12,525) | 17,564 (12,650 - 26,170) |
| fastBAPS L3 90% | 106 (83 - 137) | 148 (114 - 199) | 217 (159 - 309) | 323 (229 - 482) | 474 (323 - 748) | 703 (449 - 1,229) | 1,162 (660 - 2,452) | 2,060 (947 - 6,969) |
| fastBAPS L3 95% | 137 (107 - 178) | 192 (148 - 259) | 282 (206 - 402) | 420 (298 - 627) | 616 (420 - 973) | 914 (584 - 1,599) | 1,513 (859 - 3,191) | 2,680 (1,232 - 9,068) |
| fastBAPS L3 99% | 212 (166 - 275) | 296 (228 - 399) | 435 (317 - 618) | 647 (459 - 964) | 948 (646 - 1,496) | 1,406 (898 - 2,458) | 2,325 (1,320 - 4,905) | 4,121 (1,894 - 13,939) |
| Plasmids 90% | 640 (602 - 678) | 840 (789 - 897) | 1,110 (1,039 - 1,191) | 1,502 (1,394 - 1,624) | 2,150 (1,969 - 2,357) | 3,420 (3,064 - 3,833) | 7,243 (6,183 - 8,526) | 14,888 (11,878 - 18,795) |
| Plasmids 95% | 832 (783 - 883) | 1,093 (1,027 - 1,166) | 1,445 (1,352 - 1,550) | 1,954 (1,813 - 2,113) | 2,797 (2,562 - 3,067) | 4,449 (3,987 - 4,987) | 9,423 (8,044 - 11,092) | 19,370 (15,453 - 24,453) |
| Plasmids 99% | 1,279 (1,203 - 1,356) | 1,679 (1,578 - 1,793) | 2,220 (2,078 - 2,382) | 3,003 (2,787 - 3,247) | 4,299 (3,938 - 4,714) | 6,839 (6,128 - 7,666) | 14,485 (12,366 - 17,051) | 29,775 (23,756 - 37,590) |
| ARGs 90% | 178 (155 - 203) | 260 (230 - 296) | 353 (310 - 402) | 471 (411 - 542) | 646 (553 - 763) | 956 (790 - 1,176) | 1,776 (1,342 - 2,454) | 3,356 (2,217 - 5,698) |
| ARGs 95% | 231 (202 - 264) | 338 (298 - 385) | 458 (404 - 523) | 613 (535 - 705) | 840 (719 - 992) | 1,243 (1,028 - 1,529) | 2,310 (1,746 - 3,193) | 4,366 (2,884 - 7,413) |
| ARGs 99% | 354 (309 - 406) | 520 (459 - 592) | 705 (620 - 803) | 941 (822 - 1,084) | 1,291 (1,105 - 1,525) | 1,911 (1,580 - 2,351) | 3,551 (2,684 - 4,908) | 6,711 (4,434 - 11,395) |

**Supplementary Table S8: *Klebsiella* sequencing sample size estimates to reach different 'species richness' thresholds (the number of unique features detected as a proportion of the unique features in the NEKSUS sample) obtained from Bayesian modelling of frequency distributions combined with power calculation of different genomic features at 90, 95, and 99% certainty.**

| Region \ Coverage | 75% | 80% | 85% | 90% | 95% | 99% |
|---|---|---|---|---|---|---|
| **MLST** | | | | | | |
| East (95% cert.) | 1,111 (984 - 1,442) | 1,398 (1,270 - 2,175) | 1,780 (1,608 - 3,688) | 2,362 (2,115 - 8,054) | 3,585 (2,977 - 36,442) | 8,789 (6,054 - >100,000) |
| London (95% cert.) | 1,096 (976 - 1,351) | 1,385 (1,257 - 1,950) | 1,751 (1,578 - 3,057) | 2,312 (2,081 - 6,590) | 3,453 (3,012 - 19,926) | 7,947 (5,952 - >100,000) |
| Midlands (95% cert.) | 1,095 (975 - 1,237) | 1,370 (1,251 - 1,688) | 1,732 (1,579 - 2,403) | 2,284 (2,067 - 4,215) | 3,383 (2,975 - 11,895) | 7,429 (5,893 - >100,000) |
| North East A (95% cert.) | 1,352 (1,037 - 1,999) | 1,981 (1,415 - 3,291) | 3,155 (1,893 - 6,439) | 6,067 (2,737 - 19,519) | 20,157 (4,512 - >100,000) | >100,000 (17,825 - >100,000) |
| North East B (95% cert.) | 1,132 (985 - 1,558) | 1,428 (1,253 - 2,443) | 1,843 (1,596 - 4,190) | 2,539 (2,102 - 9,345) | 4,203 (3,019 - 45,426) | 13,173 (5,945 - >100,000) |
| North West (95% cert.) | 4,204 (1,185 - >100,000) | 10,467 (1,719 - >100,000) | 39,591 (2,731 - >100,000) | >100,000 (4,775 - >100,000) | >100,000 (12,542 - >100,000) | >100,000 |
| South East A (95% cert.) | 1,101 (983 - 1,344) | 1,383 (1,248 - 1,802) | 1,744 (1,575 - 2,747) | 2,299 (2,091 - 4,643) | 3,414 (2,979 - 13,925) | 7,543 (5,797 - >100,000) |
| South East B (95% cert.) | 1,141 (989 - 28,604) | 1,456 (1,256 - >100,000) | 1,930 (1,598 - >100,000) | 2,783 (2,102 - >100,000) | 5,218 (2,997 - >100,000) | 24,922 (6,033 - >100,000) |
| South East C (95% cert.) | 1,354 (998 - 3,939) | 2,028 (1,306 - 8,717) | 3,343 (1,646 - 32,471) | 6,662 (2,132 - >100,000) | 24,235 (3,096 - >100,000) | >100,000 (6,353 - >100,000) |
| South West (95% cert.) | 1,152 (981 - 1,513) | 1,465 (1,265 - 2,331) | 1,974 (1,606 - 3,739) | 2,822 (2,119 - 7,995) | 5,153 (3,052 - 26,958) | 20,606 (6,119 - >100,000) |
| National (95% cert.) | 1,092 (983 - 1,195) | 1,359 (1,233 - 1,459) | 1,693 (1,566 - 1,813) | 2,204 (2,028 - 2,364) | 3,172 (2,900 - 3,446) | 6,497 (5,684 - 7,465) |
| **fastBAPS cluster** | | | | | | |
| East (95% cert.) | 540 (468 - 635) | 667 (581 - 798) | 853 (749 - 1,095) | 1,161 (1,005 - 1,667) | 1,815 (1,573 - 3,643) | 4,261 (3,262 - 31,238) |
| London (95% cert.) | 535 (468 - 639) | 670 (579 - 825) | 857 (751 - 1,147) | 1,161 (1,014 - 1,724) | 1,830 (1,583 - 3,641) | 4,171 (3,254 - 38,244) |
| Midlands (95% cert.) | 551 (476 - 762) | 689 (590 - 1,107) | 883 (757 - 1,765) | 1,218 (1,023 - 3,361) | 1,964 (1,596 - 13,899) | 5,565 (3,197 - >100,000) |
| North East A (95% cert.) | 620 (499 - 799) | 800 (637 - 1,073) | 1,102 (818 - 1,612) | 1,685 (1,132 - 3,204) | 3,589 (1,809 - 11,433) | 29,817 (3,983 - >100,000) |
| North East B (95% cert.) | 552 (478 - 710) | 687 (585 - 929) | 890 (747 - 1,400) | 1,268 (1,017 - 2,469) | 2,112 (1,612 - 8,080) | 6,495 (3,417 - >100,000) |
| North West (95% cert.) | 565 (479 - 932) | 713 (583 - 1,336) | 942 (771 - 2,287) | 1,356 (1,040 - 5,691) | 2,530 (1,625 - 41,881) | 10,968 (3,380 - >100,000) |
| South East A (95% cert.) | 542 (468 - 649) | 672 (580 - 867) | 863 (752 - 1,179) | 1,170 (1,013 - 1,887) | 1,843 (1,573 - 3,857) | 4,455 (3,289 - 45,667) |
| South East B (95% cert.) | 557 (472 - 1,446) | 697 (597 - 2,580) | 888 (768 - 6,538) | 1,239 (1,013 - 25,107) | 2,000 (1,575 - >100,000) | 5,897 (3,299 - >100,000) |
| South East C (95% cert.) | 550 (473 - 751) | 689 (579 - 1,079) | 894 (754 - 1,723) | 1,233 (1,043 - 3,668) | 1,998 (1,578 - 14,370) | 5,274 (3,278 - >100,000) |
| South West (95% cert.) | 546 (475 - 645) | 685 (596 - 848) | 892 (767 - 1,205) | 1,254 (1,026 - 1,977) | 2,143 (1,588 - 4,933) | 6,964 (3,305 - >100,000) |
| National (95% cert.) | 531 (463 - 604) | 655 (569 - 736) | 833 (738 - 925) | 1,124 (1,004 - 1,244) | 1,729 (1,527 - 1,929) | 3,729 (3,079 - 4,457) |
| **Plasmid subcommunities** | | | | | | |
| East (95% cert.) | >100,000 | >100,000 | >100,000 | >100,000 | >100,000 | >100,000 |
| London (95% cert.) | >100,000 | >100,000 | >100,000 | >100,000 | >100,000 | >100,000 |
| Midlands (95% cert.) | >100,000 | >100,000 | >100,000 | >100,000 | >100,000 | >100,000 |
| North East A (95% cert.) | >100,000 | >100,000 | >100,000 | >100,000 | >100,000 | >100,000 |
| North East B (95% cert.) | >100,000 | >100,000 | >100,000 | >100,000 | >100,000 | >100,000 |
| North West (95% cert.) | >100,000 | >100,000 | >100,000 | >100,000 | >100,000 | >100,000 |
| South East A (95% cert.) | >100,000 | >100,000 | >100,000 | >100,000 | >100,000 | >100,000 |
| South East B (95% cert.) | >100,000 | >100,000 | >100,000 | >100,000 | >100,000 | >100,000 |
| South East C (95% cert.) | >100,000 | >100,000 | >100,000 | >100,000 | >100,000 | >100,000 |
| South West (95% cert.) | >100,000 | >100,000 | >100,000 | >100,000 | >100,000 | >100,000 |
| National (95% cert.) | 269 (252 - 288) | 313 (292 - 336) | 375 (348 - 403) | 477 (442 - 510) | 700 (649 - 757) | 1,604 (1,457 - 1,752) |
| **ARGs** | | | | | | |
| East (95% cert.) | 21 (19 - 24) | 28 (23 - 34) | 51 (39 - 66) | 105 (85 - 129) | 324 (237 - 460) | 3,131 (1,417 - 11,976) |
| London (95% cert.) | 21 (17 - 25) | 27 (23 - 35) | 47 (36 - 60) | 97 (73 - 125) | 379 (268 - 611) | 12,868 (2,074 - >100,000) |
| Midlands (95% cert.) | 23 (19 - 28) | 30 (25 - 38) | 56 (46 - 73) | 107 (87 - 133) | 300 (225 - 402) | 2,915 (1,376 - 13,521) |
| North East A (95% cert.) | 19 (17 - 22) | 27 (22 - 34) | 57 (46 - 70) | 105 (86 - 127) | 356 (256 - 510) | 4,777 (2,352 - 14,413) |
| North East B (95% cert.) | 21 (18 - 23) | 27 (22 - 32) | 43 (34 - 54) | 100 (77 - 127) | 343 (252 - 497) | 9,722 (3,472 - 51,320) |
| North West (95% cert.) | 21 (19 - 26) | 28 (23 - 36) | 54 (42 - 69) | 115 (89 - 151) | 343 (246 - 519) | 5,681 (1,563 - >100,000) |
| South East A (95% cert.) | 25 (21 - 31) | 34 (27 - 42) | 60 (46 - 78) | 125 (101 - 166) | 435 (305 - 649) | 14,621 (4,131 - >100,000) |
| South East B (95% cert.) | 23 (19 - 29) | 31 (25 - 40) | 52 (39 - 67) | 105 (82 - 144) | 360 (242 - 862) | 14,839 (1,733 - >100,000) |
| South East C (95% cert.) | 28 (23 - 36) | 40 (30 - 54) | 67 (50 - 91) | 139 (101 - 192) | 605 (350 - 1,350) | >100,000 (61,331 - >100,000) |
| South West (95% cert.) | 30 (25 - 38) | 40 (30 - 52) | 72 (57 - 94) | 133 (107 - 177) | 429 (291 - 657) | >100,000 (21,981 - >100,000) |
| National (95% cert.) | 21 (19 - 24) | 28 (24 - 34) | 52 (44 - 62) | 101 (85 - 116) | 280 (218 - 360) | 1,324 (1,182 - 1,491) |

**Supplementary Table S9: Regional *E. coli* sequencing sample size estimates to reach different coverage thresholds (the proportion of the national *E. coli* BSI population represented by a feature detected in the regional sample) at 95% certainty, derived from hierarchical Bayesian modelling combined with power calculation.** Higher regional estimates indicate the region is less representative of national diversity. National estimates are obtained directly from the hierarchical model, which estimates an overall (isolate-level) frequency for each feature (multi-locus sequence type [MLST], fastBAPS cluster, plasmid subcommunities, and antimicrobial resistance genes [ARGs]). Regional estimates are obtained by drawing from regional posterior frequency distributions, where regional distributions differ from the overall distribution by a region-specific $\tau$ parameter, which is also estimated by the model.

| Region \ Coverage | 75% | 80% | 85% | 90% | 95% | 99% |
|---|---|---|---|---|---|---|
| **MLST** | | | | | | |
| East (95% cert.) | 1,282 (1,174 - 1,748) | 1,470 (1,352 - 2,204) | 1,744 (1,573 - 2,886) | 2,174 (1,940 - 5,006) | 3,137 (2,688 - 13,304) | 7,088 (5,525 - >100,000) |
| London (95% cert.) | 15,756 (1,222 - >100,000) | 42,238 (1,394 - >100,000) | >100,000 (1,617 - >100,000) | >100,000 (2,018 - >100,000) | >100,000 (2,937 - >100,000) | >100,000 (6,287 - >100,000) |
| Midlands (95% cert.) | 1,350 (1,188 - 8,295) | 1,562 (1,359 - 16,797) | 1,902 (1,592 - 40,781) | 2,502 (1,947 - >100,000) | 3,872 (2,739 - >100,000) | 11,589 (5,616 - >100,000) |
| North East A (95% cert.) | 1,336 (1,184 - 3,119) | 1,551 (1,354 - 4,257) | 1,879 (1,595 - 7,342) | 2,419 (1,952 - 15,746) | 3,776 (2,719 - >100,000) | 10,601 (5,603 - >100,000) |
| North East B (95% cert.) | 2,288 (1,218 - 13,694) | 3,010 (1,389 - 30,253) | 4,384 (1,629 - >100,000) | 8,054 (2,012 - >100,000) | 24,034 (2,804 - >100,000) | >100,000 (5,918 - >100,000) |
| North West (95% cert.) | 1,744 (1,200 - >100,000) | 2,192 (1,367 - >100,000) | 2,973 (1,590 - >100,000) | 4,600 (1,964 - >100,000) | 10,288 (2,744 - >100,000) | >100,000 (5,795 - >100,000) |
| South East A (95% cert.) | 1,281 (1,183 - 1,776) | 1,474 (1,350 - 2,303) | 1,742 (1,581 - 3,142) | 2,171 (1,927 - 4,852) | 3,139 (2,691 - 10,773) | 7,081 (5,449 - >100,000) |
| South East B (95% cert.) | >100,000 (1,246 - >100,000) | >100,000 (1,428 - >100,000) | >100,000 (1,672 - >100,000) | >100,000 (2,066 - >100,000) | >100,000 (2,966 - >100,000) | >100,000 (6,711 - >100,000) |
| South East C (95% cert.) | 1,311 (1,182 - 3,954) | 1,512 (1,348 - 6,484) | 1,809 (1,598 - 13,759) | 2,312 (1,959 - 29,940) | 3,448 (2,755 - >100,000) | 9,423 (5,571 - >100,000) |
| South West (95% cert.) | 1,364 (1,192 - 10,864) | 1,642 (1,366 - 22,040) | 2,036 (1,604 - >100,000) | 2,659 (1,955 - >100,000) | 4,255 (2,699 - >100,000) | 13,772 (5,589 - >100,000) |
| National (95% cert.) | 1,242 (1,161 - 1,327) | 1,421 (1,333 - 1,518) | 1,666 (1,561 - 1,778) | 2,058 (1,903 - 2,205) | 2,888 (2,629 - 3,141) | 6,132 (5,253 - 7,046) |
| **fastBAPS cluster** | | | | | | |
| East (95% cert.) | 94 (63 - 177) | 125 (81 - 220) | 175 (107 - 352) | 270 (160 - 892) | 628 (300 - 9,163) | 11,759 (776 - >100,000) |
| London (95% cert.) | 176 (70 - 4,846) | 290 (94 - >100,000) | 624 (135 - >100,000) | 2,150 (204 - >100,000) | 31,346 (476 - >100,000) | >100,000 (4,052 - >100,000) |
| Midlands (95% cert.) | 91 (57 - 142) | 121 (73 - 204) | 162 (110 - 324) | 255 (153 - 704) | 535 (261 - 4,208) | 3,670 (790 - >100,000) |
| North East A (95% cert.) | 112 (67 - 239) | 149 (92 - 317) | 211 (127 - 566) | 326 (187 - 1,468) | 683 (300 - 6,994) | 5,354 (785 - >100,000) |
| North East B (95% cert.) | 91 (57 - 150) | 124 (81 - 242) | 180 (112 - 469) | 302 (157 - 1,189) | 682 (303 - 17,037) | 8,805 (823 - >100,000) |
| North West (95% cert.) | 101 (62 - 306) | 140 (87 - 598) | 198 (122 - 1,684) | 306 (164 - 6,811) | 689 (308 - >100,000) | 10,233 (790 - >100,000) |
| South East A (95% cert.) | 95 (58 - 148) | 125 (86 - 215) | 170 (116 - 334) | 275 (155 - 699) | 605 (284 - 4,602) | 4,767 (859 - >100,000) |
| South East B (95% cert.) | 106 (60 - 23,084) | 151 (84 - >100,000) | 218 (120 - >100,000) | 389 (164 - >100,000) | 1,141 (318 - >100,000) | >100,000 (877 - >100,000) |
| South East C (95% cert.) | 97 (58 - 178) | 132 (85 - 391) | 192 (120 - 677) | 321 (164 - 3,155) | 790 (314 - >100,000) | 11,707 (717 - >100,000) |
| South West (95% cert.) | 95 (60 - 214) | 126 (77 - 295) | 174 (110 - 769) | 276 (154 - 2,152) | 826 (296 - 52,764) | 20,654 (913 - >100,000) |
| National (95% cert.) | 93 (64 - 123) | 118 (86 - 153) | 148 (110 - 190) | 206 (148 - 278) | 356 (254 - 477) | 917 (590 - 1,270) |
| **Plasmid subcommunities** | | | | | | |
| East (95% cert.) | >100,000 | >100,000 | >100,000 | >100,000 | >100,000 | >100,000 |
| London (95% cert.) | >100,000 | >100,000 | >100,000 | >100,000 | >100,000 | >100,000 |
| Midlands (95% cert.) | >100,000 | >100,000 | >100,000 | >100,000 | >100,000 | >100,000 |
| North East A (95% cert.) | >100,000 | >100,000 | >100,000 | >100,000 | >100,000 | >100,000 |
| North East B (95% cert.) | >100,000 | >100,000 | >100,000 | >100,000 | >100,000 | >100,000 |
| North West (95% cert.) | >100,000 | >100,000 | >100,000 | >100,000 | >100,000 | >100,000 |
| South East A (95% cert.) | >100,000 | >100,000 | >100,000 | >100,000 | >100,000 | >100,000 |
| South East B (95% cert.) | >100,000 | >100,000 | >100,000 | >100,000 | >100,000 | >100,000 |
| South East C (95% cert.) | >100,000 | >100,000 | >100,000 | >100,000 | >100,000 | >100,000 |
| South West (95% cert.) | >100,000 | >100,000 | >100,000 | >100,000 | >100,000 | >100,000 |
| National (95% cert.) | 209 (194 - 222) | 243 (226 - 259) | 294 (274 - 313) | 377 (351 - 403) | 562 (520 - 603) | 1,346 (1,222 - 1,475) |
| **ARGs** | | | | | | |
| East (95% cert.) | 70 (56 - 92) | 107 (84 - 143) | 179 (141 - 254) | 337 (243 - 589) | 876 (425 - 2,815) | 10,968 (1,324 - >100,000) |
| London (95% cert.) | 81 (56 - 117) | 143 (100 - 257) | 313 (177 - 857) | 1,044 (387 - 6,047) | 13,255 (1,088 - >100,000) | >100,000 (45,380 - >100,000) |
| Midlands (95% cert.) | 69 (59 - 83) | 98 (81 - 118) | 151 (125 - 188) | 246 (211 - 334) | 470 (368 - 909) | 1,829 (913 - 16,644) |
| North East A (95% cert.) | 81 (64 - 105) | 121 (93 - 162) | 205 (153 - 290) | 404 (281 - 628) | 1,291 (629 - 3,182) | 30,506 (3,200 - >100,000) |
| North East B (95% cert.) | 80 (64 - 105) | 125 (95 - 164) | 218 (162 - 303) | 473 (306 - 814) | 1,903 (764 - 7,433) | >100,000 (9,726 - >100,000) |
| North West (95% cert.) | 71 (58 - 88) | 101 (83 - 124) | 155 (129 - 203) | 256 (216 - 393) | 523 (376 - 1,445) | 2,639 (916 - 59,275) |
| South East A (95% cert.) | 103 (79 - 138) | 159 (118 - 230) | 286 (202 - 457) | 720 (420 - 1,398) | 4,309 (1,526 - 23,764) | >100,000 (51,053 - >100,000) |
| South East B (95% cert.) | 166 (72 - 1,172) | 546 (166 - 11,759) | 3,319 (445 - >100,000) | >100,000 (3,247 - >100,000) | >100,000 | >100,000 |
| South East C (95% cert.) | 77 (62 - 104) | 110 (86 - 155) | 174 (134 - 269) | 327 (228 - 665) | 961 (406 - 4,648) | 18,501 (1,171 - >100,000) |
| South West (95% cert.) | 116 (80 - 181) | 185 (120 - 309) | 390 (211 - 1,023) | 1,376 (507 - 8,756) | 22,703 (2,110 - >100,000) | >100,000 |
| National (95% cert.) | 67 (58 - 80) | 95 (80 - 110) | 149 (126 - 170) | 233 (209 - 263) | 389 (349 - 438) | 924 (806 - 1,067) |

**Supplementary Table S10: Regional *Klebsiella* sequencing sample size estimates to reach different coverage thresholds (the proportion of the national *Klebsiella* spp. BSI population represented by a feature detected in the regional sample) at 95% certainty, derived from hierarchical Bayesian modelling combined with power calculation.** Higher regional estimates indicate the region is less representative of national diversity. National estimates are obtained directly from the hierarchical model, which estimates an overall (isolate-level) frequency for each feature (multi-locus sequence type [MLST], fastBAPS cluster, plasmid subcommunities, and antimicrobial resistance genes [ARGs]). Regional estimates are obtained by drawing from regional posterior frequency distributions, where regional distributions differ from the overall distribution by a region-specific $\tau$ parameter, which is also estimated by the model.

| Region \ Species richness | 30% | 40% | 50% | 60% | 70% | 80% | 90% |
|---|---|---|---|---|---|---|---|
| **MLST** | | | | | | | |
| East (95% cert.) | 1,370 (1,201 - 1,556) | 1,773 (1,587 - 2,274) | 2,277 (2,031 - 3,907) | 2,908 (2,495 - 7,745) | 3,853 (3,189 - 19,438) | 5,518 (4,265 - >100,000) | 10,366 (6,789 - >100,000) |
| London (95% cert.) | 1,354 (1,185 - 1,526) | 1,754 (1,577 - 2,223) | 2,245 (2,013 - 3,500) | 2,862 (2,528 - 6,500) | 3,802 (3,230 - 13,596) | 5,346 (4,350 - 48,451) | 9,531 (6,971 - >100,000) |
| Midlands (95% cert.) | 1,362 (1,209 - 1,508) | 1,745 (1,581 - 1,988) | 2,224 (1,994 - 2,696) | 2,813 (2,483 - 4,657) | 3,688 (3,178 - 8,418) | 5,168 (4,274 - 19,574) | 8,903 (6,626 - >100,000) |
| North East A (95% cert.) | 1,424 (1,219 - 1,756) | 2,153 (1,725 - 2,980) | 3,409 (2,320 - 5,992) | 5,688 (3,114 - 14,832) | 11,889 (4,308 - 52,959) | 36,306 (6,932 - >100,000) | >100,000 (17,623 - >100,000) |
| North East B (95% cert.) | 1,382 (1,208 - 1,570) | 1,807 (1,605 - 2,521) | 2,340 (2,040 - 4,482) | 3,057 (2,524 - 8,887) | 4,189 (3,269 - 22,538) | 6,464 (4,319 - >100,000) | 13,330 (6,850 - >100,000) |
| North West (95% cert.) | 3,020 (1,298 - 28,018) | 8,955 (1,923 - >100,000) | 36,755 (3,012 - >100,000) | >100,000 (4,386 - [illegible] | >100,000 (9,072 - >100,000) | >100,000 (36,429 - [illegible] | >100,000 |
| South East A (95% cert.) | 1,371 (1,223 - 1,527) | 1,761 (1,598 - 2,079) | 2,244 (2,026 - 2,999) | 2,848 (2,504 - 4,948) | 3,744 (3,213 - 9,098) | 5,263 (4,167 - 24,567) | 9,407 (6,659 - >100,000) |
| South East B (95% cert.) | 1,370 (1,173 - 12,298) | 1,831 (1,589 - >100,000) | 2,398 (2,010 - >100,000) | 3,285 (2,514 - >100,000) | 4,867 (3,222 - >100,000) | 7,999 (4,274 - >100,000) | 22,362 (6,899 - >100,000) |
| South East C (95% cert.) | 1,468 (1,195 - 3,087) | 2,243 (1,659 - 8,457) | 3,629 (2,080 - 34,393) | 6,458 (2,569 - >100,000) | 14,979 (3,333 - >100,000) | 54,117 (4,582 - >100,000) | >100,000 (7,660 - >100,000) |
| South West (95% cert.) | 1,371 (1,210 - 1,531) | 1,815 (1,612 - 2,354) | 2,380 (2,052 - 3,806) | 3,204 (2,560 - 7,146) | 4,717 (3,274 - 15,509) | 7,703 (4,417 - 56,218) | 19,119 (6,873 - >100,000) |
| National (95% cert.) | 1,387 (1,257 - 1,516) | 1,748 (1,598 - 1,935) | 2,195 (1,976 - 2,420) | 2,726 (2,453 - 3,072) | 3,531 (3,105 - 3,994) | 4,809 (4,123 - 5,687) | 7,943 (6,439 - 9,983) |
| **fastBAPS cluster** | | | | | | | |
| East (95% cert.) | 520 (451 - 589) | 726 (642 - 830) | 1,004 (878 - 1,198) | 1,360 (1,196 - 1,739) | 1,917 (1,598 - 2,890) | 2,836 (2,224 - 6,441) | 5,156 (3,691 - 31,723) |
| London (95% cert.) | 516 (452 - 587) | 728 (643 - 831) | 1,007 (883 - 1,211) | 1,374 (1,181 - 1,770) | 1,928 (1,627 - 3,220) | 2,833 (2,262 - 6,915) | 5,168 (3,684 - 30,672) |
| Midlands (95% cert.) | 514 (446 - 588) | 735 (645 - 902) | 1,032 (890 - 1,486) | 1,421 (1,197 - 2,919) | 2,016 (1,645 - 6,935) | 3,135 (2,296 - 29,046) | 6,268 (3,856 - >100,000) |
| North East A (95% cert.) | 528 (458 - 607) | 767 (659 - 890) | 1,119 (937 - 1,462) | 1,648 (1,292 - 2,486) | 2,742 (1,876 - 5,242) | 5,601 (2,629 - 19,910) | 22,993 (4,801 - >100,000) |
| North East B (95% cert.) | 512 (446 - 585) | 730 (642 - 844) | 1,040 (877 - 1,294) | 1,456 (1,209 - 2,252) | 2,110 (1,702 - 4,596) | 3,282 (2,379 - 15,392) | 6,958 (3,797 - >100,000) |
| North West (95% cert.) | 512 (444 - 592) | 734 (628 - 980) | 1,056 (896 - 1,928) | 1,491 (1,200 - 4,181) | 2,252 (1,676 - 13,928) | 3,886 (2,311 - >100,000) | 11,128 (3,831 - >100,000) |
| South East A (95% cert.) | 516 (447 - 592) | 734 (648 - 846) | 1,015 (896 - 1,229) | 1,370 (1,168 - 1,793) | 1,957 (1,618 - 2,903) | 2,891 (2,269 - 5,860) | 5,255 (3,736 - 29,648) |
| South East B (95% cert.) | 519 (451 - 704) | 743 (642 - 1,406) | 1,035 (898 - 3,798) | 1,431 (1,195 - 15,398) | 2,052 (1,631 - >100,000) | 3,180 (2,247 - >100,000) | 6,342 (3,853 - >100,000) |
| South East C (95% cert.) | 512 (444 - 591) | 736 (643 - 881) | 1,032 (893 - 1,454) | 1,419 (1,194 - 2,774) | 2,072 (1,644 - 6,191) | 3,170 (2,275 - 24,876) | 6,199 (3,718 - >100,000) |
| South West (95% cert.) | 509 (447 - 579) | 719 (628 - 838) | 1,020 (865 - 1,201) | 1,415 (1,211 - 1,855) | 2,090 (1,679 - 3,525) | 3,229 (2,286 - 8,376) | 7,013 (3,772 - >100,000) |
| National (95% cert.) | 521 (464 - 593) | 735 (650 - 818) | 1,006 (884 - 1,138) | 1,344 (1,172 - 1,584) | 1,853 (1,601 - 2,225) | 2,632 (2,197 - 3,312) | 4,506 (3,502 - 6,056) |
| **Plasmid subcommunities** | | | | | | | |
| East (95% cert.) | >100,000 | >100,000 | >100,000 | >100,000 | >100,000 | >100,000 | >100,000 |
| London (95% cert.) | >100,000 | >100,000 | >100,000 | >100,000 | >100,000 | >100,000 | >100,000 |
| Midlands (95% cert.) | >100,000 | >100,000 | >100,000 | >100,000 | >100,000 | >100,000 | >100,000 |
| North East A (95% cert.) | 47,683 (23,029 - >100,000) | >100,000 | >100,000 | >100,000 | >100,000 | >100,000 | >100,000 |
| North East B (95% cert.) | >100,000 | >100,000 | >100,000 | >100,000 | >100,000 | >100,000 | >100,000 |
| North West (95% cert.) | >100,000 | >100,000 | >100,000 | >100,000 | >100,000 | >100,000 | >100,000 |
| South East A (95% cert.) | >100,000 | >100,000 | >100,000 | >100,000 | >100,000 | >100,000 | >100,000 |
| South East B (95% cert.) | >100,000 | >100,000 | >100,000 | >100,000 | >100,000 | >100,000 | >100,000 |
| South East C (95% cert.) | >100,000 | >100,000 | >100,000 | >100,000 | >100,000 | >100,000 | >100,000 |
| South West (95% cert.) | >100,000 | >100,000 | >100,000 | >100,000 | >100,000 | >100,000 | >100,000 |
| National (95% cert.) | 224 (209 - 239) | 287 (267 - 311) | 373 (343 - 403) | 491 (449 - 535) | 682 (622 - 749) | 1,043 (941 - 1,178) | 2,115 (1,794 - 2,477) |
| **ARGs** | | | | | | | |
| East (95% cert.) | 282 (230 - 347) | 559 (461 - 707) | 890 (707 - 1,257) | 1,404 (1,032 - 2,489) | 2,475 (1,469 - 6,876) | 5,827 (2,280 - 39,038) | 31,420 (4,412 - >100,000) |
| London (95% cert.) | 290 (219 - 389) | 633 (467 - 995) | 1,238 (802 - 2,823) | 2,694 (1,254 - 10,763) | 7,894 (1,975 - >100,000) | 43,455 (3,361 - >100,000) | >100,000 (10,454 - >100,000) |
| Midlands (95% cert.) | 265 (215 - 339) | 519 (419 - 672) | 844 (672 - 1,139) | 1,343 (1,004 - 2,486) | 2,325 (1,421 - 6,574) | 5,409 (2,169 - 35,493) | 25,406 (3,852 - >100,000) |
| North East A (95% cert.) | 302 (242 - 370) | 617 (515 - 785) | 1,013 (803 - 1,303) | 1,704 (1,229 - 2,694) | 3,206 (2,024 - 6,571) | 8,678 (3,575 - 36,585) | >100,000 (11,466 - >100,000) |
| North East B (95% cert.) | 265 (213 - 324) | 556 (460 - 752) | 1,057 (779 - 1,543) | 2,174 (1,337 - 4,350) | 5,486 (2,557 - 18,586) | 25,312 (6,668 - >100,000) | >100,000 (39,168 - >100,000) |
| North West (95% cert.) | 276 (219 - 349) | 548 (438 - 742) | 950 (710 - 1,730) | 1,815 (1,058 - 5,386) | 3,992 (1,538 - 27,306) | 13,069 (2,439 - >100,000) | >100,000 (5,492 - >100,000) |
| South East A (95% cert.) | 320 (258 - 410) | 648 (523 - 846) | 1,172 (879 - 1,662) | 2,448 (1,530 - 4,614) | 6,698 (2,906 - 19,994) | 37,008 (7,270 - >100,000) | >100,000 |
| South East B (95% cert.) | 255 (202 - 347) | 562 (419 - 1,017) | 1,111 (708 - 4,690) | 2,562 (1,067 - 28,197) | 8,090 (1,673 - >100,000) | 50,110 (2,986 - >100,000) | >100,000 (7,153 - >100,000) |
| South East C (95% cert.) | 337 (245 - 488) | 1,141 (671 - 2,417) | 4,548 (1,657 - 30,708) | 26,525 (5,093 - >100,000) | >100,000 (25,595 - >100,000) | >100,000 | >100,000 |
| South West (95% cert.) | 283 (230 - 360) | 738 (556 - 1,094) | 2,059 (1,241 - 4,143) | 7,904 (3,500 - 29,391) | 51,957 (12,172 - >100,000) | >100,000 | >100,000 |
| National (95% cert.) | 272 (226 - 334) | 564 (483 - 690) | 804 (697 - 939) | 1,089 (937 - 1,270) | 1,479 (1,255 - 1,762) | 2,125 (1,767 - 2,627) | 3,626 (2,860 - 4,903) |

**Supplementary Table S11: Regional *E. coli* sequencing sample size estimates to reach different thresholds of species richness (the proportion of the unique MLST, fastBAPS clusters, plasmid subcommunities, and antimicrobial resistance genes (ARGs) detected in the NEKSUS dataset) at 95% certainty, derived from hierarchical Bayesian modelling combined with power calculation.** Higher regional estimates indicate the region is less representative of national diversity. National estimates are obtained directly from the hierarchical model, which estimates an overall (isolate-level) frequency for each feature. Regional estimates are obtained by drawing from regional posterior frequency distributions, where regional distributions differ from the overall distribution by a region-specific $\tau$ parameter, which is also estimated by the model.

| Region \ Species Richness | 30% | 40% | 50% | 60% | 70% | 80% | 90% |
|---|---|---|---|---|---|---|---|
| **MLST** | | | | | | | |
| East (95% cert.) | 928 (863 - 1,005) | 1,175 (1,092 - 1,283) | 1,478 (1,354 - 1,769) | 1,891 (1,672 - 2,615) | 2,544 (2,236 - 4,321) | 3,748 (3,117 - 9,869) | 7,244 (5,367 - >100,000) |
| London (95% cert.) | 1,552 (889 - 30,136) | 3,480 (1,115 - >100,000) | 9,521 (1,395 - >100,000) | 37,821 (1,777 - >100,000) | >100,000 (2,396 - >100,000) | >100,000 (3,427 - >100,000) | >100,000 (6,284 - >100,000) |
| Midlands (95% cert.) | 933 (854 - 1,273) | 1,200 (1,108 - 2,398) | 1,529 (1,361 - 5,618) | 1,992 (1,715 - 18,543) | 2,753 (2,259 - >100,000) | 4,195 (3,212 - >100,000) | 9,179 (5,547 - >100,000) |
| North East A (95% cert.) | 929 (865 - 1,013) | 1,188 (1,096 - 1,515) | 1,511 (1,360 - 2,420) | 1,964 (1,724 - 4,151) | 2,683 (2,251 - 10,770) | 4,141 (3,152 - 40,137) | 9,091 (5,382 - >100,000) |
| North East B (95% cert.) | 969 (869 - 1,438) | 1,370 (1,123 - 3,257) | 2,061 (1,395 - 8,317) | 3,431 (1,764 - 30,383) | 6,919 (2,315 - >100,000) | 21,211 (3,260 - >100,000) | >100,000 (5,870 - >100,000) |
| North West (95% cert.) | 950 (865 - 2,980) | 1,272 (1,106 - 12,455) | 1,742 (1,362 - >100,000) | 2,541 (1,720 - >100,000) | 4,349 (2,279 - >100,000) | 9,186 (3,169 - >100,000) | 46,922 (5,696 - >100,000) |
| South East A (95% cert.) | 926 (859 - 994) | 1,177 (1,091 - 1,291) | 1,478 (1,358 - 1,776) | 1,893 (1,694 - 2,575) | 2,536 (2,214 - 4,099) | 3,749 (3,024 - 8,926) | 7,231 (5,229 - 48,165) |
| South East B (95% cert.) | >100,000 (898 - >100,000) | >100,000 (1,145 - >100,000) | >100,000 (1,436 - >100,000) | >100,000 (1,837 - >100,000) | >100,000 (2,400 - >100,000) | >100,000 (3,557 - >100,000) | >100,000 (6,941 - >100,000) |
| South East C (95% cert.) | 930 (861 - 1,060) | 1,181 (1,089 - 1,786) | 1,497 (1,347 - 2,973) | 1,941 (1,730 - 6,515) | 2,643 (2,266 - 18,765) | 3,999 (3,124 - >100,000) | 8,477 (5,315 - >100,000) |
| South West (95% cert.) | 936 (857 - 1,324) | 1,195 (1,098 - 2,688) | 1,544 (1,364 - 6,299) | 2,037 (1,720 - 20,622) | 2,890 (2,240 - >100,000) | 4,547 (3,135 - >100,000) | 10,464 (5,483 - >100,000) |
| National (95% cert.) | 935 (872 - 999) | 1,170 (1,089 - 1,253) | 1,449 (1,346 - 1,581) | 1,840 (1,672 - 2,018) | 2,428 (2,181 - 2,715) | 3,457 (3,006 - 4,056) | 6,327 (5,096 - 8,021) |
| **fastBAPS cluster** | | | | | | | |
| East (95% cert.) | 127 (87 - 189) | 187 (129 - 312) | 300 (187 - 668) | 512 (290 - 2,704) | 1,011 (446 - 27,761) | 3,090 (695 - >100,000) | 21,949 (1,115 - >100,000) |
| London (95% cert.) | 186 (95 - 4,478) | 451 (140 - >100,000) | 1,489 (214 - >100,000) | 8,597 (363 - >100,000) | >100,000 (622 - >100,000) | >100,000 (1,205 - >100,000) | >100,000 (4,156 - >100,000) |
| Midlands (95% cert.) | 128 (92 - 207) | 187 (131 - 308) | 289 (177 - 554) | 449 (272 - 1,566) | 747 (395 - 7,481) | 1,462 (564 - >100,000) | 5,237 (909 - >100,000) |
| North East A (95% cert.) | 153 (108 - 236) | 226 (157 - 365) | 331 (217 - 650) | 483 (308 - 1,508) | 797 (436 - 3,534) | 1,562 (605 - 30,630) | 6,153 (1,056 - >100,000) |
| North East B (95% cert.) | 134 (95 - 199) | 203 (142 - 393) | 331 (207 - 1,189) | 540 (308 - 3,401) | 1,028 (447 - 21,464) | 2,686 (641 - >100,000) | 15,031 (1,155 - >100,000) |
| North West (95% cert.) | 136 (93 - 271) | 208 (139 - 605) | 311 (193 - 2,435) | 489 (264 - 17,695) | 894 (385 - >100,000) | 2,109 (586 - >100,000) | 13,551 (1,048 - >100,000) |
| South East A (95% cert.) | 130 (97 - 183) | 194 (137 - 317) | 299 (200 - 543) | 471 (287 - 1,750) | 880 (410 - 8,275) | 1,890 (656 - >100,000) | 8,483 (1,053 - >100,000) |
| South East B (95% cert.) | 133 (86 - 25,621) | 213 (133 - >100,000) | 362 (192 - >100,000) | 721 (278 - >100,000) | 2,123 (438 - >100,000) | 10,711 (667 - >100,000) | >100,000 (1,072 - >100,000) |
| South East C (95% cert.) | 139 (93 - 248) | 216 (129 - 525) | 330 (193 - 1,350) | 547 (286 - 5,946) | 998 (414 - 52,087) | 2,694 (607 - >100,000) | 15,239 (989 - >100,000) |
| South West (95% cert.) | 121 (84 - 187) | 188 (122 - 336) | 297 (182 - 861) | 539 (271 - 5,646) | 1,210 (427 - 62,964) | 4,379 (660 - >100,000) | 62,261 (1,253 - >100,000) |
| National (95% cert.) | 132 (99 - 178) | 181 (136 - 261) | 257 (188 - 367) | 374 (255 - 594) | 541 (343 - 909) | 788 (489 - 1,375) | 1,347 (747 - 3,013) |
| **Plasmid subcommunities** | | | | | | | |
| East (95% cert.) | >100,000 | >100,000 | >100,000 | >100,000 | >100,000 | >100,000 | >100,000 |
| London (95% cert.) | >100,000 | >100,000 | >100,000 | >100,000 | >100,000 | >100,000 | >100,000 |
| Midlands (95% cert.) | >100,000 | >100,000 | >100,000 | >100,000 | >100,000 | >100,000 | >100,000 |
| North East A (95% cert.) | >100,000 | >100,000 | >100,000 | >100,000 | >100,000 | >100,000 | >100,000 |
| North East B (95% cert.) | >100,000 | >100,000 | >100,000 | >100,000 | >100,000 | >100,000 | >100,000 |
| North West (95% cert.) | >100,000 | >100,000 | >100,000 | >100,000 | >100,000 | >100,000 | >100,000 |
| South East A (95% cert.) | >100,000 | >100,000 | >100,000 | >100,000 | >100,000 | >100,000 | >100,000 |
| South East B (95% cert.) | >100,000 | >100,000 | >100,000 | >100,000 | >100,000 | >100,000 | >100,000 |
| South East C (95% cert.) | >100,000 | >100,000 | >100,000 | >100,000 | >100,000 | >100,000 | >100,000 |
| South West (95% cert.) | >100,000 | >100,000 | >100,000 | >100,000 | >100,000 | >100,000 | >100,000 |
| National (95% cert.) | 168 (159 - 179) | 220 (206 - 235) | 291 (270 - 312) | 391 (363 - 425) | 561 (507 - 614) | 894 (794 - 1,003) | 1,880 (1,598 - 2,233) |
| **ARGs** | | | | | | | |
| East (95% cert.) | 191 (158 - 239) | 304 (244 - 439) | 480 (334 - 859) | 799 (463 - 1,950) | 1,523 (680 - 7,345) | 4,136 (1,063 - >100,000) | 44,535 (2,558 - >100,000) |
| London (95% cert.) | 240 (166 - 444) | 561 (281 - 1,778) | 1,643 (499 - 13,209) | 6,413 (942 - >100,000) | 55,190 (2,276 - >100,000) | >100,000 (9,622 - >100,000) | >100,000 |
| Midlands (95% cert.) | 172 (147 - 207) | 254 (220 - 309) | 360 (308 - 460) | 512 (417 - 775) | 765 (559 - 1,606) | 1,347 (814 - 5,485) | 4,055 (1,521 - >100,000) |
| North East A (95% cert.) | 209 (174 - 261) | 350 (271 - 474) | 585 (396 - 890) | 1,044 (631 - 2,274) | 2,381 (1,021 - 9,039) | 8,464 (2,087 - >100,000) | >100,000 (9,901 - >100,000) |
| North East B (95% cert.) | 205 (170 - 259) | 366 (274 - 534) | 691 (441 - 1,318) | 1,495 (722 - 4,816) | 4,668 (1,333 - 33,970) | 30,744 (3,457 - >100,000) | >100,000 (33,743 - >100,000) |
| North West (95% cert.) | 173 (144 - 208) | 257 (216 - 323) | 375 (300 - 562) | 539 (417 - 1,117) | 845 (567 - 3,199) | 1,654 (859 - 16,032) | 6,642 (1,585 - >100,000) |
| South East A (95% cert.) | 251 (202 - 335) | 477 (343 - 736) | 1,028 (589 - 2,044) | 2,643 (1,132 - 9,693) | 11,593 (2,890 - >100,000) | >100,000 (13,180 - >100,000) | >100,000 |
| South East B (95% cert.) | 583 (202 - 7,205) | 6,136 (801 - >100,000) | >100,000 (5,606 - >100,000) | >100,000 (41,354 - >100,000) | >100,000 | >100,000 | >100,000 |
| South East C (95% cert.) | 176 (148 - 222) | 284 (222 - 423) | 464 (324 - 933) | 816 (439 - 2,499) | 1,784 (635 - 12,718) | 5,675 (1,028 - >100,000) | >100,000 (2,424 - >100,000) |
| South West (95% cert.) | 280 (182 - 488) | 717 (369 - 1,882) | 2,304 (790 - 18,787) | 11,765 (1,758 - >100,000) | >100,000 (5,970 - >100,000) | >100,000 (49,680 - >100,000) | >100,000 |
| National (95% cert.) | 182 (156 - 211) | 260 (225 - 304) | 349 (299 - 405) | 462 (391 - 544) | 630 (520 - 757) | 922 (756 - 1,132) | 1,700 (1,287 - 2,326) |

**Supplementary Table S12: Regional *Klebsiella* sequencing sample size estimates to reach different thresholds of species richness (the proportion of the unique MLST, fastBAPS clusters, plasmid subcommunities, and antimicrobial resistance genes (ARGs) detected in the NEKSUS dataset) at 95% certainty, derived from hierarchical Bayesian modelling combined with power calculation.** Higher regional estimates indicate the region is less representative of national diversity. National estimates are obtained directly from the hierarchical model, which estimates an overall (isolate-level) frequency for each feature. Regional estimates are obtained by drawing from regional posterior frequency distributions, where regional distributions differ from the overall distribution by a region-specific $\tau$ parameter, which is also estimated by the model.

### Supplementary Figures

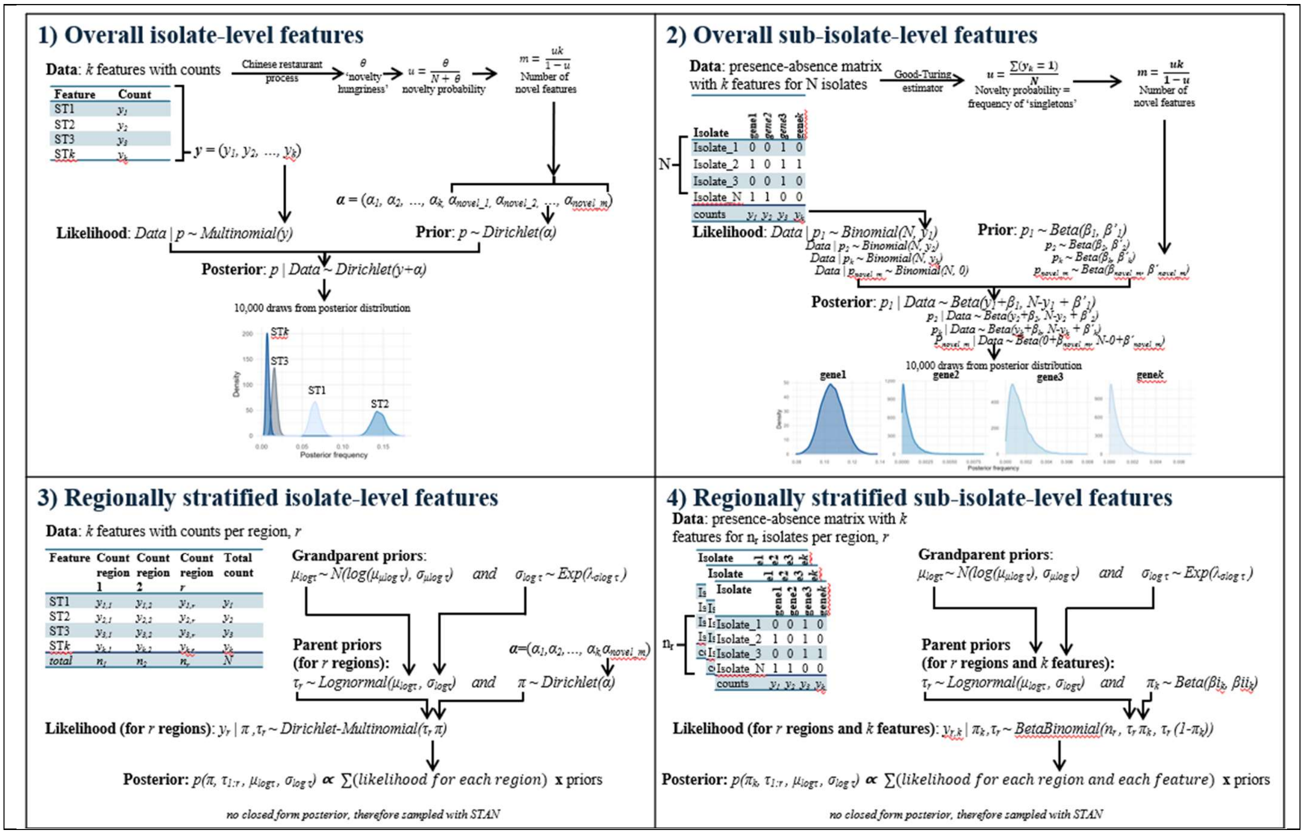

**Supplementary Figure S1: Four Bayesian modelling frameworks used to estimate the posterior predictive distributions of genomic features.** Frameworks 1-2) show standard (overall) Bayesian estimation of overall national feature counts, while 3-4) show regionally-stratified counts. Frameworks 1) and 3) model isolate-level features using a Dirichlet-Multinomial framework, where features are mututally-exclusive, and therefore their frequencies add up to 1. Frameworks 2) and 4) model sub-isolate-level features using separate Beta-Binomial distributions for each feature, thus allowing frequencies to sum >1 for these non-mututally-exclusive features.

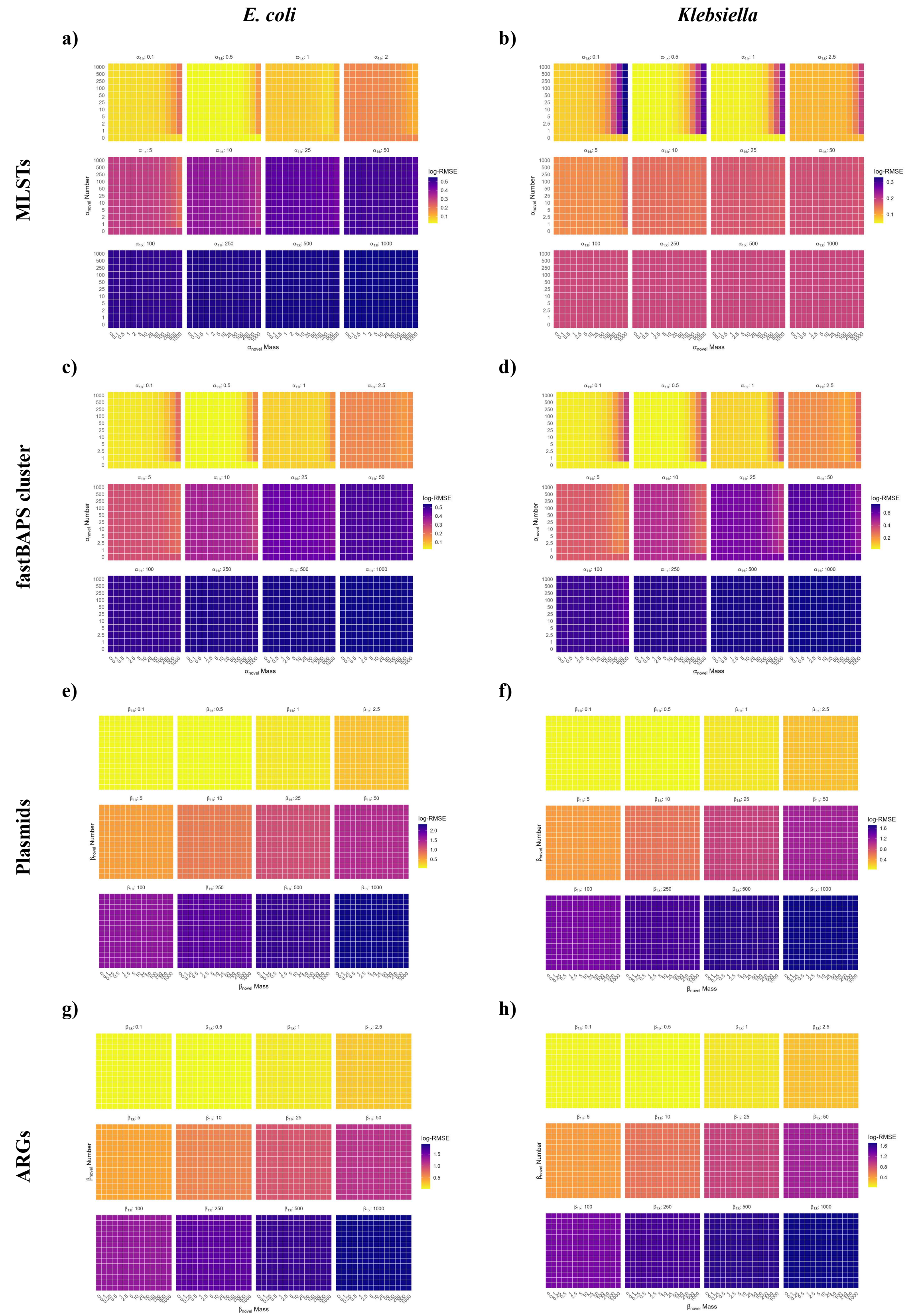

**Supplementary Figure S2: Heatmaps showing goodness of fit of prior parameter combinations for Bayesian modelling of *E. coli* (left column; a,c,e,g) and *Klebsiella* (right column; b,d,f,h) genomic features**. Goodness of fit of Bayesian posterior estimates to observed genomic feature frequencies were measured by root mean squared error. Genomic features represented are, from top to bottom: MLSTs (**a-b)**), fastBAPS clusters (**c-d)**), plasmids subcommunities (**e-f)**), and antimicrobial resistance genes (ARGs) (**g-h)**).

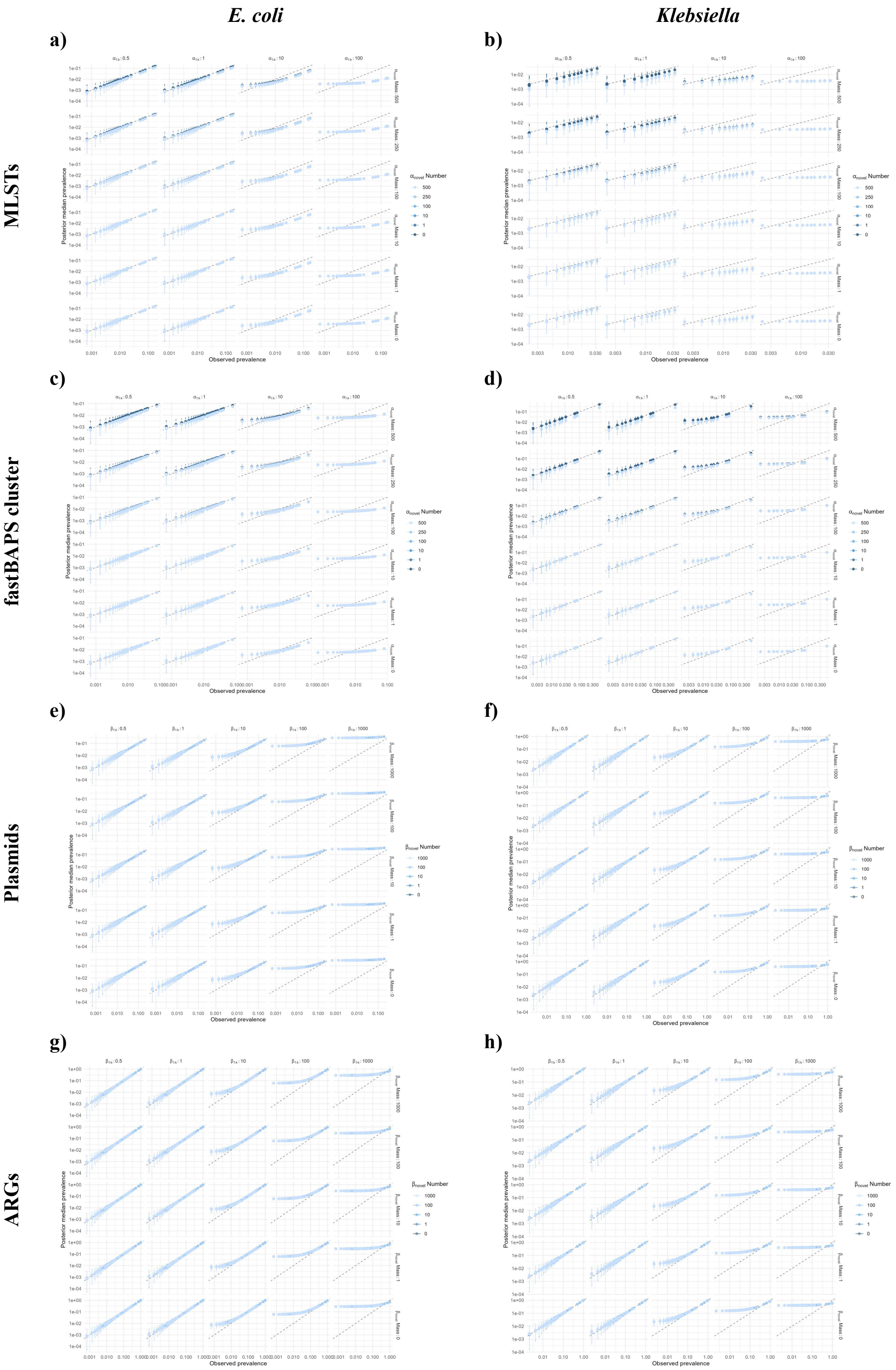

**Supplementary Figure S3: Panel plots showing observed (x-axis) vs predicted feature frequencies from Bayesian modelling (y-axis) for different prior parameter combinations for *E. coli* (left column; a,c,e,g) and *Klebsiella* (right column; b,d,f,h).** Points represent the median posterior estimate, and vertical error bars represent the 95% credible intervals (CrIs). Genomic features represented are, from top to bottom: MLSTs (**a-b**)), fastBAPS clusters (**c-d**)), plasmids subcommunities (**e-f**)), and antimicrobial resistance genes (ARGs; **g-h**)).

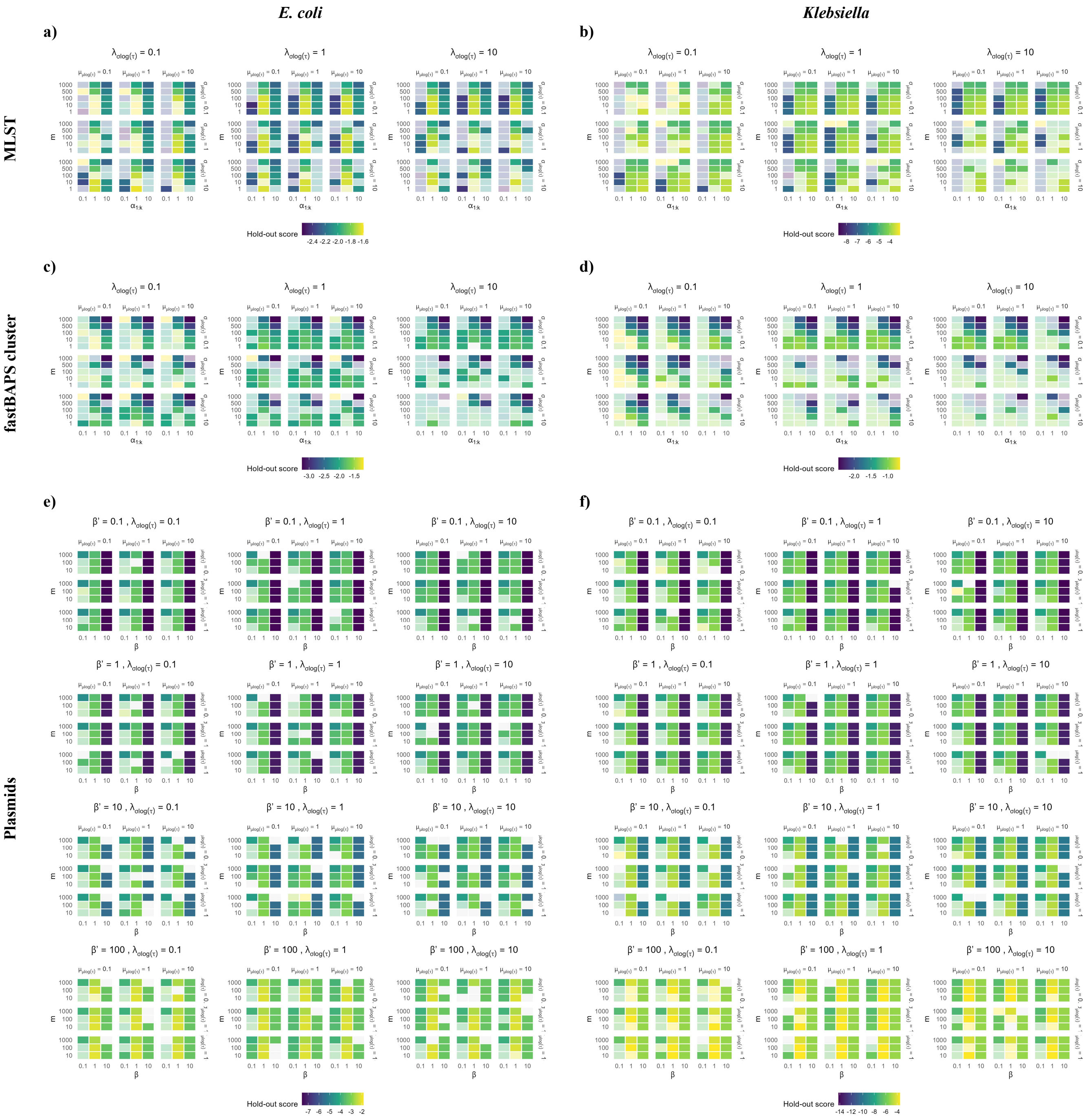
E. coli
Klebsiella
a)
b)
MLST
Hold-out score
c)
d)
fastBAPS cluster
Hold-out score
e)
f)
Plasmids
Hold-out score

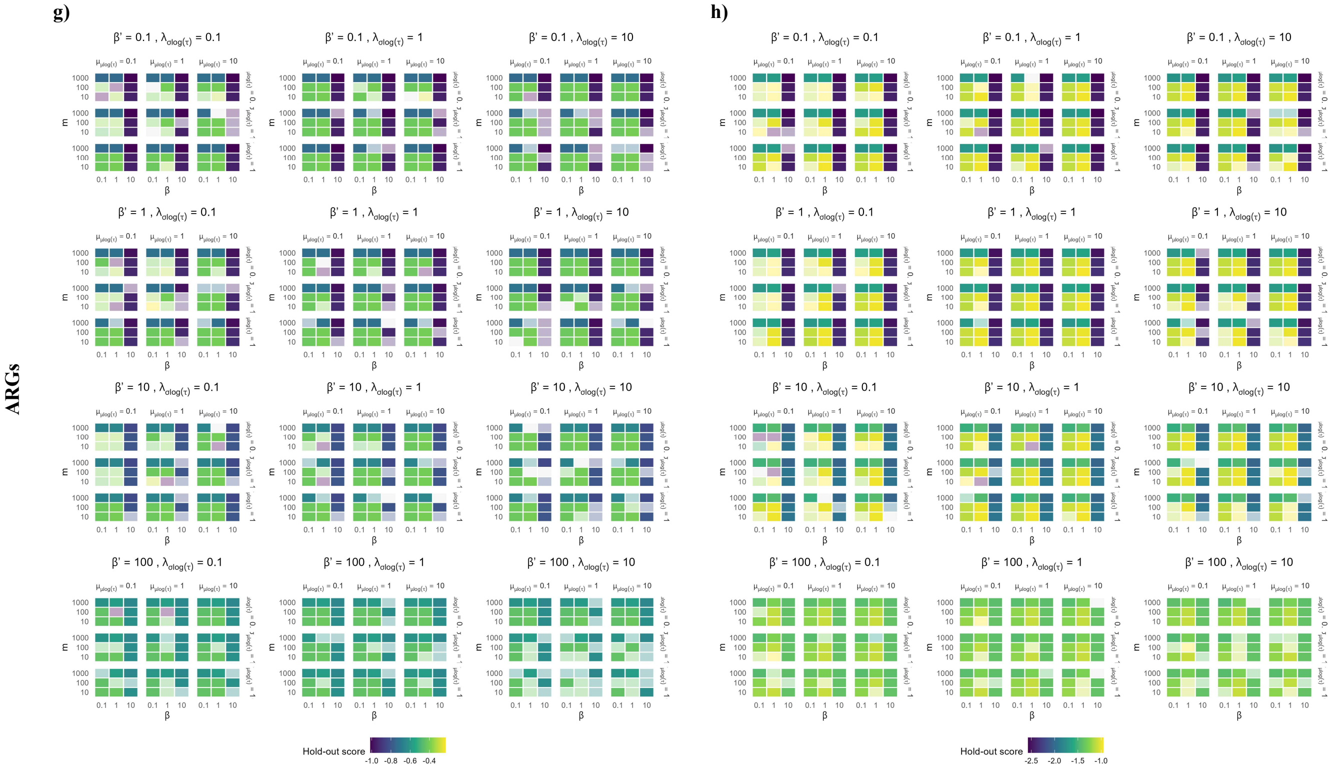

**Supplementary Figure S4: Heatmaps showing predictive performance of prior parameter combinations for hierarchical Bayesian modelling of *E. coli* (left column; a,c,e,g) and *Klebsiella* (right column; b,d,f,h) genomic features, stratified by region**. Predictive performance was quantified using Dirichlet-Multinomial likelihood for MLSTs and fastBAPS clusters, and Beta-Binomial likelihood for plasmids and antimicrobial resistance genes (ARGs), using a regionally-stratified, randomly-selected one third of the NEKSUS dataset as a held-out test set, and fitting model to the remaining two-thirds training set. Genomic features represented are, from top to bottom: MLSTs (**a-b)**), fastBAPS clusters (**c-d)**), plasmids subcommunities (**e-f)**), and ARGs (**g-h)**). Models that failed chain convergence diagnostic checks are shown with reduced opacity (more translucent) tiles (i.e.: divergent transitions and transitions hitting maximum tree depth $\geq$5% of sampling iteration; the third quantile of $\hat{R}$ $\geq$1.05, and lower quantile of effective sample size $\leq$100, which may indicate poor exploration of the posterior distribution by the Markov chain and biased estimates). The log-holdout scored of goodness of fit / predictive performance were truncated at the 2.5$^{th}$ and 97.5$^{th}$ percentiles where all truncated models failed chain convergence diagnostics, to improve visual discrimination of colours of valid models. Blank tiles indicate where the model failed to run for that parameter combination.

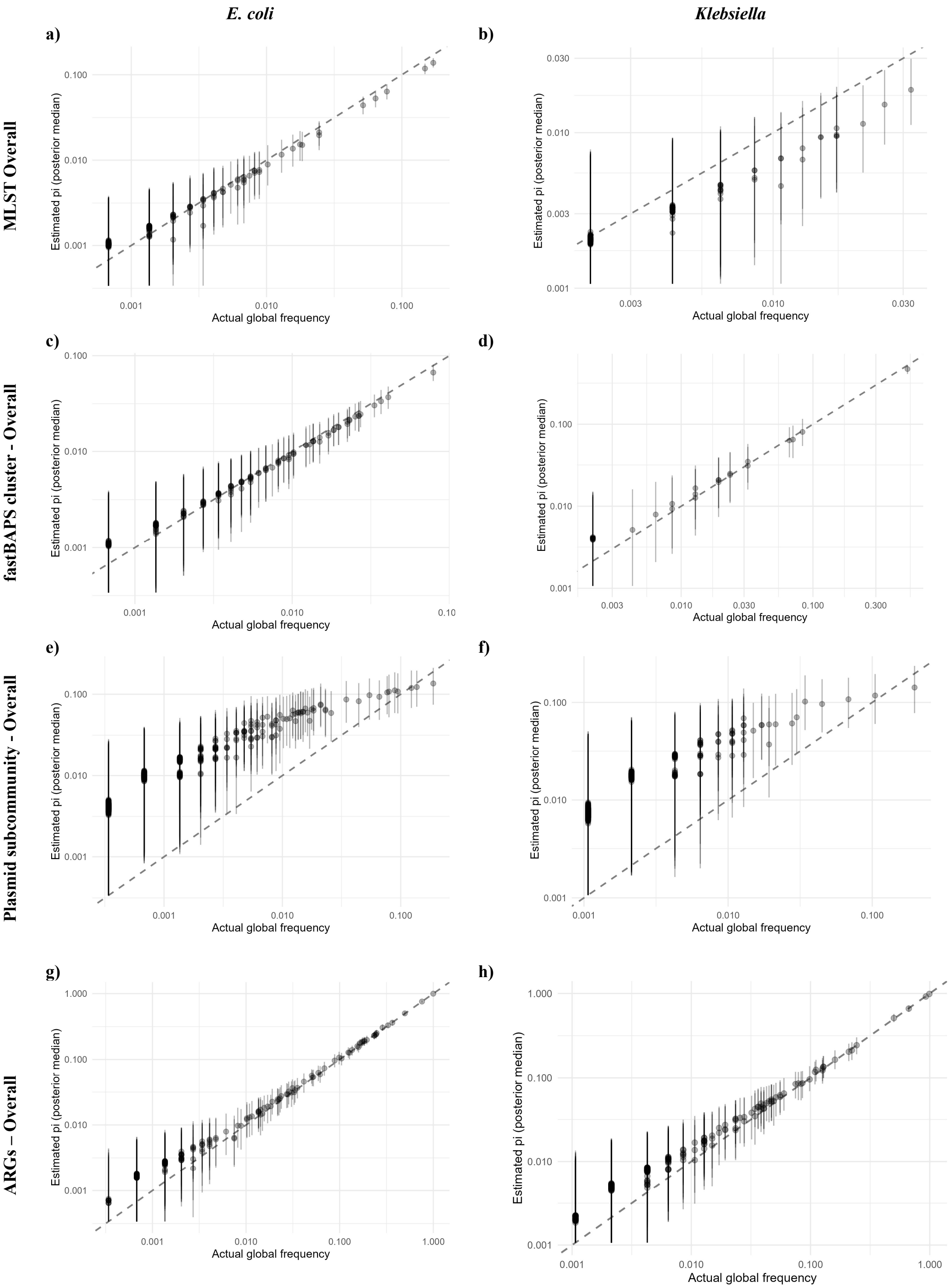

**Supplementary Figure S5: Panel plots showing actual observed global feature frequency in the NEKSUS dataset (x-axis) vs predicted feature frequencies from hierarchical Bayesian modelling (y-axis) which incorporates regionally-stratified data, for *E. coli* (left column; a,c,e,g) and *Klebsiella* (right column; b,d,f,h).** Genomic features represented are, from top to bottom: MLSTs (**a-b)**), fastBAPS clusters (**c-d)**), plasmids subcommunities (**e-f)**), and antimicrobial resistance genes (ARGs) (**g-h)**). Points represent the median posterior estimate, and vertical error bars represent the 95% credible intervals (CrIs).

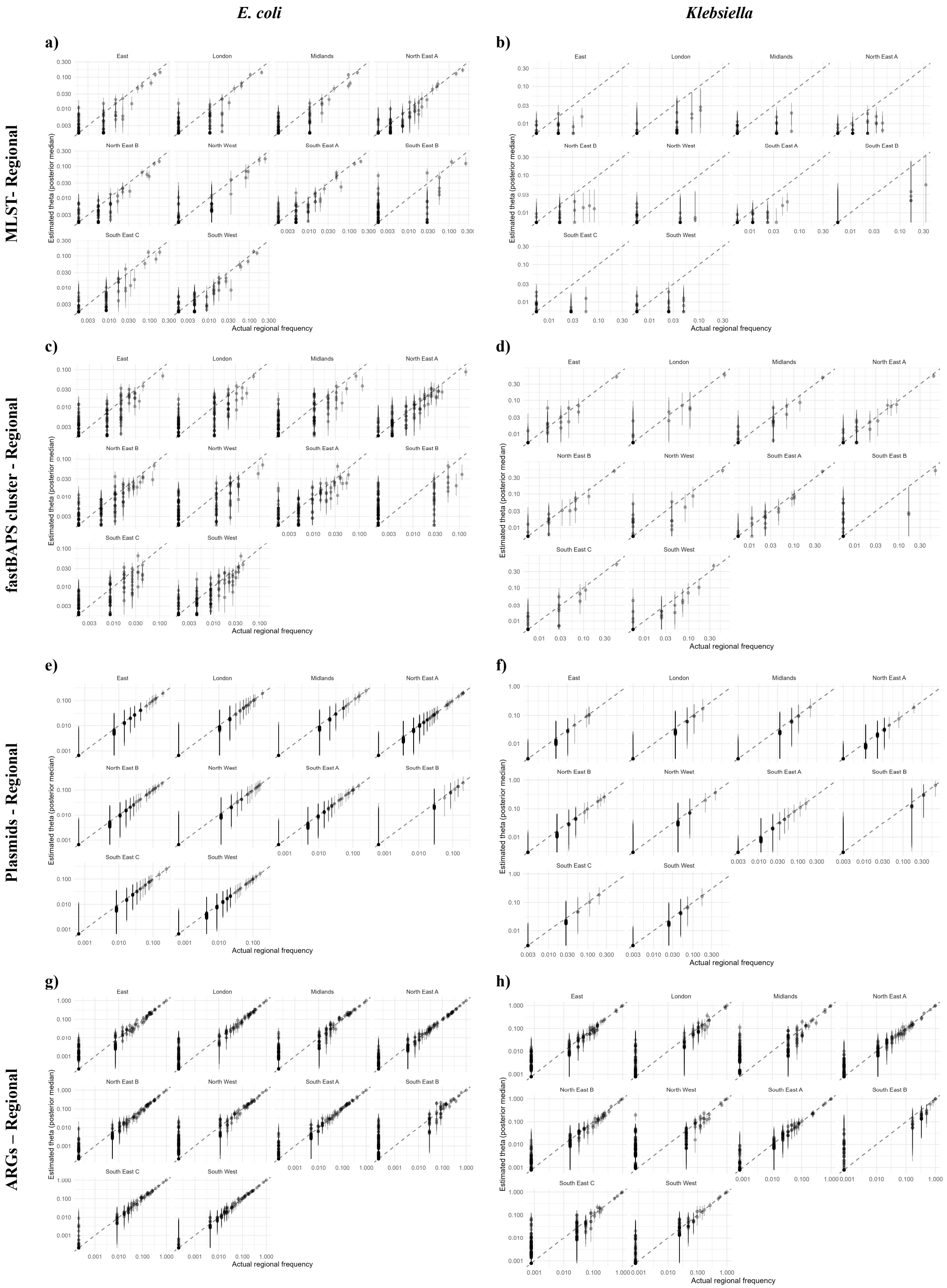

**Supplementary Figure S6: Panel plots showing actual observed <u>regional</u> feature frequency in the NEKSUS dataset (x-axis) vs predicted feature frequencies from hierarchical Bayesian modelling (y-axis) which incorporates regionally-stratified data, for *E. coli* (left column; a,c,e,g) and *Klebsiella* (right column; b,d,f,h).** Genomic features represented are, from top to bottom: MLSTs (**a-b)**), fastBAPS clusters (**c-d)**), plasmids subcommunities (**e-f)**), and antimicrobial resistance genes (ARGs) (**g-h)**). Points represent the median posterior estimate, and vertical error bars represent the 95% credible intervals (CrIs).

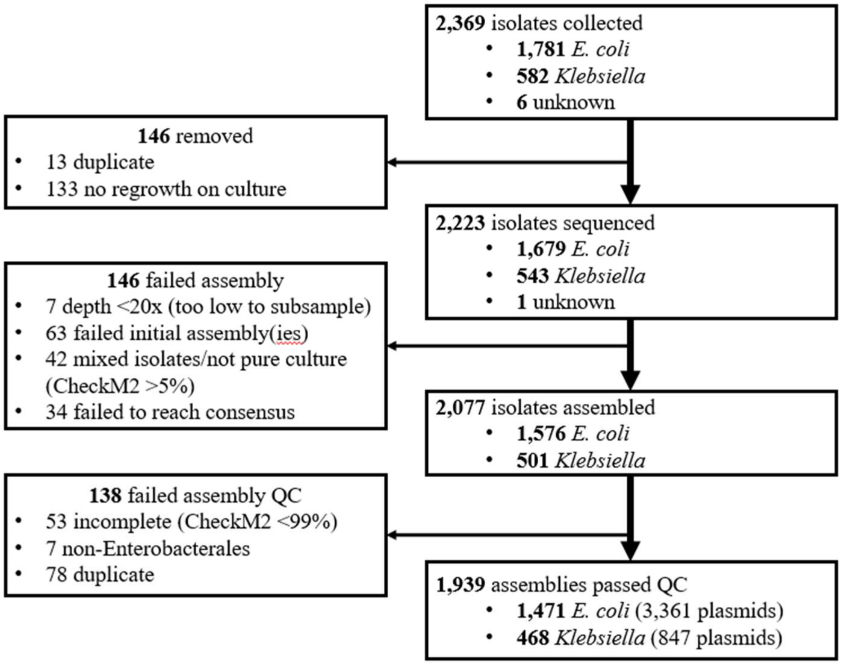

**Supplementary Figure S7: Numbers of *E. coli* and *Klebsiella* bloodstream infection (BSI) isolates collected, sequenced, assembled, and passing assembly quality control (QC) in the NEKSUS study.** A 14-day rolling-window deduplication was applied, such that an isolate was excluded as a duplicate if there was another isolate from the same patient, of the same sample type and the same bacterial species collected within the preceding 14 days. Duplicates were excluded at different stages as metadata was linked at different stages of data processing. Assembly quality control criteria included >95% completeness and <5% contamination using CheckM2.

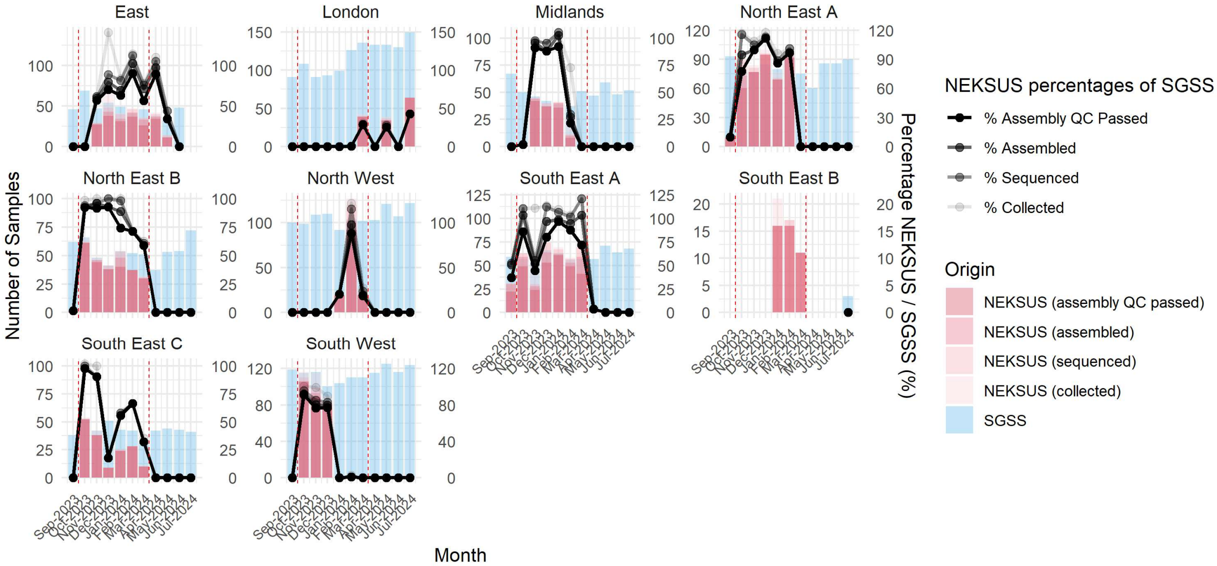

**Supplementary Figure S8: Completeness of collection of *E. coli* and *Klebsiella* bloodstream infection (BSI)-associated isolates in the NEKSUS study, compared to all *Escherichia* and *Klebsiella* app. isolates from blood cultures submitted to the national (non-mandatory) laboratory surveillance system in England (Second Generation Surveillance System [SGSS]).** A 14-day rolling-window deduplication was applied, such that an isolate was excluded as a duplicate if there was another isolate from the same patient, of the same sample type and the same bacterial species collected within the preceding 14 days. Dashed vertical red lines show the period during which NEKSUS aimed to collect isolates. Note that no BSI isolates were retrieved from SGSS in the study period from the South East B region, due to electronic feed failure, and percentages were therefore not calculated with a 0 denominator.

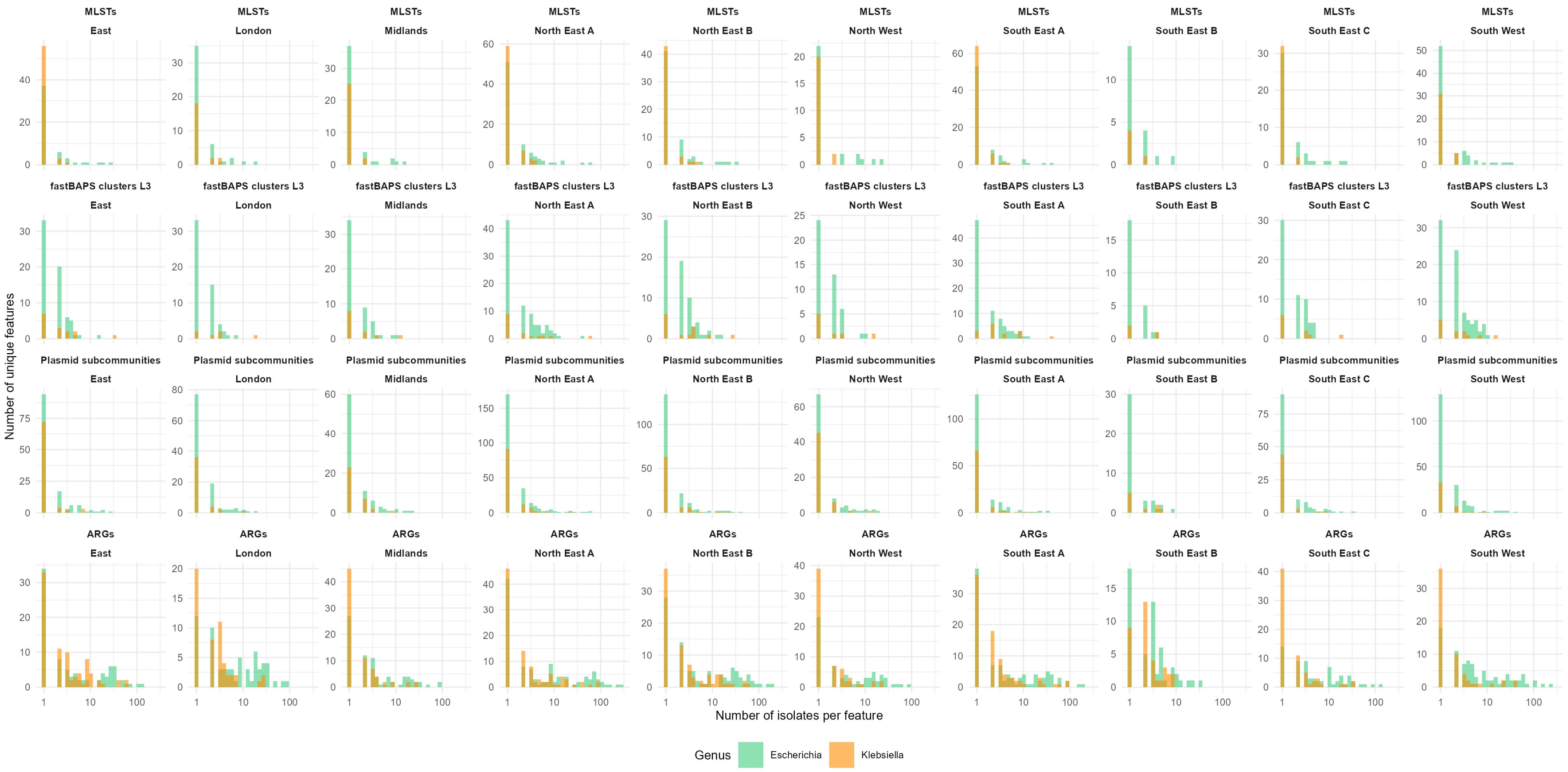

**Supplementary Figure S9: Frequency histograms of genomic features annotated in the NEKSUS dataset, stratified by region.** Features shown are, top to bottom rows: MLSTs, fastBAPS clusters, plasmid subcommunities, and antimicrobial resistance genes (ARGs), stratified by region for *E. coli* (green) and *Klebsiella* (orange). The x-axis is on the log scale.

a)

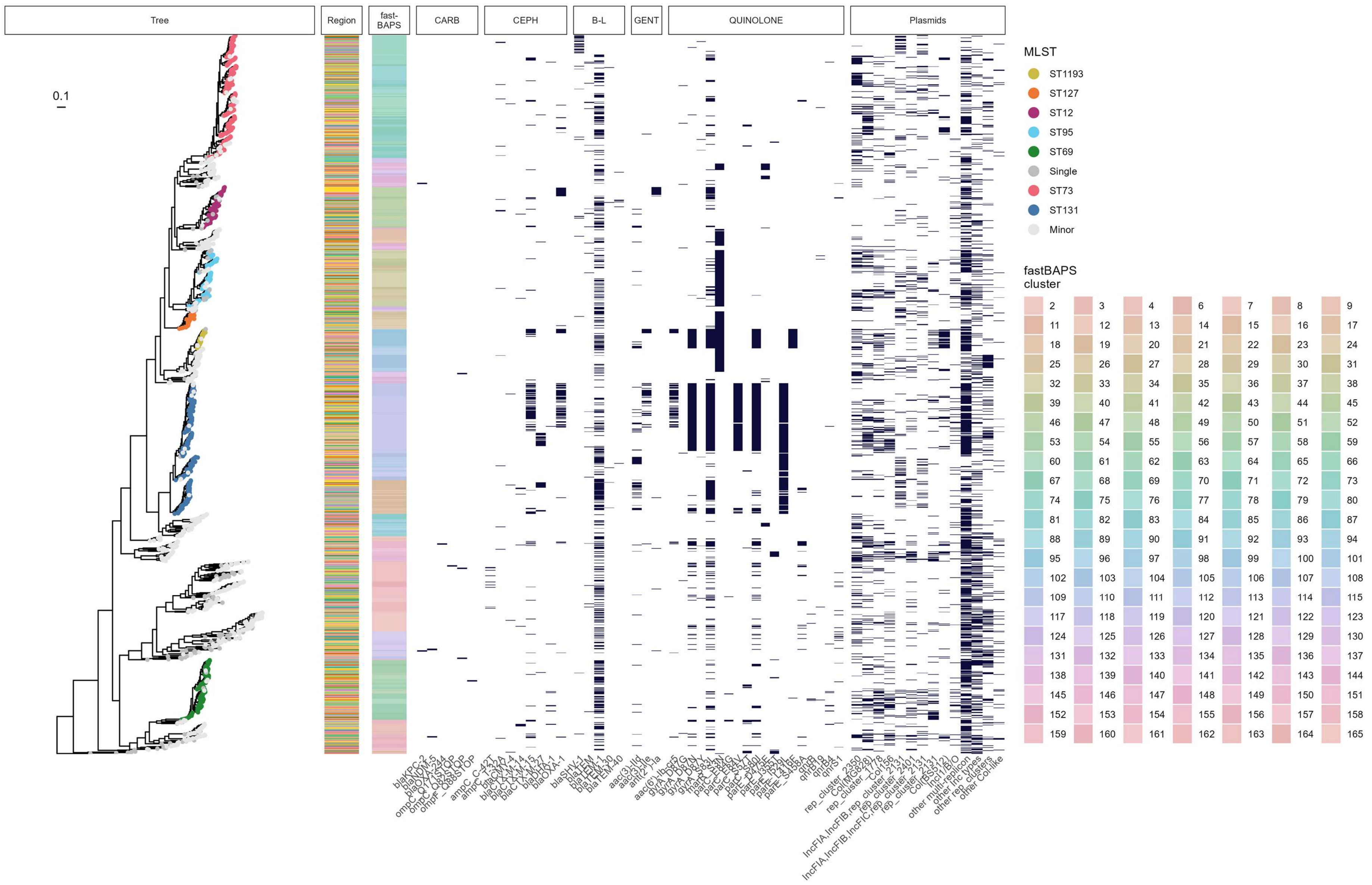

Tree
Region
fast-BAPS
CARB
CEPH
B-L
GENT
QUINOLONE
Plasmids
0.1
blaKPC-2
blaNDM-5
blaOXA-244
ompC_Q171STOP
ompC_Q82STOP
ompF_Q88STOP
ampC_C-42T
ampC_T-32A
blaCMY-4
blaCTX-M-14
blaCTX-M-15
blaCTX-M-27
blaDHA-1
blaOXA-1
blaSHV-1
blaTEM
blaTEM-1
blaTEM-30
blaTEM-40
aac(3)-IId
aac(3)-IIe
ant(2'')-Ia
aac(6')-Ib-cr5
qnrB
qnrB19
qnrB4
qnrS1
rep_cluster_2350
Col(MG828)
rep_cluster_1778
Col156
IncFIA,IncFIB,rep_cluster_2131
rep_cluster_2401
IncFIA,IncFIB,IncFIC,rep_cluster_2131
Col(BS512)
IncI1/B/O
other multi-replicon
other Inc types
other rep_clusters
other Col-like
MLST
ST1193
ST127
ST12
ST95
ST69
Single
ST73
ST131
Minor
fastBAPS cluster
2
3
4
6
7
8
9

b)

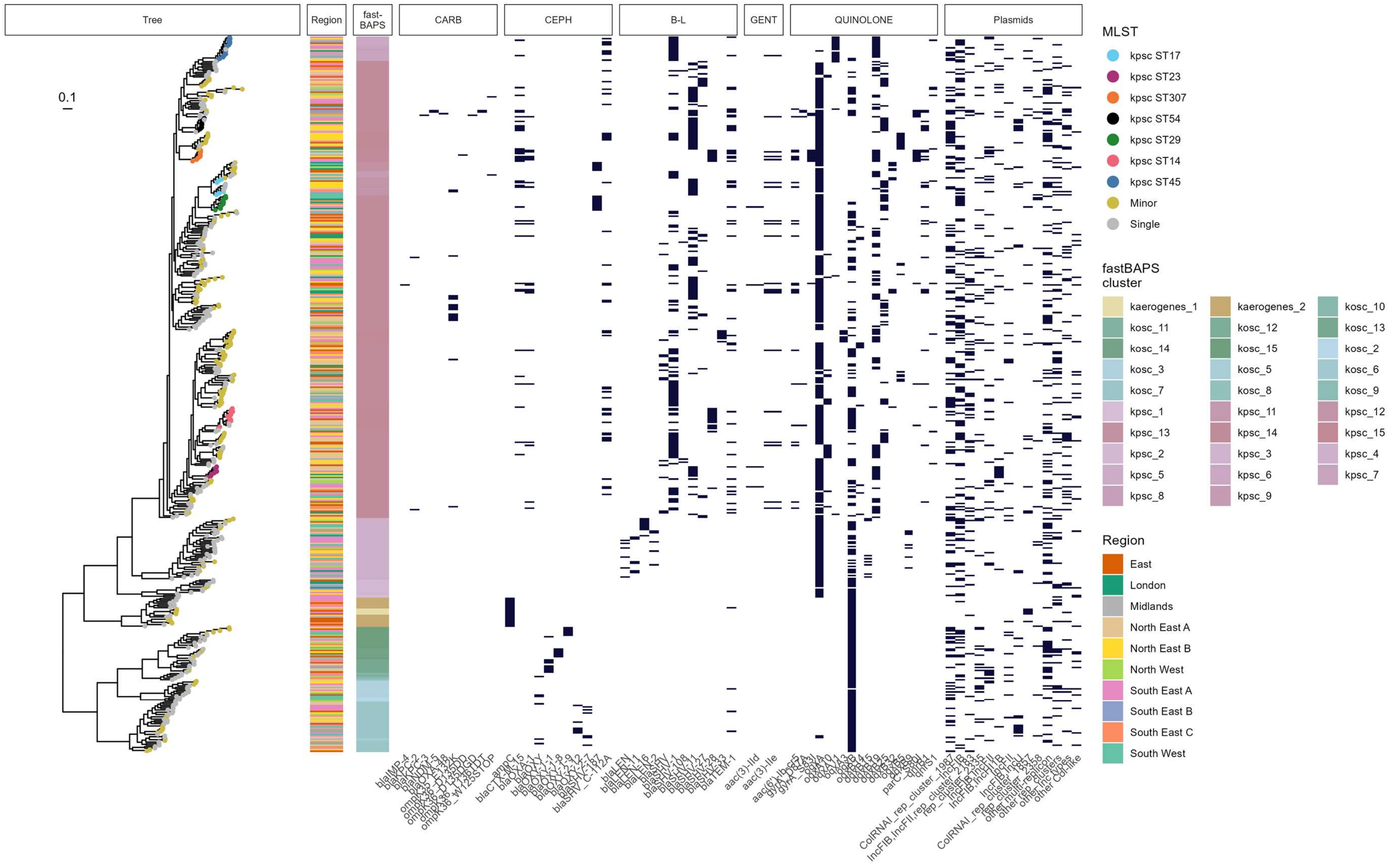

**Supplementary Figure S10: Phylogenetic trees of a) *E. coli*, b) *Klebsiella pneumoniae* species complex, and c) *Klebsiella oxytoca* species complex bloodstream infection (BSI)-associated isolates in the NEKSUS dataset.** Tree tip colours show Multi-Locus Sequence Type (scheme), and metadata panels show region of collection, suspected healthcare exposure of acquisition, key antimicrobial resistance gene and plasmid associations.

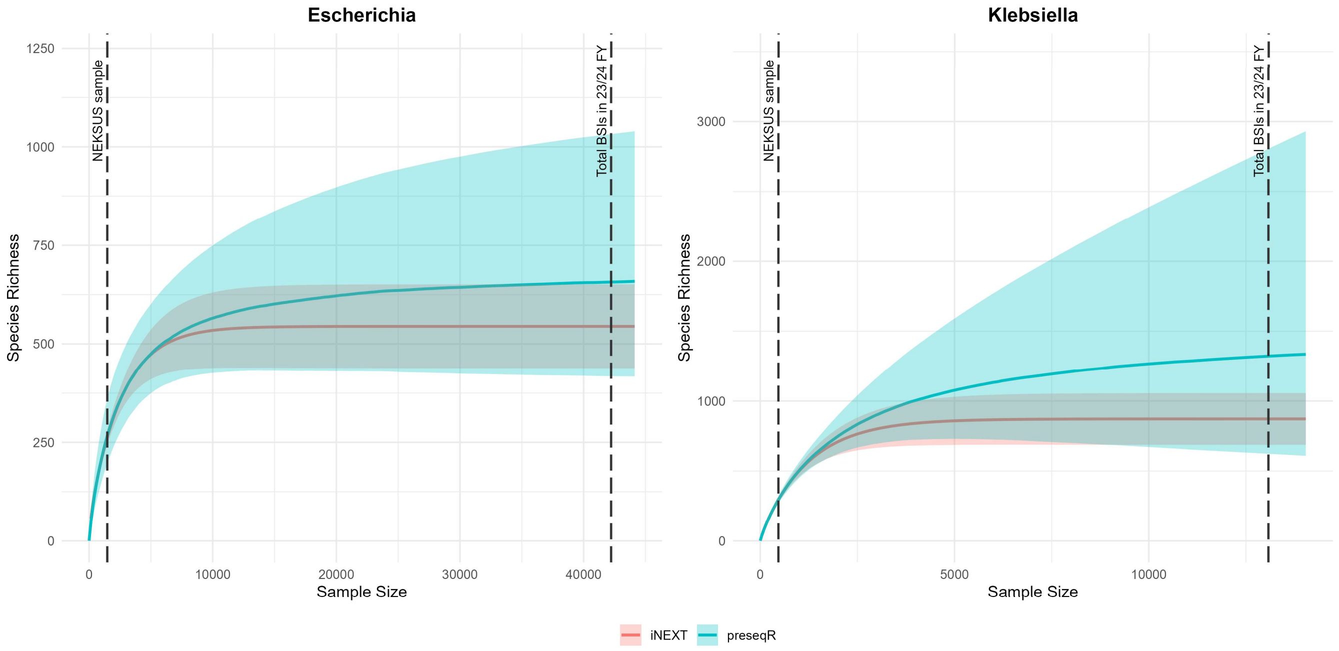

**Supplementary Figure S11: Estimates of the number of unique MLSTs (species richness) in *E. coli* (left) and *Klebsiella* spp. (right) bloodstream infection (BSI) isolates, using two of the best ecological diversity estimators reviewed by Schmitz et al., 2025.[32] (preseqR,[31] and iNEXT[30]).** The dashed vertical lines on the left of each plot represent the actual collected sample size in the NEKSUS study, 1,471 *E. coli* and 468 *Klebsiella* spp isolates, and on the right, the total number of BSIs for the respective genera occurring in the 2023/24 financial year[33] (42,224 *E. coli* and 13,078 *Klebsiella* spp. BSIs). The portion of the iNEXT line left of the actual collected NEKSUS sample size is identical to rarefaction (down-sampling) of the NEKSUS data. iNEXT is known to systematically underestimate species richness when the extrapolated sample is two or more times the actual collected sample.[31,32]

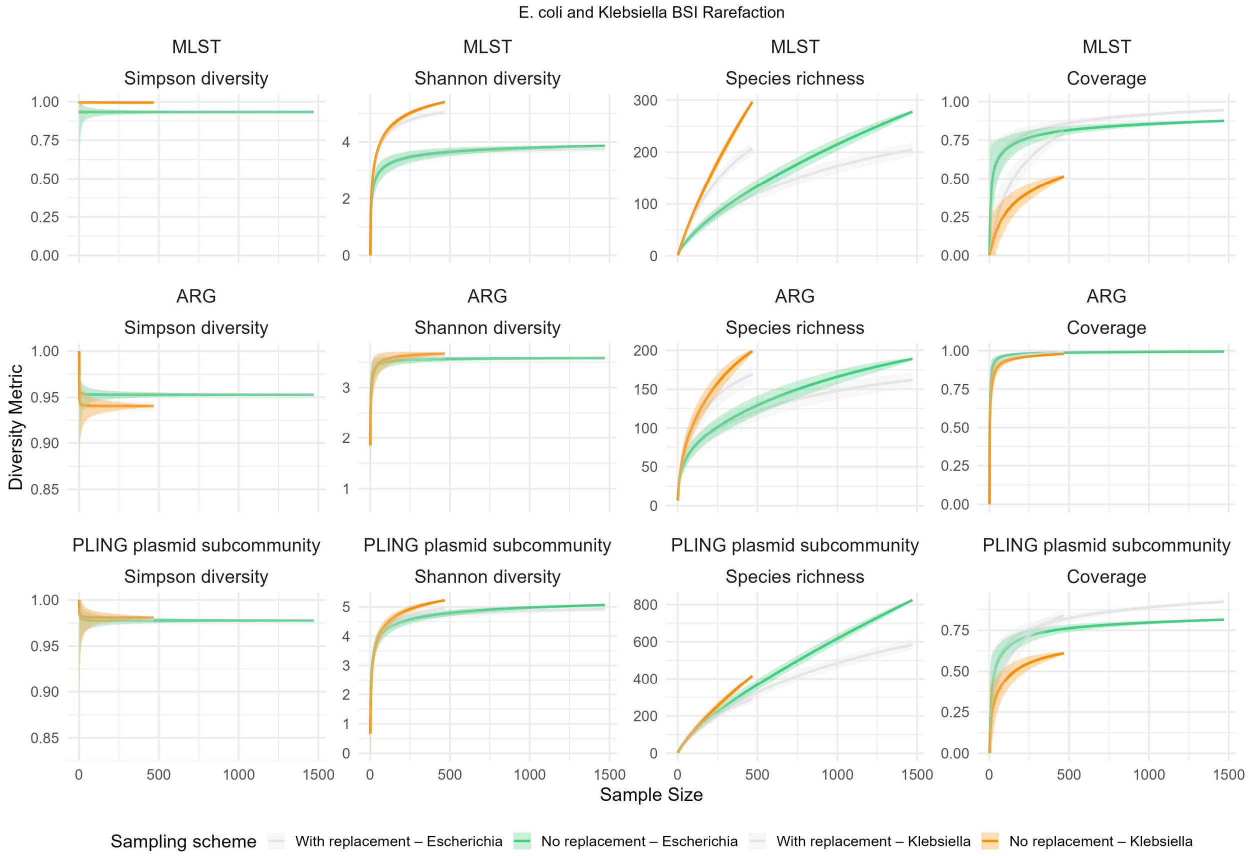

**Supplementary Figure S12: Rarefaction (sampling without replacement) from the *E. coli* (green), and *Klebsiella* (orange) bloodstream infection (BSI)-associated isolates collected in the NEKSUS study.** Rows represent different genomic features (multi-locus sequence type [MLST], top, antimicrobial resistance genes (ARGs), middle, and plasmid PLING subcommunities [DCJ-indel threshold ≤4], bottom). Columns correspond to different measures of genetic diversity (from left to right, Simpson diversity, Shannon diversity, Species richness, and coverage). Coverage is defined as the proportion of the population represented by features that are frequent enough to be detected at least once in the sample, and is estimated using the Good-Turing estimator ($C = 1-f_1/N$ – i.e.: the proportion of the sample not made up of singleton species). Samples for each genus are drawn up to the maximum number of isolates collected in the NEKSUS study (1,471 for *Escherichia* and 468 for *Klebsiella* spp.). Light grey lines show corresponding rarefaction curves for sampling *with* replacement. Note that the grey (sampling *with* replacement) curves are systematically lower for species richness when sampling from the finite population of the NEKSUS sample, as common 'species' (features) are repeatedly sampled, while they are higher for coverage for the same reason, as the proportion of the population made up of singleton species is lower.

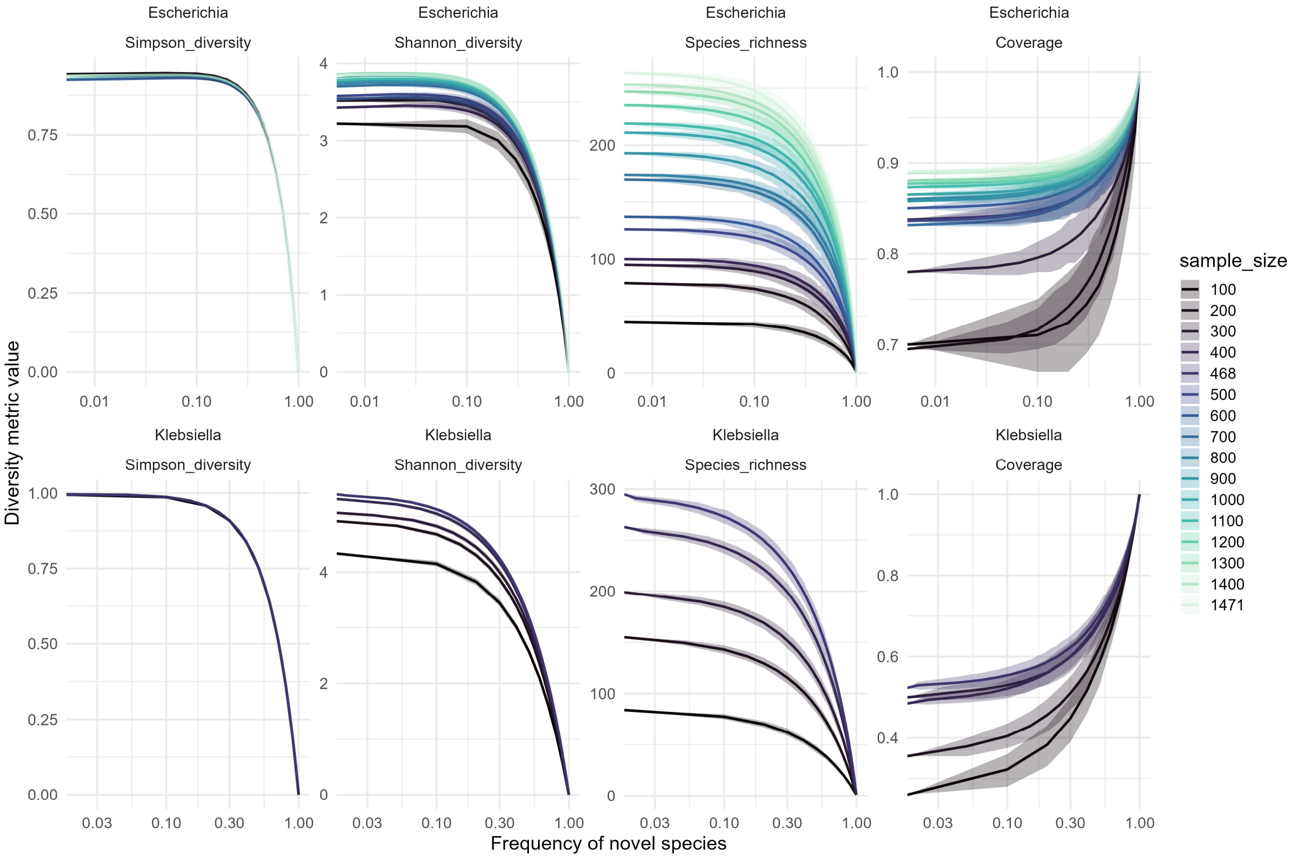

**Supplementary Figure S13: Simulated incursion of a novel MLST into the NEKSUS sample to demonstrate sensitivity of different diversity measures to detecting 'emergent' lineages.** The incursion of novel MLSTs was simulated by randomly replacing 1, 2, 3, etc. isolates in the NEKSUS sample with a hypothetical 'novel MLST', at various sample sizes. Although this is a highly simplified scenario, as most lineage incursions historically have only caused a transient disruption in population structure[63,64], this demonstrates that Shannon and Simpson diversity (left two columns) are insensitive to detecting incursions, as the 'novel' species/feature has to reach >10% before these measures change. For comparison, only three of the dominant *E. coli* MLSTs occur at a frequency of above 10%.

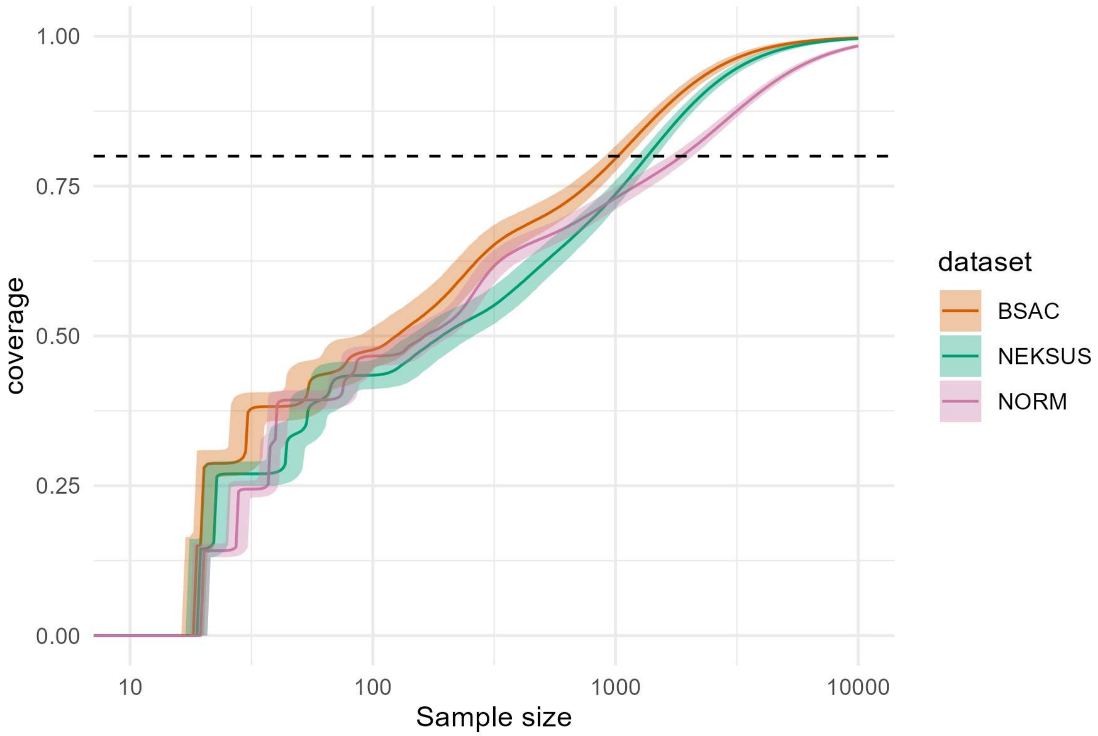

**Supplementary Figure S14: Sequencing sample size estimates (solid lines) with 95% credible intervals (ribbons) obtained from Bayesian modelling to reach different MLST coverages, applied to different external genomic surveys of *E. coli* BSIs.** Coverage represents the proportion of the population that has a feature (i.e.: MLST) that has been detected at least once in the sample. BSAC refers to the Britch Society for Antimicrobial Chemotherapy[64], and NORM refers to the Norwegian surveillance program on resistant microbes[63]. The dashed horizontal line refers denotes 80% coverage.
